# Feasibility trial of a new digital training package to enhance primary care practitioners’ communication of clinical empathy and realistic optimism

**DOI:** 10.1101/2024.10.11.24315303

**Authors:** Felicity L Bishop, Jeremy Howick, Jane Vennik, Jennifer Bostock, Paul Little, Christian Mallen, Leanne Morrison, Mary Steele, Beth Stuart, Stephanie Hughes, Kirsten Smith, Mohana Ratnapalan, Emily Lyness, Hajira Dambha-Miller, Riya Tiwari, Clare Lockyer-Stevens, Hazel Everitt

## Abstract

**Background:** Patients can benefit when primary care practitioners communicate clinical empathy and optimism during consultations, but previous training interventions for practitioners are overly time-consuming and evidence on patient outcomes is limited. This study assessed the feasibility of a cluster-randomized controlled trial in UK general practice to evaluate effects of a new brief digital learning package in empathy and optimism (EMPathicO) for primary care practitioners.

**Methods:** The study ran January to October 2020, with COVID-19 related modifications (mostly, practitioner and patient data had to be collected separately) from March 2020. 9 practices and 12 primary care practitioners recruited from UK (Southern England, Midlands). 12 practitioners completed EMPathicO training and 11 completed qualitative telephone interviews. Patients recruited through social media completed web-based questionnaires at baseline (<2 weeks post- consultation) and 2-week follow-up (n=437). Purposively sampled patients completed qualitative telephone interviews (n=30). Data analysed descriptively and thematically.

**Results:** Practitioners were keen to reflect on and enhance communication skills and were willing to undertake digital training, even during COVID-19 pandemic. However, some practices and practitioners would have declined if video-recording consultations was a mandatory aid to reflection during training. Practitioners found EMPathicO brief, relevant and engaging and could implement techniques taught in the training. Patients found the online questionnaires acceptable, though retention was suboptimal at 57%; minor easily remedied feasibility and process issues were identified (including incentivizing participation); and patients were enthusiastic about research to improve communication.

**Conclusions:** An agile research strategy enabled useful feasibility data to be collected despite the challenges of the COVID pandemic. It is feasible to proceed to a full trial of the effects of EMPathicO on patient outcomes in primary care, if video-recording consultations is optional not mandatory.

Feasibility work to develop and test sophisticated questionnaire structures is valuable when planning primary care patient surveys.

## Introduction

Patient-practitioner communication is often sub-optimal,[1, 2] and improving practitioner communication skills can improve patients’ symptoms, quality of life, adherence to and satisfaction with care, producing modest benefits that are comparable to many pharmaceutical interventions.[3–5] Improved communication may also reduce the risk of worsening quality of life and symptom management, unwanted prescriptions and non-adherence;[6, 7] unnecessary economic costs;[7] deviation from guideline-recommended treatment;[8] and complaints and litigation.[9, 10] In particular, there is scope to better harness the benefits of communicating clinical empathy and positive expectations[11] using verbal <u>and</u> non-verbal communication in primary care.[12–14] While practitioners are willing to engage in brief training,[14, 15] few interventions have been tested clinically for effects on patients’ health,[16] or are sufficiently well-described and brief to be implementable in over-burdened, pressurised, primary care settings.[17–19]

We recently developed a new brief digital training package for primary care practitioners (PCPs), called EMPathicO (see Fig 1). This training package is designed for PCPs (including GPs, nurses, physiotherapists, and others) to enhance their communication of clinical empathy and realistic optimism through verbal and non-verbal behaviours. To develop EMPathicO we used the systematic multi-component person-based approach (PBA) to put intervention users and beneficiaries at the heart of the design and development process[20] and integrated evidence- and theory-based approaches[21] to ground our training package. A full account of how we developed EMPathicO using the LifeGuide open-source software for creating digital health interventions[22] and its basis in evidence, theory, and users’ experiences, is presented elsewhere.[23] Fig 2 presents the underpinning logic model summarising our theory of how EMPathicO could change PCP communication behaviours which in turn could change patient expectancies, affect and cognitions and subsequent patient outcomes (adapted from [23, 24]).

**Fig 1.**
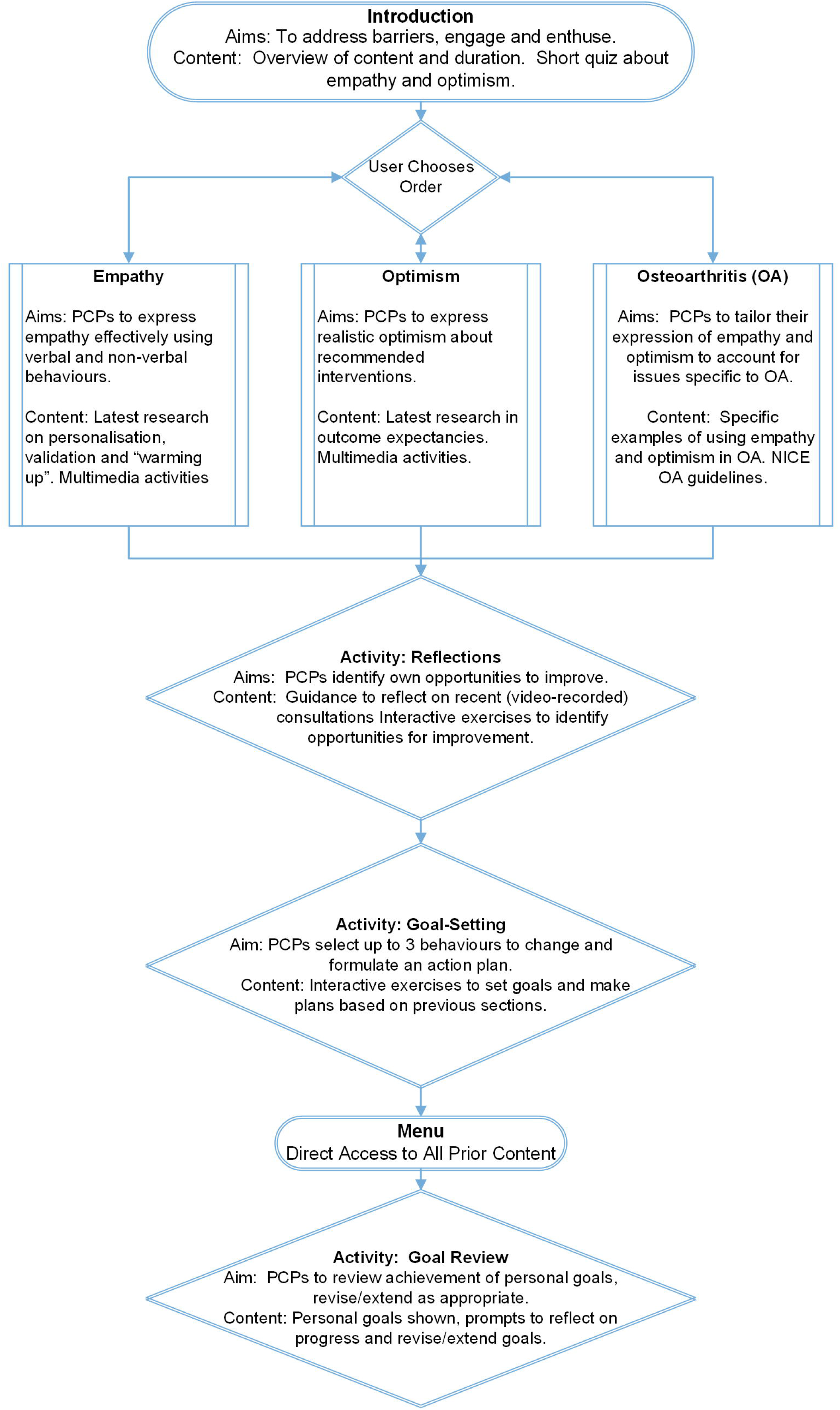
Summary of EMPathicO Digital Training Intervention

**Fig 2.**
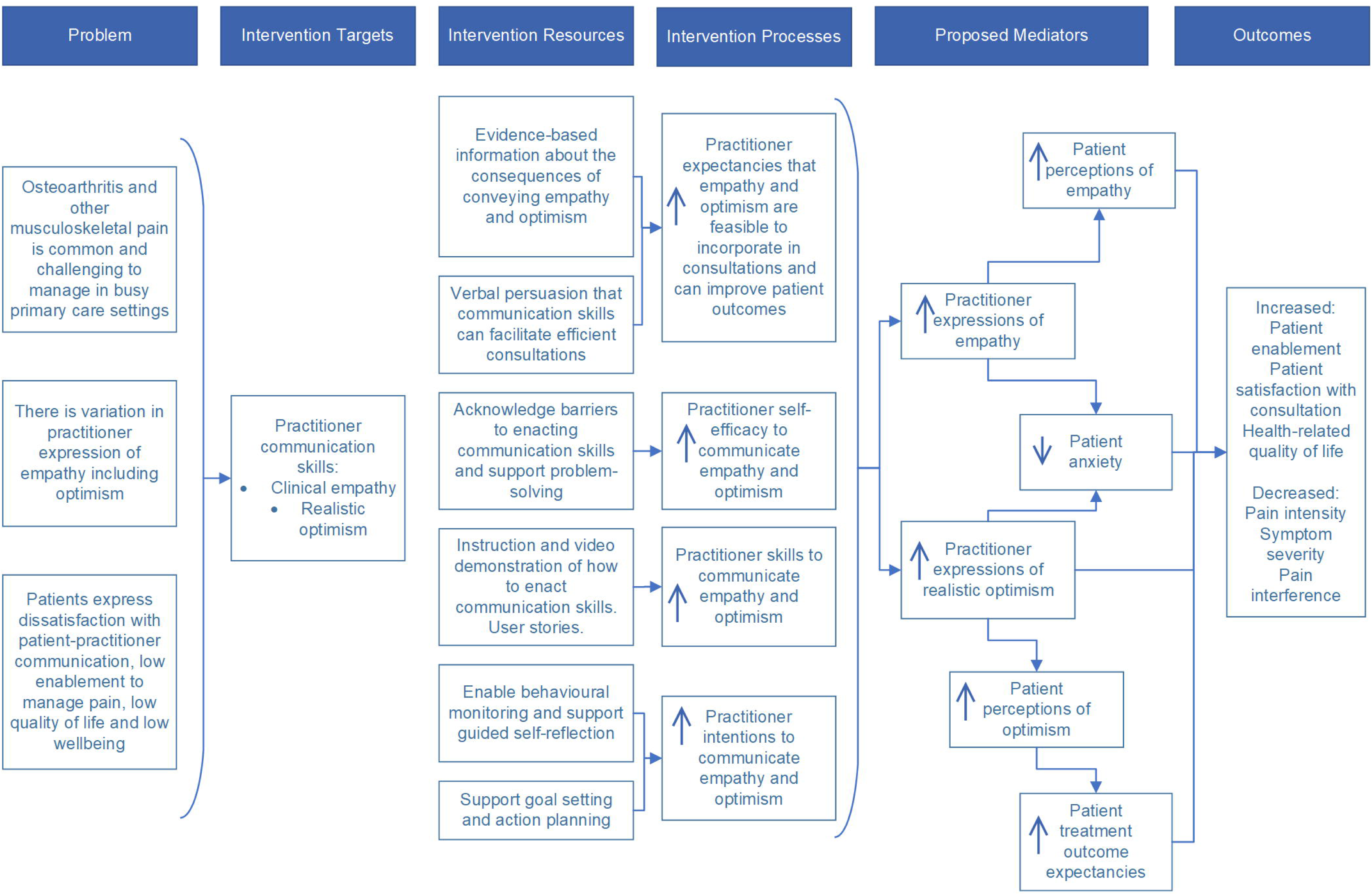
Logic Model for EMPathicO

This study was designed to test the feasibility of evaluating EMPathicO in a cluster-randomized controlled trial in UK general practices, with two groups of patients – those consulting for OA and those consulting for other reasons. Cluster-randomization at the level of general practices was chosen because randomizing individual PCPs risks cross-contamination within practices if practitioners discussed the training with each other. A randomized feasibility trial was chosen because this would best enable us to test the feasibility of recruitment and randomization methods and recruitment and retention rates for the planned full trial. Two groups of patients were included because: interventions targeted to specific audiences and conditions are likely to be more relevant to recipients[25] (i.e., our PCPs) and changes in communication skills are likely to ‘spill over’ into PCPs’ wider practice and thus benefit more patients, even if examples in the intervention are focused on a specific condition. OA was chosen as it is common - approximately ten percent of UK adults had OA in 2017^2^ - it is primarily managed through general practice, NICE Guidelines recommend a patient-centred approach, and evidence suggests that OA is likely to be responsive to empathic, optimistic communication.[2, 11, 26]

The aims were: to establish methods to maximise recruitment and minimise attrition of practices and patients in practices with a range of socio-demographic areas; to identify feasible randomisation and consent procedures and finalise inclusion/exclusion criteria; and to finalise outcome and process measures.

## Methods

### Ethical Approval

Ethical approval for our original study design was granted by the South Central – Hampshire B Research Ethics Committee on 6th December 2019 (19/SC/0553). The sponsor reviewed and approved the modified study design on 31^st^ March 2020 (ERGO number 52146). An amendment to restart study activity in practices was approved by the Research Ethics Committee on 13^th^ July 2020.

### Design

A mixed methods feasibility study was designed to evaluate methods for a cluster-randomised trial of EMPathicO in patients with hip /or knee OA and a wider sample of all-consulters. The study is reported in accordance with applicable reporting guidelines and checklists are available in supplementary materials: TIDieR (S1),[27] Consort pilot and feasibility trials extension (S2),[28] Consort conserve extension (S3).[29]

### Original Study Design

This original study design was a cluster-randomized trial allocating practices to EMPathicO or no-intervention control using a 1:1 ratio; data were to be collected from practitioners and their patients via video-recordings of consultations, qualitative focus groups and interviews, and self-report questionnaires. This original study design is summarised in Fig 3 with planned methods in S4 Appendix. The original study design was implemented from January to February 2020, during which 5 practices, 8 practitioners, and no patients were recruited, two practices (three practitioners) completed baseline activities and were randomised, and one practitioner completed the intervention.

**Fig 3:**
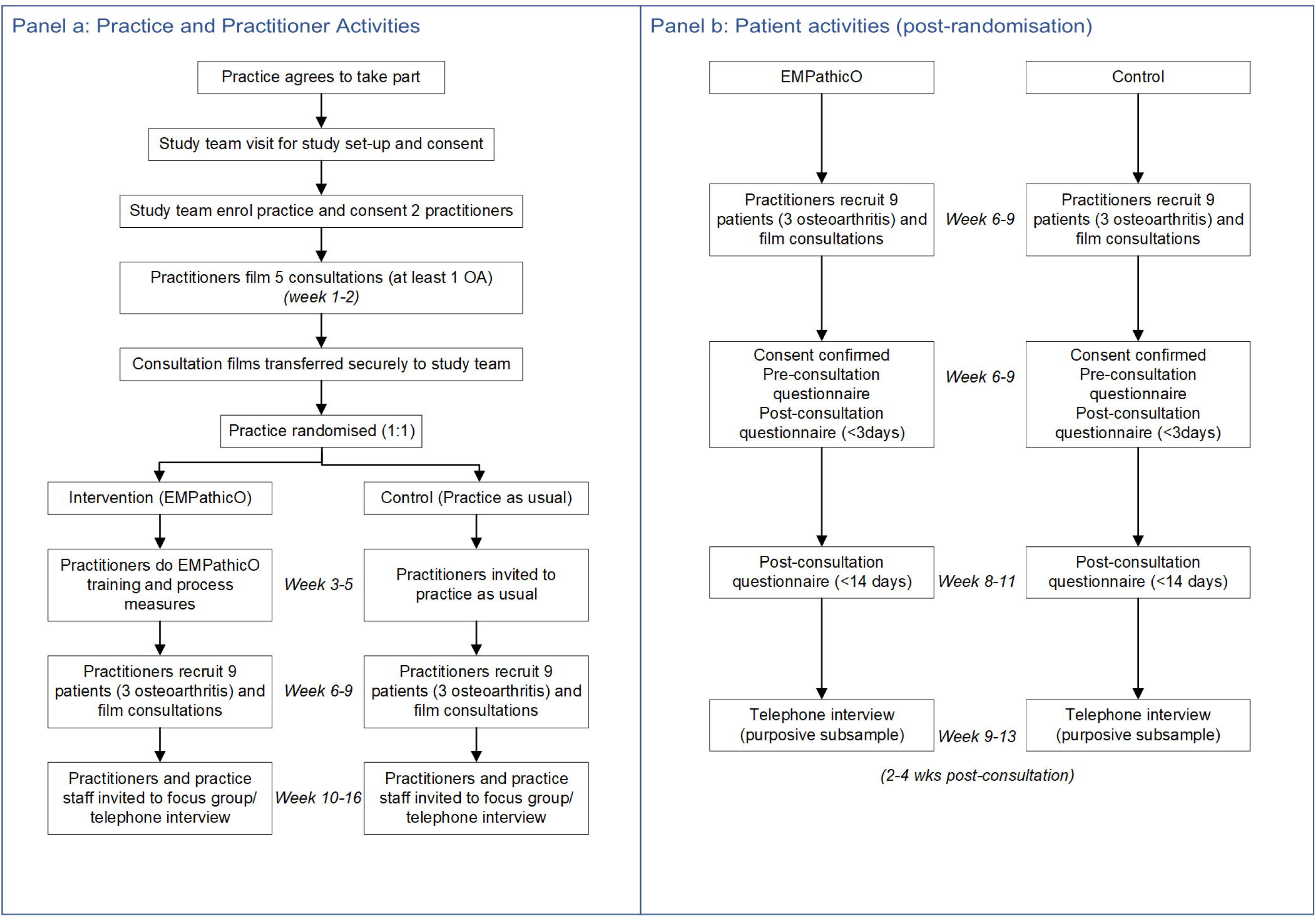
Original Study Design

### Extenuating Circumstances

On 19^th^ March 2020, the NIHR paused all non-essential clinical research in response to the COVID-19 pandemic.[30] Therefore, we ceased activity with enrolled practices/practitioners and modified our research design to meet as many aims and objectives as possible within our funding window without involving practices in patient recruitment. Modifications were planned by the research team and reviewed by the sponsor. Previously enrolled practices and practitioners were transferred to the modified study design on regaining capacity for research. It was not possible to return to the original study design because capacity for research in primary care remained extremely limited.

### Modified Study Design

The modified study design entailed two separate studies. One mixed methods study with practitioners comprised a qualitative study with embedded quantitative data. Practitioners worked through the intervention and were interviewed about their experiences of the intervention and views on the planned trial; quantitative intervention usage data was captured and described.

One mixed methods study with patients comprised an online survey with an embedded qualitative component. In the online survey patients completed process and outcome measures at two time- points via a web-based questionnaire. In the qualitative study a purposive sub-sample of survey respondents took part in a single qualitative interview about their experiences of the questionnaires and their recent primary care consultations.

The modified study design is summarised in Fig 4, study components are mapped to objectives in Table 1 and the methods are described below. Important modifications were: all practitioners allocated to the intervention; patients recruited via social media within two weeks following a self-reported primary care consultation with any PCP; no pre-consultation patient reported measures; no filming of consultations; no practice staff interviews; one-to-one interviews with practitioners instead of focus groups. Additional items were added to the patient survey asking about the impact of COVID on participants’ survey responses and work situation. Furthermore, part-way through the patient survey we ceased collecting data on two outcome measures (the HADS and the SF-12) as we recruited more patients to the survey than planned for the original study design and exhausted our licensed administrations. The modified study design was implemented from May to October 2020. The protocol is available in S5.

**Fig 4:**
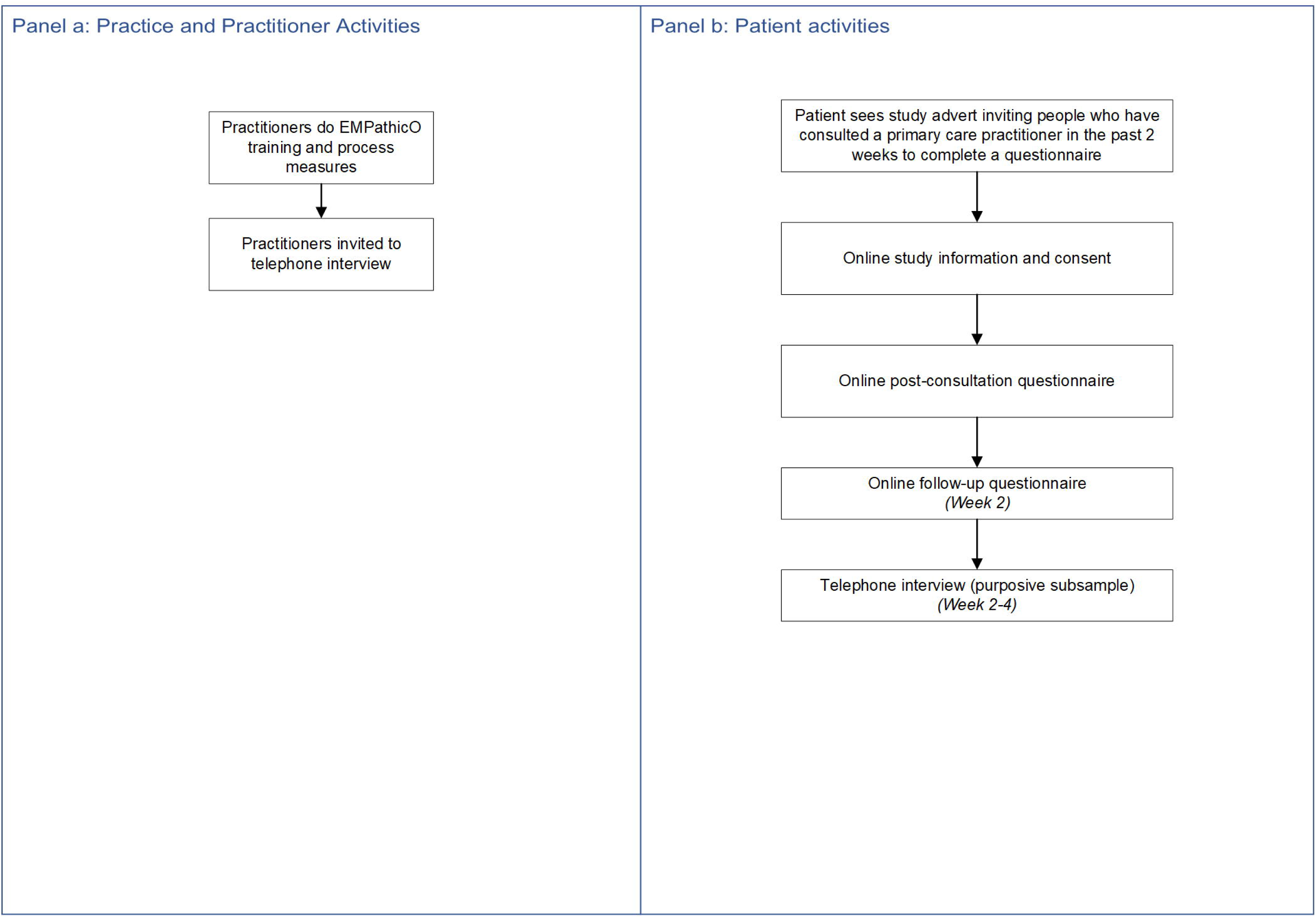
Modified Study Design

**Table 1.**
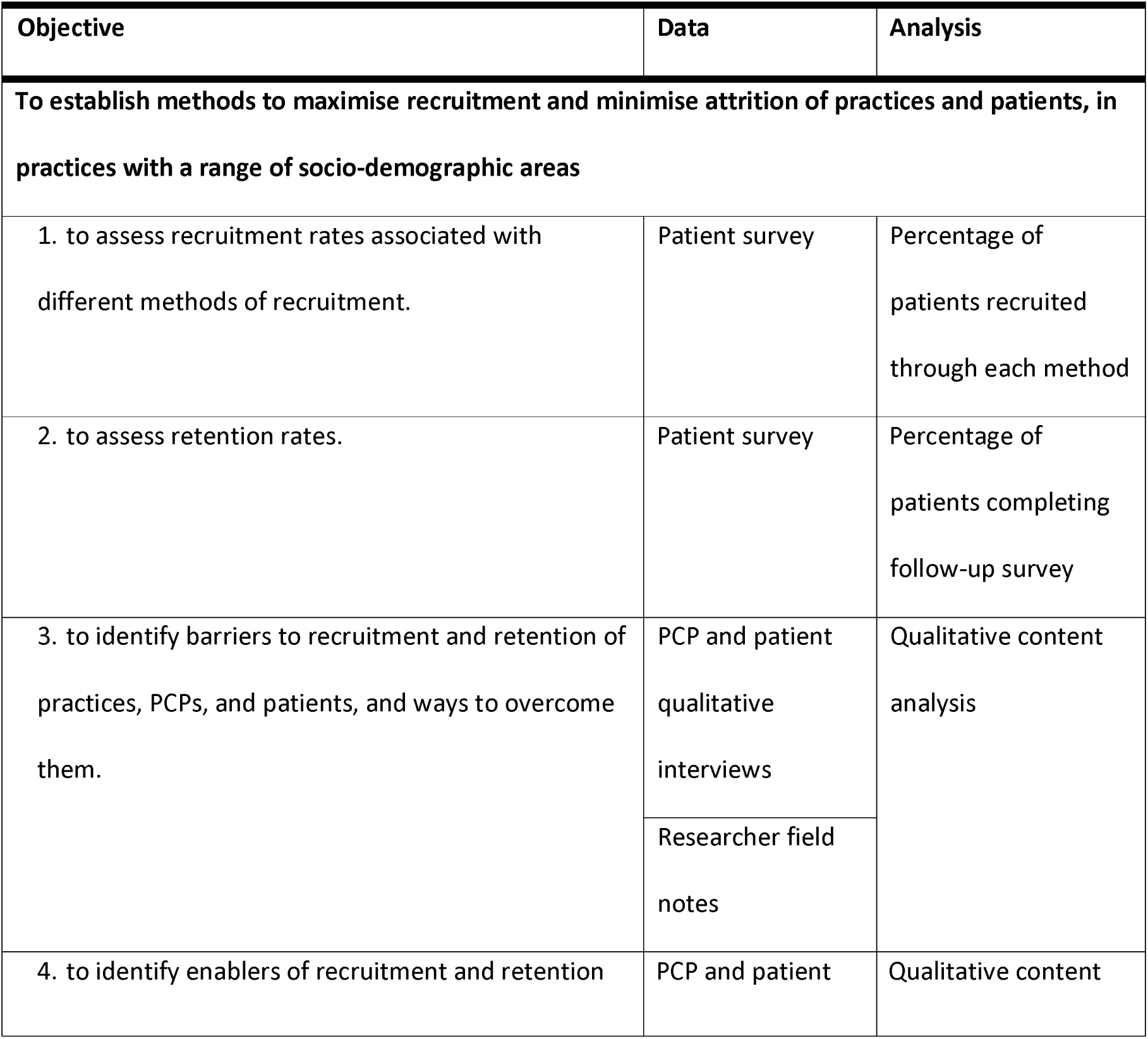
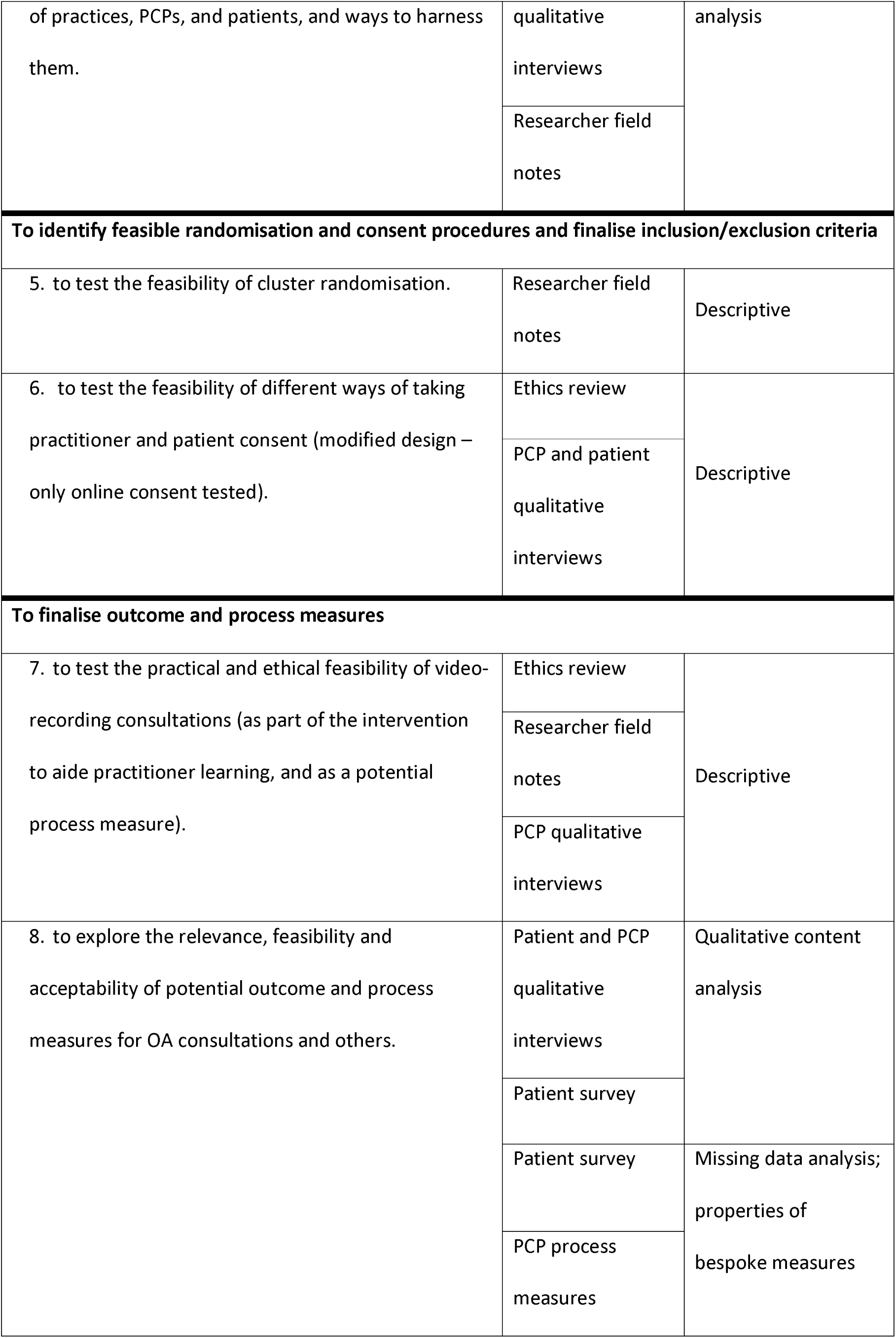
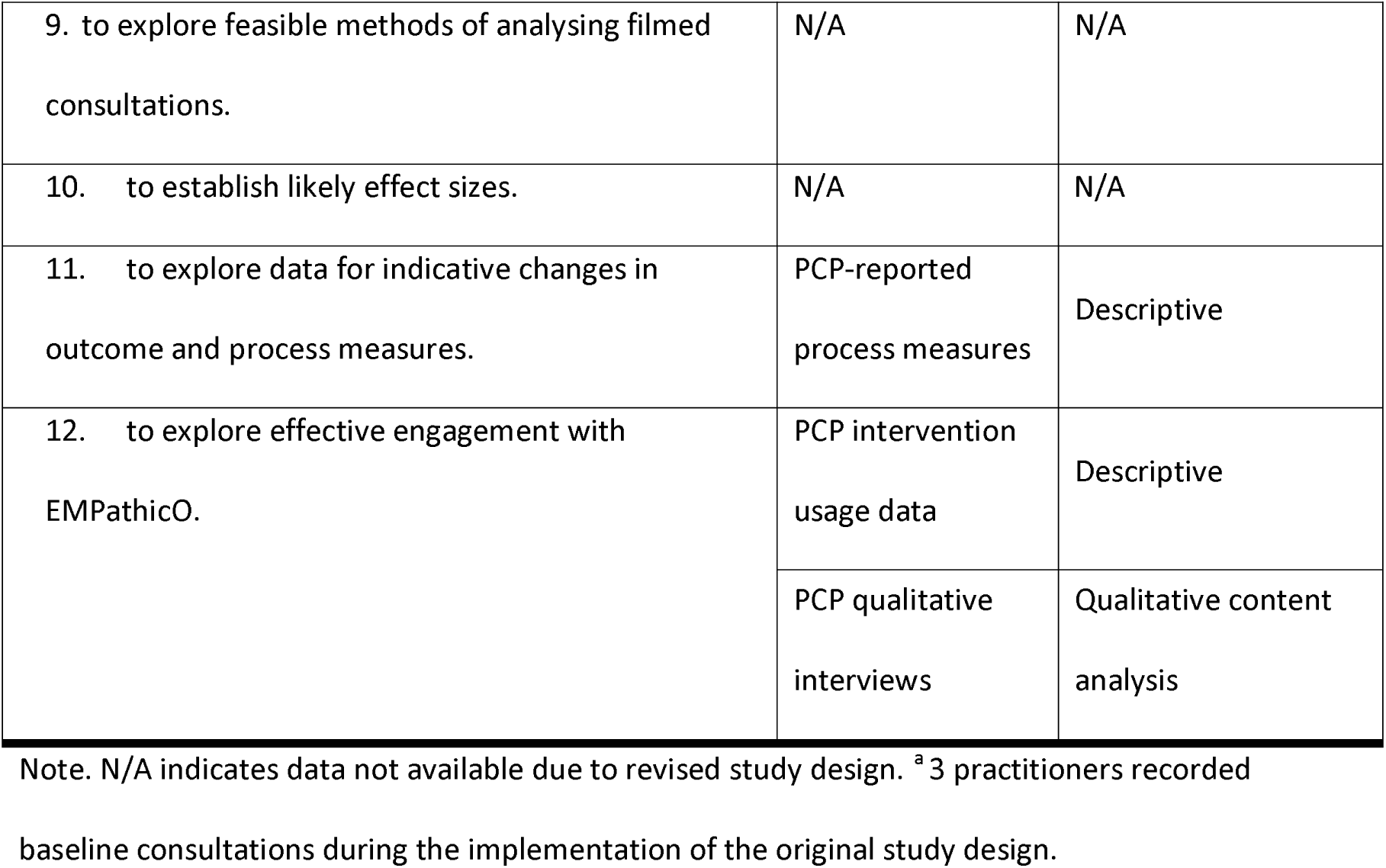
Summary of Objectives, Associated Data, and Analytic Techniques

### Patient Public Involvement (PPI)

Co-author Jennifer Bostock contributed a patient perspective to the design, conduct, and write-up of this study as a member of our trial management group. We received additional PPI input from others (see Acknowledgements) on specific aspects of this study. Our PPI partners were people with OA and/or carers for people with OA.

### Recruitment, Eligibility, and Consent: Practitioners

Practitioners were recruited from practices who had already enrolled or expressed interest in the original study design and practices known to the research team. Practitioners were offered feedback on the study, certificates, and CPD guidance; NHS support costs and research costs to cover their time for participation were paid to practices in line with recommendations from the NIHR-CRN.

Eligible practitioners reported regularly seeing people with OA in primary care in England.

All practitioners received a participant information sheet and the opportunity to ask any questions before giving informed consent via the trial website. Consent was reconfirmed verbally before commencing qualitative interviews.

### Recruitment, Eligibility, and Consent: Patients

Patients were recruited via targeted advertising on Facebook; social media posts (twitter, Instagram, Facebook); and printed posters and flyers distributed to pharmacies, retail and community settings in the Wessex region and other areas chosen to increase potential to reach people from diverse ethnic backgrounds (e.g., parts of London and the Midlands). When practices regained capacity for some research activity from July 2020, study adverts were also placed on general practice websites/social media and practices sent SMS messages to their recent consulters. There were no (financial or non-financial) incentives for survey respondents.

Eligible patients self-reported being at least 18 years old and having consulted a PCP within the previous two weeks. Using targeted advertising, we sought to include some patients who had consulted about OA symptoms and some who had consulted about other symptoms.

Study advertisements directed patients to the study website, on which they were presented with an information sheet, screening questions, and consent questions before accessing the study questionnaire on Qualtrics (Qualtrics, Provo, UT). The information sheet provided contact details of the research team for patients to ask any questions before giving informed consent online. Consent was reconfirmed verbally before commencing qualitative interviews.

### Sample Size

As per the original study design, we aimed to recruit up to 20 PCPs from 10 practices, 60 patients with OA and 120 patients with other reasons for consulting. We considered this would be sufficient to examine our objectives related to practice, practitioner and patient recruitment, patient retention, and patient reported outcome and process measures; this size is also typical of UK feasibility trials.[31] However, as the pandemic continued into summer 2020 it became clear that we would likely under-recruit PCPs and over-recruit patients, and so we sought and obtained approval from the sponsor and ethics committee accordingly.

### Outcome and Process Measures

Table 2 lists all patient-reported outcome and process measures by time-point. Patient-reported outcomes were guided by the OMERACT-OARSI core outcome domains for trials in hip and/or knee OA: pain, physical function, quality of life, patient global assessment of the target joint, and adverse events including mortality.[32] Process variables and measures were selected to assess key variables hypothesised to mediate the relationship between doing EMPathicO training and improved patient outcomes (as shown in logic model, Fig 2). Patients completed questionnaires via Qualtrics.

**Table 2.**
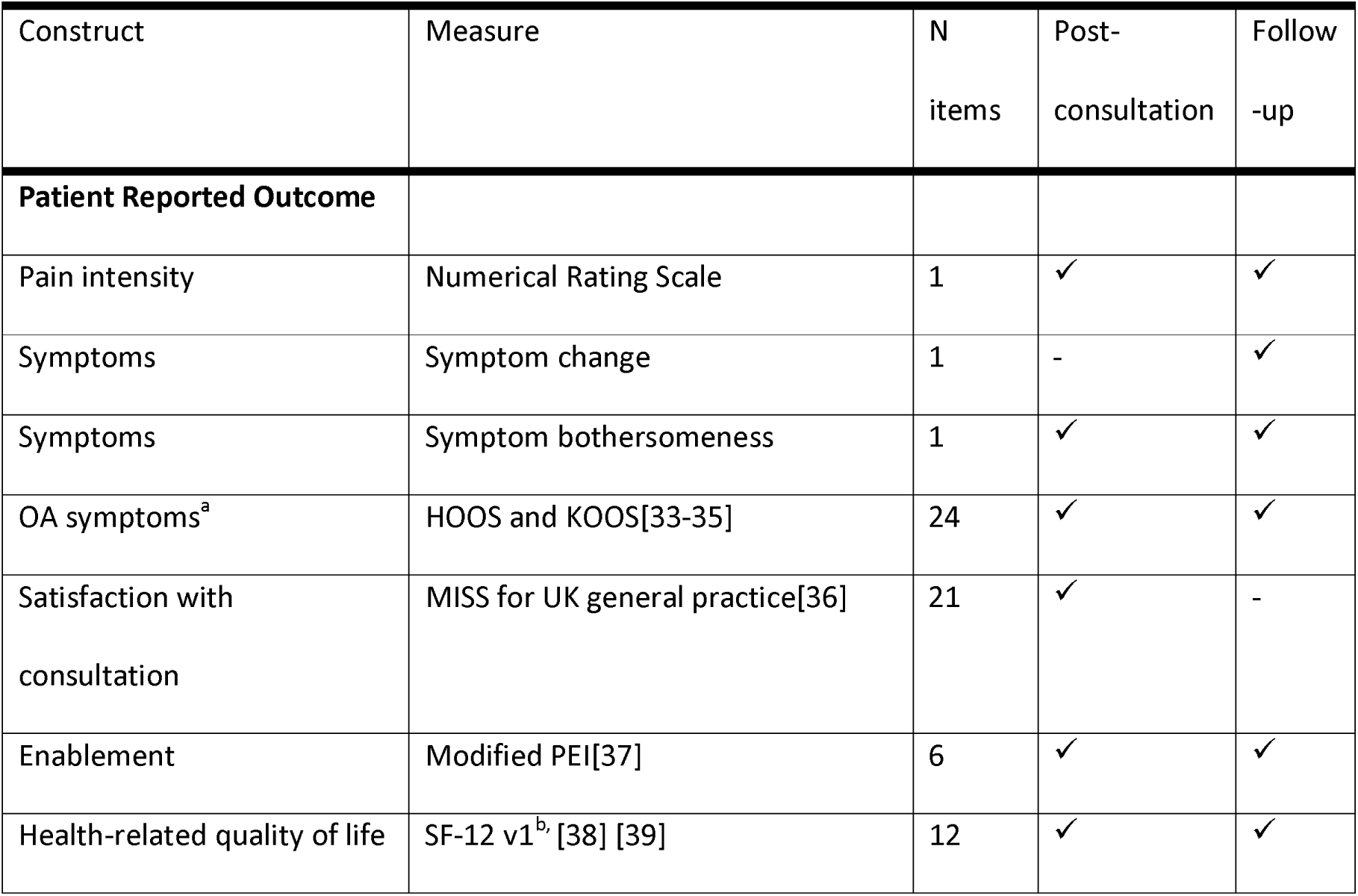
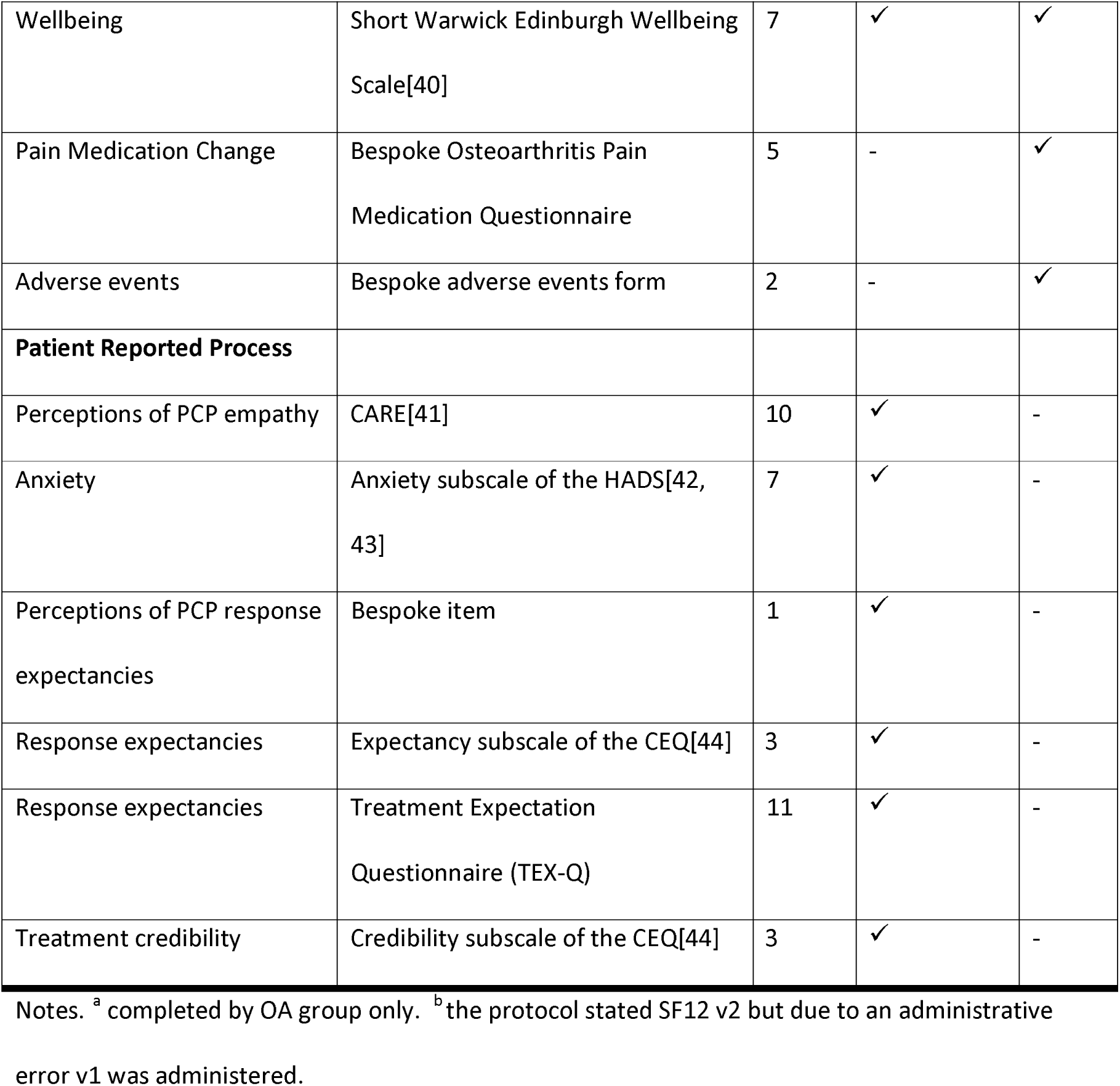
Patient Reported Outcome and Process Measures.

| Construct | Measure | N<br>items | Post-consultation | Follow<br>-up |
| --- | --- | --- | --- | --- |
| **Patient Reported Outcome** |  |  |  |  |
| Pain intensity | Numerical Rating Scale | 1 | ✓ | ✓ |
| Symptoms | Symptom change | 1 | - | ✓ |
| Symptoms | Symptom bothersomeness | 1 | ✓ | ✓ |
| OA symptoms<sup>a</sup> | HOOS and KOOS[33-35] | 24 | ✓ | ✓ |
| Satisfaction with<br>consultation | MISS for UK general practice[36] | 21 | ✓ | - |
| Enablement | Modified PEI[37] | 6 | ✓ | ✓ |
| Health-related quality of life | SF-12 v1<sup>b</sup> [38] [39] | 12 | ✓ | ✓ |
| Wellbeing | Short Warwick Edinburgh Wellbeing Scale[40] | 7 | ✓ | ✓ |
| Pain Medication Change | Bespoke Osteoarthritis Pain Medication Questionnaire | 5 | - | ✓ |
| Adverse events | Bespoke adverse events form | 2 | - | ✓ |
| **Patient Reported Process** |  |  |  |  |
| Perceptions of PCP empathy | CARE[41] | 10 | ✓ | - |
| Anxiety | Anxiety subscale of the HADS[42, 43] | 7 | ✓ | - |
| Perceptions of PCP response expectancies | Bespoke item | 1 | ✓ | - |
| Response expectancies | Expectancy subscale of the CEQ[44] | 3 | ✓ | - |
| Response expectancies | Treatment Expectation Questionnaire (TEX-Q) | 11 | ✓ | - |
| Treatment credibility | Credibility subscale of the CEQ[44] | 3 | ✓ | - |

Notes. <sup>a</sup> completed by OA group only. <sup>b</sup> the protocol stated SF12 v2 but due to an administrative error v1 was administered.

Practitioners completed questionnaires within the EMPathicO intervention on LifeGuide.

### Patient-Reported Outcomes

#### Patients Consulting for Hip-Knee OA Symptoms

The short form of the Hip and Disability Osteoarthritis Score (HOOS-12) and the Knee Injury and Osteoarthritis Score (KOOS-12) each assess pain, function, and quality of life, and produce an overall summary hip/knee impact score respectively and are scored by summing items on each scale and transforming to a 0 (extreme symptoms) to 100 (no symptoms).[33] The 12-item versions reduce patient burden and demonstrated promising psychometric properties compared to the 40-item versions in patients undergoing joint replacement surgery.[33–35] To assess patient global assessment of target joint,[32] the OA group rated their knee or hip symptoms now compared to two weeks ago. A single item 11-point numerical rating scale (pain in last week, 0=no pain; 10 = worst possible pain) was also used in the OA group, to explore whether this would be more feasible for patients to complete before a consultation, compared to the 12-item HOOS or KOOS.

A bespoke Osteoarthritis Pain Medication Questionnaire was used to assess medication change in the OA group. This instrument, adapted from the validated Medication Change Questionnaire[45] to reduce complexity for patients, asks patients to list all the osteoarthritis pain medications they are using (to include all tablets, medicines, gels, and creams) and to rate any changes in use since starting the study. The main adaptations were: removing the questions “Is there any of this medication that you would like to cut down, to take less of?” and “Is there any of this medication that you would like to take more of?” and the associated details; replacing daily dosage of all medications with a qualitative judgment on any changes in medication use (“For each of these medications, please tell us whether the amount you use has changed since you saw the doctor/nurse/physiotherapist and joined this study approximately two weeks ago. Think about how much you used this before you joined the study. Now think about how much you have use this since you joined the study, about 2 weeks ago. Have you used this medication more, less, or about the same as you did before?” Response options: Much more/A bit more/About the same/A little bit less/Much less).

#### All Patients

To explore the feasibility of two candidate primary outcomes that could apply to both OA consulters and others, we measured symptom change and symptom bothersomeness over the past two weeks. The symptom change item asks patients to rate their overall symptom change (Much better/A little better/About the same/A little worse/Much worse), and was adapted from the COOP-WONCA charts.[46, 47] The symptom bothersomeness item asks patients to rate how bothersome their symptoms are (Not at all/Slightly/Moderately/Very much/Extremely), and was adapted from the item developed to assess severity of back pain in primary care.[48] Both are single item generic symptom measures, feasible to collect from a large number of patients with diverse health conditions.

The Patient Enablement Index (PEI) captures the extent to which patients feel confident and empowered by a consultation to cope with their illness, to keep healthy and to help themselves.[37] The original publication described six items with 4 response options (much better/never/same or less/not applicable). We used a modified 7-point response scale using agree-disagree anchors to increase sensitivity to change.

Overall satisfaction with the consultation was measured using the validated, reliable, 21-item UK primary care version of the Medical Interview Satisfaction Scale (MISS-21).[36] The MISS-21 uses 7- point agree-disagree Likert scales to assess patients’ experiences of four aspects of the consultation – satisfaction with communication (4 items, e.g., “The doctor did not really understand my main reason for coming”), satisfaction with rapport (8 items, e.g., “the doctor seemed warm and friendly to me”), feeling relief from distress (6 items, e.g., “after talking with the doctor, I know just how serious my illness is”), and intending to follow advice (3 items, e.g., “It may be difficult for me to do exactly what the doctor told me to do”).

Wellbeing was assessed using the Short Warwick Edinburgh Wellbeing Scale.[40] The Warwick Edinburgh Wellbeing Scale underwent extensive development, focuses exclusively on positive aspects of wellbeing, and captures both hedonic (pleasure) and eudaimonic (self-actualisation) aspects of mental health.[49] The short version reduces participant burden and retains robust psychometric properties as a unidimensional interval level scale. [40]

Quality of life was assessed using the SF-12 v1.[39] The SF-12 has acceptable psychometric properties[38] and comprises 12 items evaluating patient-perceived impact of health concerns (physical, emotional, pain) on activities of daily living including work and social activities, calculated as physical health and mental health component scores.

An adverse events form was included at the follow-up measurement point. This form was adapted from the ACTIB trial[50] and asks whether, since starting the study, participants had any of the following events: a life threatening event, admission to hospital where you had to stay overnight, other medical events requiring medical attention. They are asked to provide details of any such events. They are then asked “Has your health been adversely affected since the start of the study?” with Yes/No response options, and space for details if ‘Yes’.

### Process Measures

#### Directly Assessed Process Measures

Intervention usage data collected in a feasibility study can suggest essential and non-essential parts of an intervention and provide insight into how it is used, potentially informing further tweaks to the intervention before final full trial. Usage data was collected via LifeGuide and included: when and for how long practitioners logged on to EMPathicO; which content was accessed (and for interactive components, engaged with) and for how long; the order in which content was accessed.

#### Practitioner-Reported Process Measures

Our logic model includes three main precursors to practitioners adopting the behaviours taught in EMPathicO: self-efficacy, outcome expectancy, and intentions for conveying empathy and optimism in consultations. We wrote items to assess these constructs by following recommendations from Bandura’s work on outcome expectancies and self-efficacy,[51] the Theory of Planned Behaviour on intentions,[52, 53] and the Health Action Process Approach on coping efficacy,[54] and combining standard item stems and response options with bespoke wording informed by qualitative interviews conducted with PCPs during EMPathicO’s development.[23] This process resulted in 12 items measuring self-efficacy for conveying empathy (7 items) and optimism (5 items) on 11-point response scales; 8 pairs of items measuring outcome expectancy and outcome value for implementing the changes selected as part of EMPathicO (scores on each pair were multiplied to give a measure of expectancy in the likelihood and value of the outcome); and 3 items measuring intentions to implement the changes selected as part of EMPathicO, on 7-point response scales (see S6 Appendix).

#### Patient-Reported Process Measures

EMPathicO aims to improve PCPs’ communication of clinical empathy and realistic optimism, and we expect that any such changes, to be clinically meaningful, should be noticed by patients. Patient perceptions of PCP clinical empathy were assessed using the 10-item CARE;[41] patients rate how well their practitioner demonstrated 10 aspects of clinical empathy in their consultation, using 5- point response scales. This is validated, reliable, questionnaire has been used extensively in UK primary care settings to assess patient perceptions of GP clinical empathy. In the absence of an existing measure, patient perceptions of practitioner response expectancies were assessed using a bespoke single item with 7 response options drafted for this study (see S6 Appendix).

Increases in practitioners’ communication of clinical empathy and realistic optimism should lead to increases in patients’ response expectancies and perceptions of treatment credibility. The Credibility Expectancy Questionnaire (CEQ)[44] was used to assess patient response expectancies and perceptions of treatment credibility. The 3-item expectancy subscale assesses the extent to which patients believe their symptoms will improve. The 3-item credibility subscale assesses the extent to which patients believe their treatment to be credible in general for their condition. The CEQ is reliable and valid and has been used across many diverse settings and patient populations, including OA and primary care.[55, 56] We also assessed patient response expectancies using a recently developed questionnaire specifically designed to assess patient expectations with respect to the outcome of medical treatments, the Treatment Expectation Questionnaire (TEX-Q).[57] This data will be reported separately.

### Qualitative Data

We invited all participating practitioners to take part in an audio-recorded semi-structured telephone interview, conducted by KS and JV and anonymised on transcription by trained supervised students. Interviews lasted between 14 and 32 minutes. The topic guide comprised open-ended questions used flexibly to explore practitioners’ experiences and perceptions of barriers and facilitators to implementing the trial and to accessing and implementing EMPathicO (see S7 Appendix).

We invited by email a purposefully varied sample of 66 patient survey respondents to take part in a telephone interview, aiming to include people with a range of age, gender, ethnicity, education level, pain condition and OA, consultation modality (telephone, face-to-face or multiple) and practitioner profession. Thirty-three people responded to invitations, 3 declined (without giving reasons) and 30 were interviewed. The topic guide, developed and piloted by the study team, comprised open-ended questions about experiences of the consultation, perceptions of practitioner empathy and optimism, and experiences of the survey (see S8 Appendix). Participants were encouraged to elaborate on their views and experiences of recent primary care consultations.

Interviews were conducted by three experienced female qualitative researchers (KS, JV, MS) and lasted on average 28 minutes (range 15 to 43 minutes). Interviews were audio-recorded, transcribed verbatim by a professional service, and anonymised using pseudonyms. Interviewees were given £20 shopping e-vouchers.

Interviewers made field notes after each interview, capturing initial impressions and reflections on the interview. Researchers also made field notes throughout the project, capturing reflections on methods and processes including for example notes from conversations with research networks, practice staff and PCPs. These were discussed at regular trial management meetings and key points captured in meeting minutes.

### Data Analysis Methods

Quantitative data were downloaded from LifeGuide and Qualtrics, cleaned, and imported into IBM SPSS version 28 (IBM Corp, Armonk, NY) for analysis. Participant characteristics, recruitment and retention rates, and patterns of intervention usage were examined using descriptive statistics. Scale scores on all outcome and process measures were computed following published guidelines, patterns of missing data were examined, and internal consistencies for new, bespoke, measures were analysed using Cronbach’s alpha. Free text responses to survey questions were categorised by meaning and described.

Qualitative interviews were transcribed verbatim, identifying details were removed and names replaced with pseudonyms. Thematic analysis was applied to the practitioner and patient interviews, to identify barriers and facilitators to recruitment and retention and to explore the relevance, feasibility and acceptability of the outcome and process measures.[58] Multiple researchers (JV, KS, CL-S, FB) were involved in the qualitative analysis to guard against idiosyncratic or overly selective coding. NVivo version 14 (Lumivero, Denver, CO) was used to facilitate coding, organise qualitative data, and maintain an audit trail of the analysis.

Table 1 maps the data and analyses to the objectives.

## Results

### Participants

Twelve practitioners from 9 primary care practices took part by working through the intervention (Table 3). Four hundred and thirty seven patients consented to the online survey and 387 (89%) answered at least one post-consent question, of whom 30 also took part in a qualitative interview (see Table 4). The majority of patients (67%, n=294) were recruited via Facebook, others were recruited via personal networks (11%, n=50), GP surgeries (9%, n=40), twitter (6%, n=25), and adverts placed with charities, community settings, pharmacies, social prescribers, Universities, and press release (<5% each).

**Table 3.**
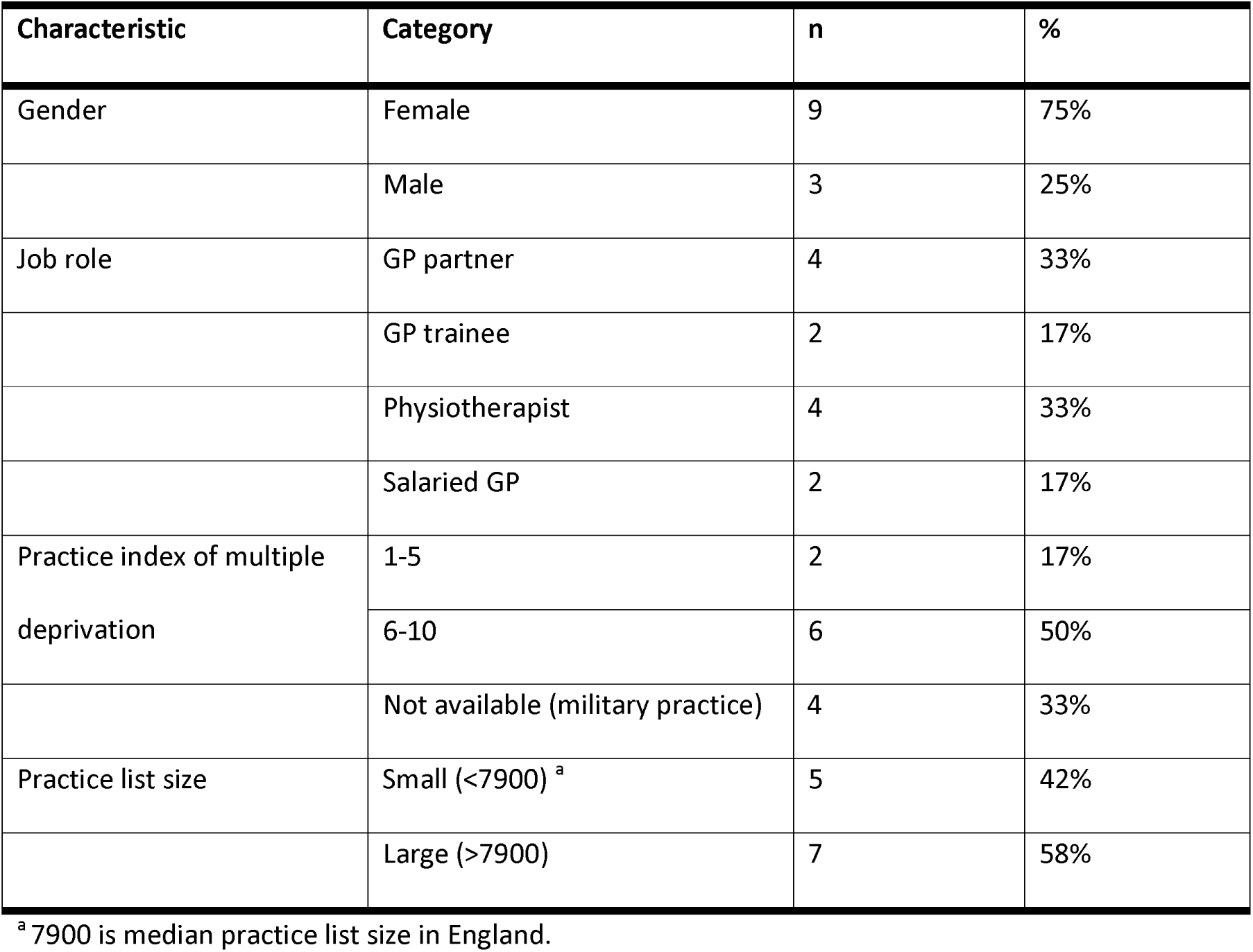
Characteristics of Practitioner Participants (n=12)

**Table 4.**
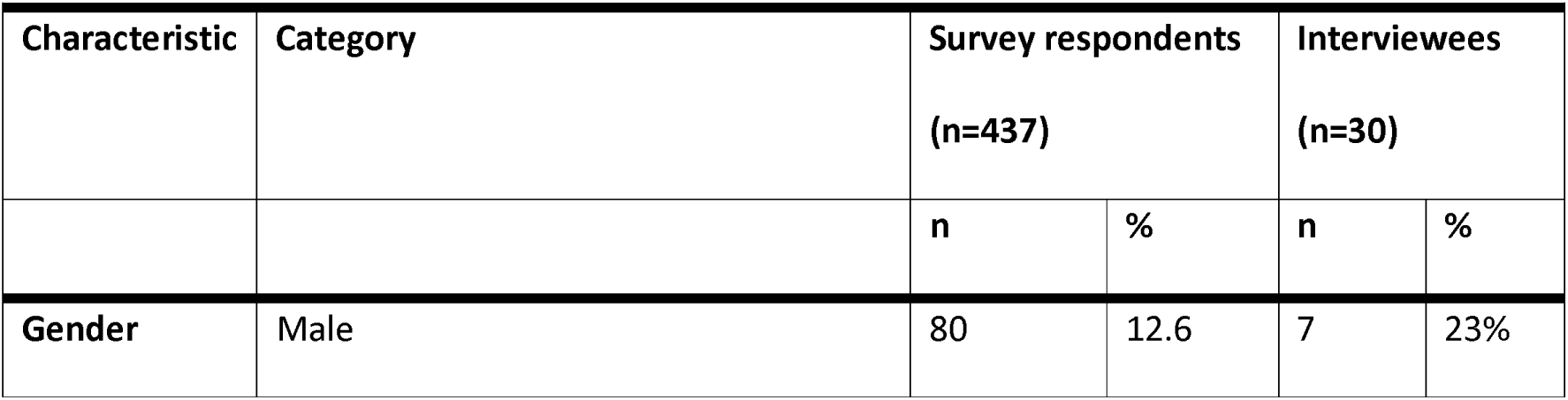
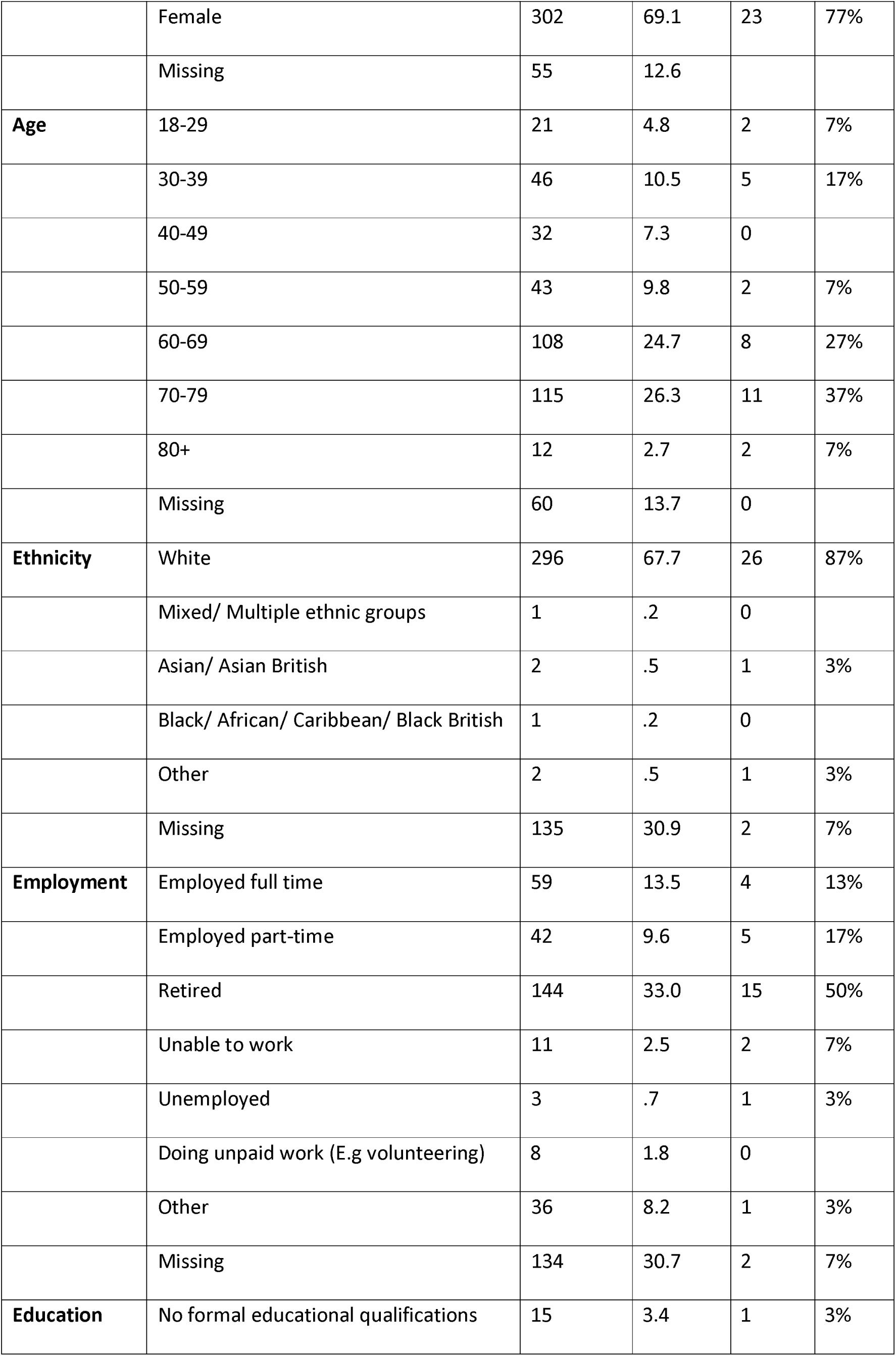
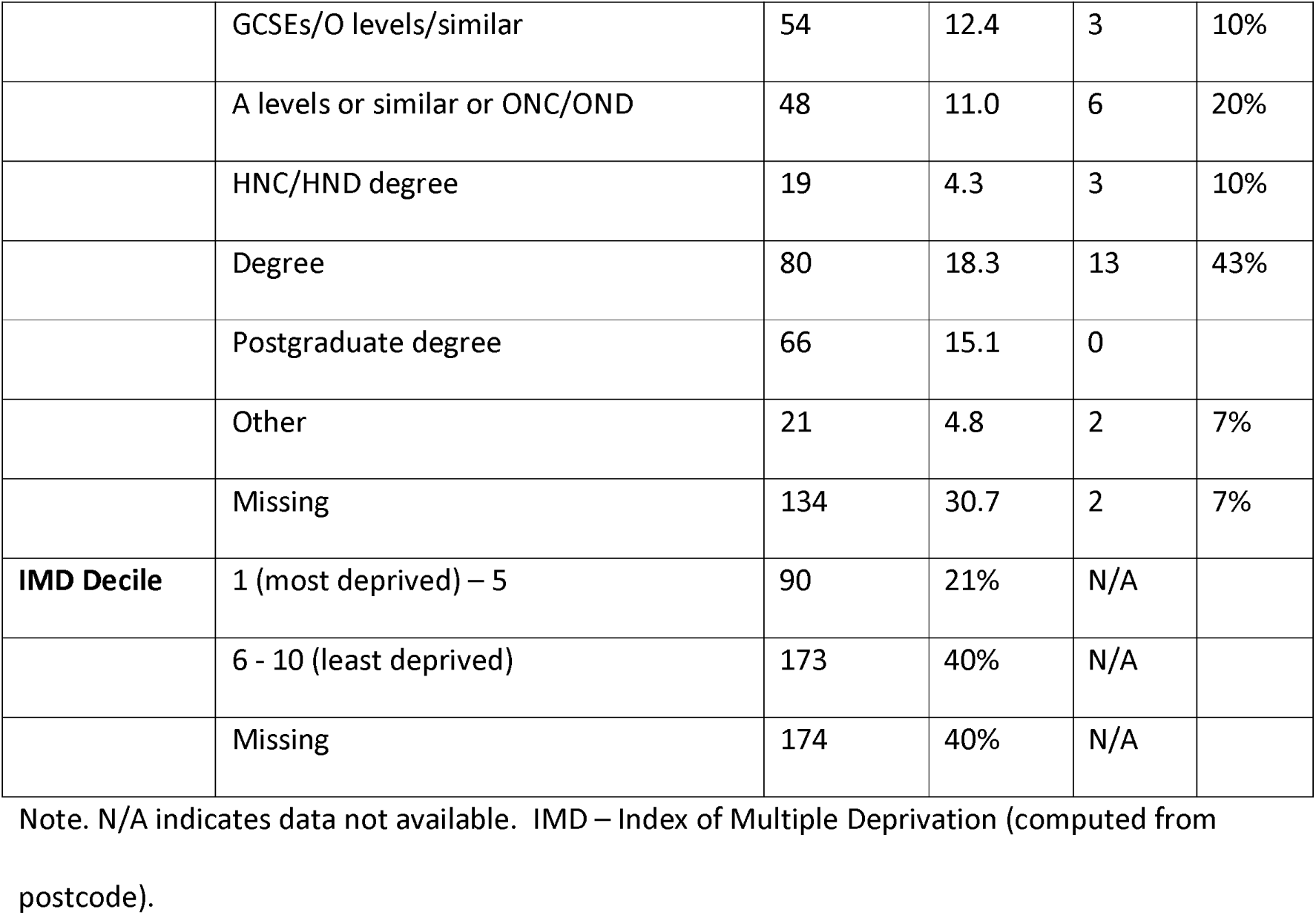
Characteristics of Patient Participants.

### Objective 1: Recruitment and Retention

#### Recruitment and Retention Rates

The flow of practices and practitioners through the study is shown in Fig 5. Twenty practices expressed interest in the study, of whom 9 (45%) went on to participate. Twelve practitioners were recruited over 6 months (January to June 2020) and 11 (92%) completed the study (one, 8% did not respond to attempts to organise the qualitative interview).

**Fig 5.**
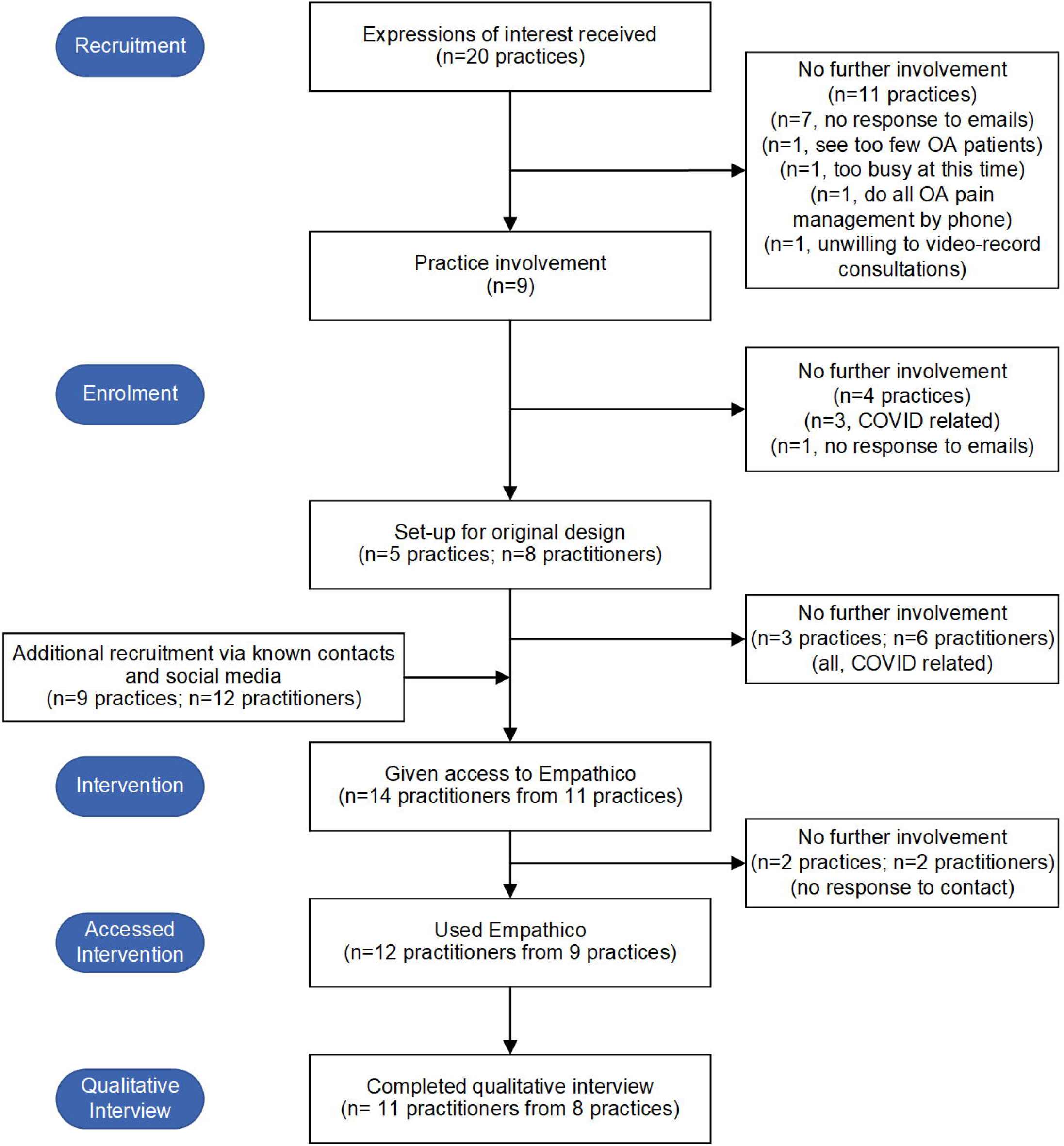
Flow of Practices and Practitioners through Study

The flow of patients through the study is shown in Fig 6. It is not possible to identify how many unique individuals visited the website, but after 1029 recorded visits, 437 eligible patients (42%) went on to complete (at least some of) the baseline survey over 5 months (May to October 2020). Most patients (91%) were consulting for reasons other than hip or knee OA. More patients completed the follow-up questionnaire in the OA group (66%) compared to the all-comers group (48%). After removing the 50 participants who consented but did not answer any of the baseline survey, the overall retention rate from baseline to follow-up was 57% (219/387). Recruitment ceased on reaching sufficient sample size for feasibility objectives, given the modified design and the ongoing pandemic context precluding further recruitment within our funding window.

**Fig 6.**
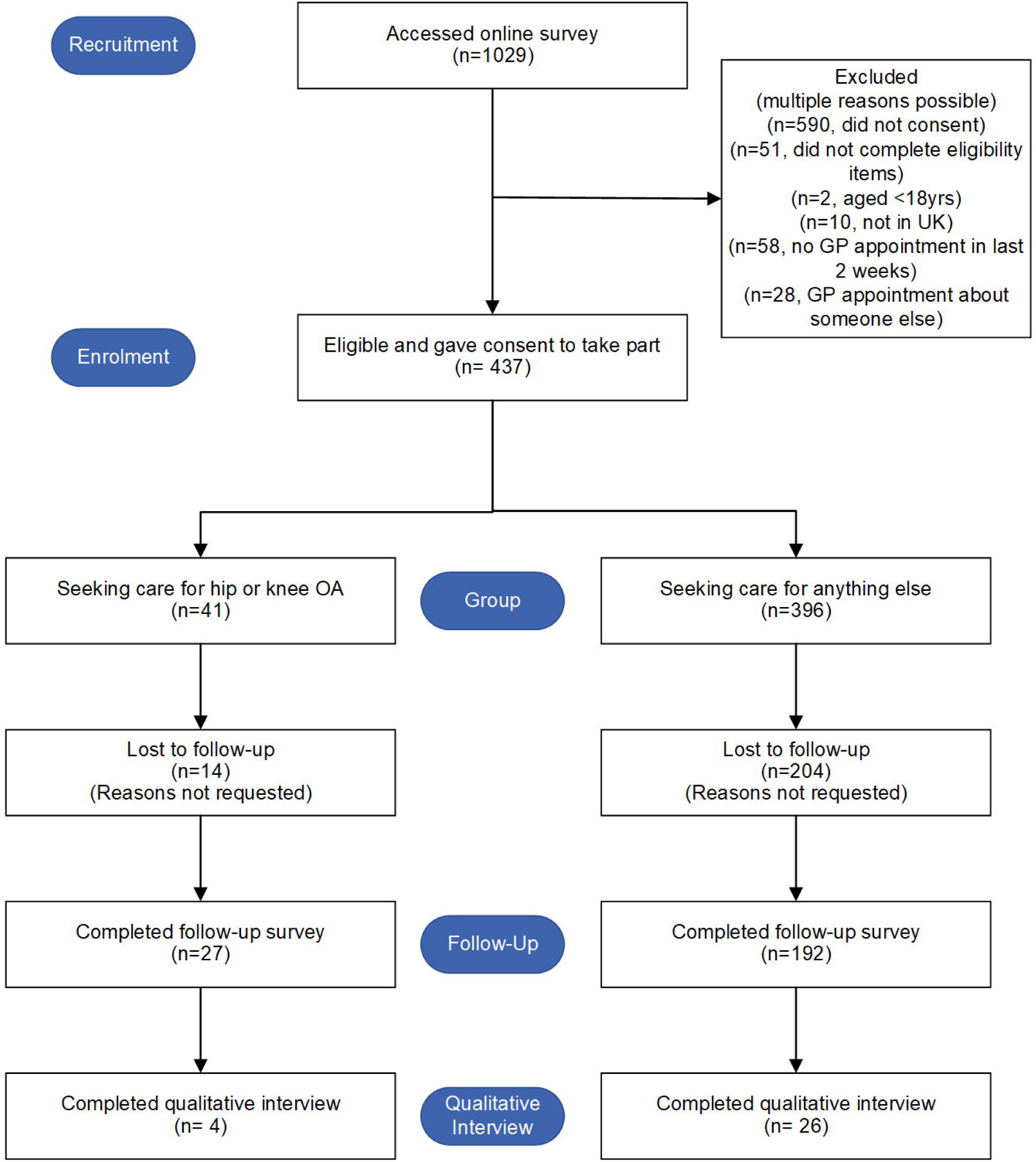
Flow of Patients through Study

#### Barriers and Enablers of Practice and Practitioner Recruitment and Retention

The team worked with four clinical research networks (each supporting research in a different geographical area) but only two successfully recruited practices. The lower service support costs (funding for practices to cover their research activity) agreed by one network may have contributed to lack of practice recruitment in their area. In the 11 practices expressing interest and subsequently declining, the main reasons were lack of research capacity and/or COVID. During the five completed site initiation visits, practices were generally enthusiastic about the study. They planned to involve mainly GPs but also nurses and physiotherapists. The three practices who withdrew after study set- up did so because of COVID.

The one practitioner who was interviewed after having experienced the original design study set up found the study appealing and feasible to conduct in practice. However, they found the multiple consent forms and paperwork complicated and recommended flow charts and “keeping it nice and streamlined and straightforward” (PCP02, GP) and avoiding the confusing discrepancy between the study name (“TIP”) and the intervention name (“EMPathicO”).

Retention of practitioner participants was very high, with 11 out of 12 completing the process measures embedded in the intervention and taking part in a qualitative interview. This may be because practitioner interviewees self-identified as having an interest in training and/or communication. They perceived that others might not prioritise this type of learning (over biomedical topics) and uptake could be lower in a larger study and/or future dissemination.

However, practitioners thought EMPathicO would be relevant to anyone who consults with patients, for example “I think it would cross lots of different clinical specialisms, from doctors to nurses, to physios, to rehabilitators like myself because ultimately it’s ensuring that within that consultation, you take as much from it as you can, but you also give the patient as much as they need from that that time” (PCP07, Physiotherapist). Nurse practitioners, advanced nurse practitioners, pharmacists and physician associates (PAs) were also mentioned as suitable training recipients. Many practitioners felt that EMPathicO would be particularly helpful for trainees or clinicians new-to- practice, but also found it useful themselves as experienced practitioners.

#### Barriers and Enablers of Patient Recruitment and Retention

Patients were recruited into the survey from targeted Facebook advertisements (n=294, 67%), personal contacts (n=50, 11%), GP websites and SMS messages (n=40, 9%), and twitter (n=25, 6%); less than 5% were recruited from the remaining sources (patient charities, community, pharmacies, social prescribers, press release, University community).

Patient interviewees described taking part because they wanted to help the researchers, to improve NHS services, to help doctors better appreciate patients’ perspectives, or to contribute to research that they perceived as being interesting or worthwhile. People who had recently had a primary care consultation when they saw the study advert perceived it to be relevant to them and were motivated to share their experiences of that consultation. This was true for people who described that consultation in broadly positive, negative, or mixed terms.

Patient interviewees expressed having had some concerns about taking part, related to data protection, governance, sharing details about a mental health consultation, the time commitment required, and not being able remember their consultation in sufficient detail. These concerns were either not strong enough to deter participation or were allayed by reassuring factors such as: feeling they understood what was involved in participating, the study being conducted by a reputable host institution (University of Southampton), having independently verified the study (by searching for and finding its webpage), understanding that research is conducted within ethical and governance standards, being able to complete study questionnaires at a time that would fit around existing commitments, and being able to withdraw from the study at any time.

Retention from baseline to follow-up was low at 57% overall. Retention was higher among patients with hip or knee OA (66%) than among all-comers (48%). Interviewees – who may have been more committed to the study than other survey respondents - reported that completing the follow-up questionnaire was “fine” or “not a problem”. Several participants reported missing the first reminder email and some suggested, for example “Another email reminder would have been helpful” (Patient01).

### Objective 2: Randomisation and Consent

#### Feasibility of Randomisation

The modified study design was implemented after only two practices had been randomised. No problems or concerns were encountered from this limited experience with cluster randomisation.

#### Feasibility of Consent

The sponsor and the Research Ethics Committee approved our original study design with a range of approaches to obtaining informed consent from patients, including: pre-consultation by a researcher in person in the surgery, pre-consultation during computerised check-in followed by post-consultation by a researcher in person in the surgery, post-consultation by a researcher in person in the surgery, by the practitioner at the start of the consultation, provisionally by the practitioner at the start of the consultation followed by a researcher in person in the surgery. Two practices expressed concern during site initiation visits about patients being consented and completing pre-consultation measures in the waiting room and planned to set aside a private space for this. In the modified study design patient and practitioner participants all completed consent online and no concerns were raised.

### Objective 3: Outcome and Process Measures

#### Filming and Analysing Consultations

One practice who expressed interest and then declined to participate reported being deterred by the requirement to video consultations as an aid to reflecting on one’s consultations during the training (see Figure 1). Three of the five practices who had a site set-up visit had their own video-recording equipment, the other two required loans from the research team.

Only one practitioner interviewee filmed their baseline consultations (before the COVID modification). They reported that patients agreed to be recorded and both parties soon forgot the camera was on, so considered there was little effect on the consultation itself. However, they also acknowledged selecting patients they knew would be open to being recorded.

GP interviewees described video-recording consultations as a powerful, educational tool that is valuable for self-reflection and self-improvement, but they associated it with their own – for some “difficult” and “painful” - experiences of recording their consultations as trainees. They considered it difficult for regular GPs to record consultations for training purposes in routine clinics due to the advanced planning and extra time required. And some would have been put off taking part had videos been mandatory in the modified study design, for example: “I may not have participated if that [videoing consultations] had been something I had to do for this [study].” (PCP01, GP), and “I think for me, it just probably wouldn’t happen. I can’t think that I would be able to easily just set that up on a day and think about how- it would just be another thing to try and do, rather than something that just fitted in easily into some learning.” (PCP05, GP) Physiotherapist interviewees did not have previous experience of recording their consultations. Overall, practitioner interviewees suggested that recording consultations should be an optional part of EMPathicO, recognising that many clinicians would not do it unless it was mandatory.

Because so few consultations were video-recorded before switching to the modified study design, there was insufficient data to develop an analytic approach based on these data alone. This aspect of the study was deprioritised and is being addressed separately.

### Relevance, Feasibility and Acceptability of Outcome and Process Measures

#### Patient Reported Measures

Fig 7 shows the flow of participants through the baseline survey. Overall, 304 out of 437 participants (70%) attempted every section of the survey that applied to them. Fifty respondents stopped completing the survey immediately after the consent pages and are excluded from further analyses. Fourteen people stopped after the MISS-21 and before the section asking about any treatments recommended during their consultation. Thirteen people stopped after the section on treatment expectations, which contained two similar measures for validation purposes (the CEQ and the TEX-Q) but may have been perceived as repetitive by participants. Very few participants dropped out in the final sections of the survey, although 12 stopped immediately before the final demographic section asking for what could be perceived as more personal, potentially identifying, characteristics including ethnicity and postcode.

**Fig 7.**
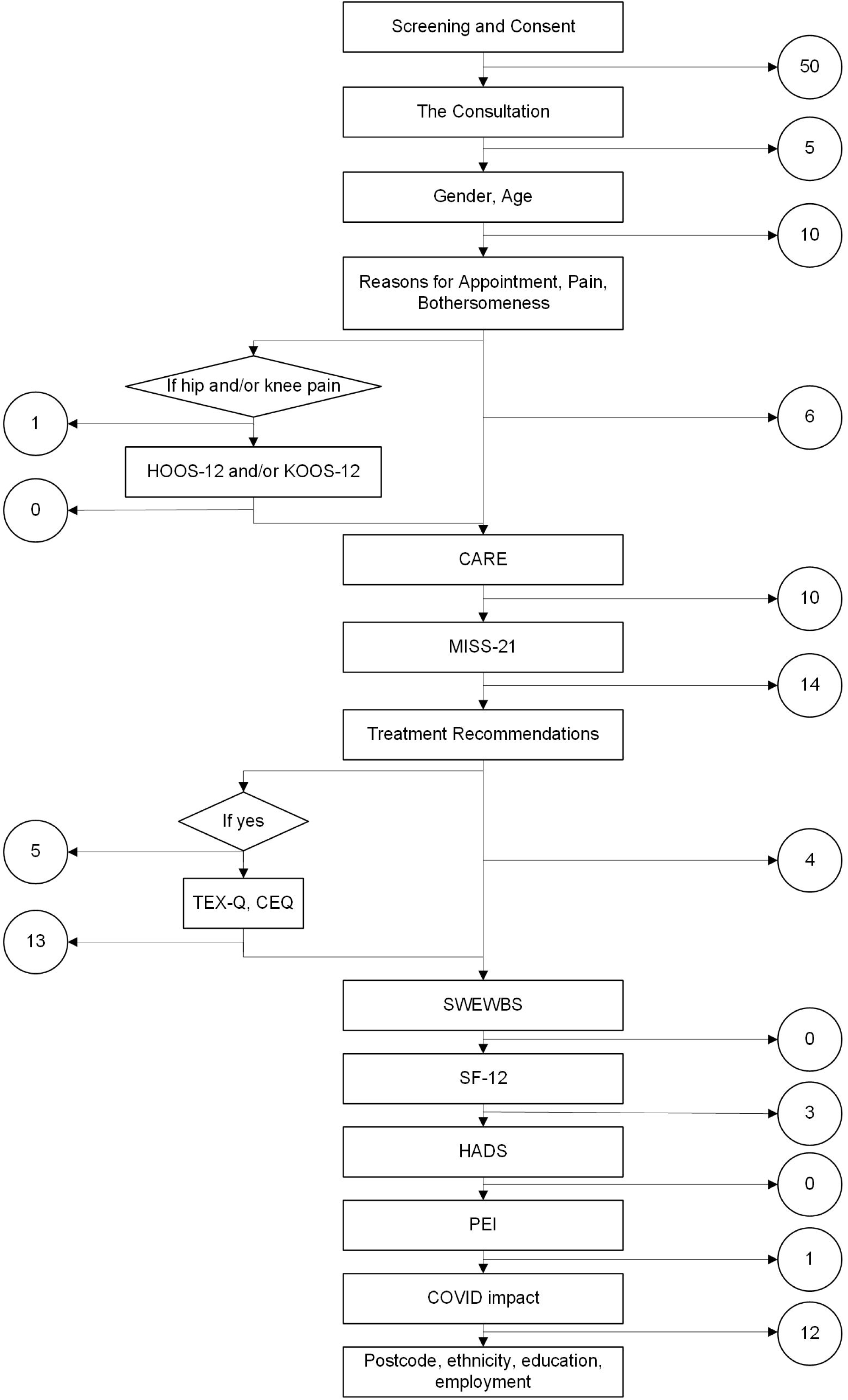
Flow of Patients through Baseline Survey Showing Numbers Withdrawing per Section

Table 5 shows the proportion of missing data points within each section of the baseline survey, among participants who were retained in the survey up to that point. Two questionnaires had notably higher rates of missing data – the MISS-21 (16%) and the CEQ (10%). On the MISS-21, four items towards the end of the questionnaire were skipped by more than 100 participants: The clinician has relieved my worries about my illness; I expect it will be easy for me to follow the clinician’s advice; It may be difficult for me to do exactly what the clinician told me to do; I’m not sure the clinician’s treatment will be worth the trouble it will take (MISS-21 items 17, 19, 20, 21). On the CEQ, the missing data was driven by 20 participants who skipped this whole questionnaire before going on to complete the next section.

**Table 5.** Missing Data by Baseline Survey Section.

| Section | Retained Participants | Items | Missing Data Points |  |
| --- | --- | --- | --- | --- |
|  | n | n | n | % |
| Consultation characteristics | 387 | 3 | 0 | 0 |
| Gender, Age | 382 | 2 | 5 | 1% |
| Reasons for appointment, pain, bothersomeness | 372 | 4 | 2 | <1% |
| Hip OA (HOOS) | 21 | 12 | 5 | 2% |
| Knee OA (KOOS) | 29 | 12 | 17 | 5% |
| Perceived empathy (CARE) | 365 | 10 | 261 | 7% |
| Satisfaction (MISS-21) | 355 | 21 | 1169 | 16% |
| Recommended treatment | 341 | 3 | 5 | 1% |
| Treatment Expectations (CEQ) | 206 | 6 | 124 | 10% |
| Wellbeing (SWEWBS) | 319 | 7 | 24 | 1% |
| Quality of Life (SF-12) | 190 | 12 | 8 | <1% |
| Anxiety and Depression (HADS) | 187 | 14 | 15 | 1% |
| Enablement (PEI) | 316 | 6 | 18 | 1% |
| COVID impact | 315 | 2 | 12 | 2% |
| Demographics | 303 | 5 | 6 | <1% |

Scores on the two bespoke 7-point items used to evaluate participants’ perceptions of clinician optimism are summarised in Table 6; the full range was used, mean scores were slightly above the scale mid-point, and missing data was minimal. Supporting the convergent validity of the perceived clinician optimism about treatment item, scores were positively correlated with scores on the established CEQ measure of patient expectancy, r(203)=.40, p<.001, and credibility of treatment r(203)=.45, p<.001.

**Table 6.** Descriptive Statistics for Bespoke Items Assessing Patient Perception of Clinician Optimism.

| Item | N | Min | Max | M | SD | Missing<br>(n) |
| --- | --- | --- | --- | --- | --- | --- |
| Thinking about your appointment, how optimistic was the clinician that your treatment will help you? | 230 | 1 | 7 | 5.40 | 1.12 | 1 |
| Thinking about your appointment, how | 103 | 1 | 7 | 4.48 | 1.21 | 2 |
| optimistic was the clinician? |  |  |  |  |  |  |

One open-ended survey question asked if participants had any problems filling in the questionnaire. Of 67 responses to this question, 26 indicated no problems; 25 suggested ‘not applicable’ options are needed and/or that some questions were difficult to answer, particularly for people who had consulted about test results or were given a referral for further investigations; 8 described problems viewing some items on a phone; and 7 provided other comments and/or clarifications (e.g., “it was a bit long”, “I was treated by a nurse not a clinician as such”).

Across all baseline survey participants, the median survey duration was 17 minutes (IQ range: 10.8 – 25.9). Most patient interviewees reported that they had found the questionnaire acceptable in terms of number of questions, time taken to answer them, clarity and ease of completion. For example, “It seemed to be a reasonable use of my time! If it had been three-quarters-of-an-hour or something I might have thought twice, but it wasn’t.” (Patient29). A few interviewees reported finding the questionnaire “a bit long” with some items that seemed repetitive, but none said the length was unacceptable with some noting that due to being in lockdown they had more spare time than usual. Interviewees reported that they had found the question content “relevant”, “clear”, “straight forward”, “simple”, and were, on the whole, willing to answer even the questions about more sensitive topics such as mental health (e.g., the HADS). For example, “I thought it was good though, it looked at it quite holistically. I know some of the questions were mental health ones, I thought that was quite interesting to not just talk about your appointment, but also how it fits in with the overall context” (Patient27).

Similar to the feedback on the survey, some interviewees whose consultation concluded with a referral, follow-up appointment, advice, or new clarity that no treatment was needed, found it difficult to answer questions that appeared to them to be about a prescribed treatment (despite some explanatory text having been provided on the questionnaire). For example, “I didn’t really have treatment and I wasn’t given medication and things so. It’s just the blood tests told us it wasn’t DVT. So then it’s just up to me to rest and not over work it and it will eventually get better. So in that way I felt a lot of it [the survey] didn’t apply to my case” (Patient9). Possible solutions suggested by interviewees including providing more ‘not applicable’ response options and more free text boxes to enable them to explain the nuance and context of their consultation. A few interviewees noted the difficulty of expressing their experiences of pain and comorbid physical and mental health conditions; one chose to answer the questionnaires in relation to her recent physical health consultation, feeling that her recent mental health consultation was “too personal”.

#### Practitioner-Reported Measures

All twelve practitioner participants completed demographic and clinical characteristics questions. All but one participant completed the new bespoke scales (practitioner self-efficacy to communicate clinical empathy and realistic optimism, practitioner intentions to achieve their individual behaviour change goals and practitioner outcome expectancies for their behaviour change goals), descriptive statistics for which are shown in Table 7. All four scales demonstrated acceptable internal consistency (Cronbach’s alphas ranged 0.69-0.98; Table 7). Practitioner interviewees reported no difficulties or concerns about the practitioner-completed measures.

**Table 7.** Descriptive Statistics for Practitioner-Completed Process Measures (n=11)

| Scale | Possible range | Actual range | M | SD | Cronbach's $\alpha$ |
| --- | --- | --- | --- | --- | --- |
| Self-Efficacy for Empathy | 1-10 | 5.0-8.5 | 6.98 | 1.12 | 0.94 |
| Self-Efficacy for Optimism | 1-10 | 5.4-9.4 | 7.05 | 1.01 | 0.88 |
| Intention to Achieve Goals | 1-7 | 2.0-7.0 | 5.94 | 1.47 | 0.98 |
| Outcome of Achieving Goals<br>(Expected Outcome x Value of<br>Outcome) | -21-+21 | 0.0-6.9 | 3.55 | 2.08 | 0.69 |

#### Effect Sizes and Indicative Changes

The modified design meant we were unable to evaluate effect sizes or indicative changes on patient reported outcome measures. Descriptive statistics for baseline patient reported outcome and process measures are presented in Table 8; mean scores should be interpreted with caution as over half of patients said that COVID had influenced their responses to the survey (51.7%, 163 of 315 respondents to that question).

**Table 8.** Descriptive Statistics at Baseline for Patient-Reported Outcome and Process Measures.

| Construct | Scale | N | N<br>Items | M | SD | se | Min-Max | Cronbach'<br>s $\alpha$ |
| --- | --- | --- | --- | --- | --- | --- | --- | --- |
| *Patient-Reported Outcomes* |  |  |  |  |  |  |  |  |
| Pain Intensity | Average pain intensity in past week | 189 | 1 | 6.03 | 2.22 | 0.16 | 1-10 | N/A |
| Symptoms | Symptom bothersomeness in past week | 372 | 1 | 3.84 | 1.23 | 0.06 | 1-6 | N/A |
| OA Symptoms | Hip Symptoms (HOOS-12) | 17 | 12 | 27.82 | 14.47 | 3.51 | 2.1-50 | 0.89 |
|  | Hip Pain (HOOS-12) | 21 | 4 | 29.46 | 16.07 | 3.51 | 0-68.8 | 0.71 |
|  | Hip Function (HOOS-12) | 17 | 4 | 30.51 | 18.47 | 4.48 | 0-62.5 | 0.89 |
|  | Hip Quality of Life (HOOS-12) | 21 | 4 | 28.3 | 16.49 | 3.60 | 0-50.0 | 0.66 |
|  | Knee Symptoms (KOOS-12) | 22 | 12 | 27.37 | 16.24 | 3.46 | 0-56.3 | 0.92 |
|  | Knee Pain (KOOS-12) | 28 | 4 | 28.57 | 17.30 | 3.27 | 0-62.5 | 0.83 |
|  | Knee Function (KOOS-12) | 22 | 4 | 28.98 | 17.73 | 3.78 | 0-68.8 | 0.87 |
|  | Knee Quality of Life (KOOS-12) | 28 | 4 | 23.44 | 17.15 | 3.24 | 0-56.3 | 0.71 |
| Satisfaction with consultation | Overall Satisfaction (MISS-21) | 164 | 21 | 4.24 | 0.71 | 0.06 | 1.9-6.1 | 0.83 |
|  | Distress Relief (MISS-21) | 196 | 6 | 4.37 | 1.31 | 0.09 | 1-7 | 0.94 |
|  | Communication Comfort (MISS-21) | 302 | 4 | 2.48 | 1.20 | 0.07 | 1-5.8 | 0.78 |
|  | Rapport (MISS-21) | 217 | 8 | 5.28 | 1.32 | 0.09 | 1.5-7 | 0.96 |
|  | Compliance Intent (MISS-21) | 226 | 3 | 3.57 | 0.66 | 0.04 | 1.7-5.3 | 0.61 |
| **Enablement** | Enablement (PEI) | 313 | 6 | 3.46 | 1.24 | 0.07 | 1-7 | 0.93 |
| **Health-related quality of life** | Quality of Life - Physical (SF-12) | 188 | 12 | 41.17 | 13.13 | 0.96 | 12.1-64.6 | N/A |
|  | Quality of Life - Mental (SF-12) | 188 | 12 | 44.74 | 11.80 | 0.86 | 8.2-44.7 | N/A |
|  | Wellbeing (Short Warwick<br>Edinburgh Wellbeing Scale) | 313 | 7 | 23.77 | 5.38 | 0.30 | 7-35 | 0.90 |
| **Process Measures** |  |  |  |  |  |  |  |  |
| **Perceived empathy and optimism** | Perceived clinician empathy (CARE) | 254 | 10 | 38.2 | 12.60 | 0.79 | 10-50 | 0.99 |
|  | Perceived clinician optimism | 333 | 1 | 5.12 | 1.23 | 0.07 | 1-7 | N/A |
|  | Treatment Expectancy (CEQ) | 204 | 3 | .05 | 7.89 | 0.55 | -17.9-10.2 | 0.94 |
|  | Treatment Credibility (CEQ) | 205 | 3 | -.01 | 6.27 | 0.44 | -16.9-7.1 | 0.86 |
| **Mental health** | Anxiety (HADS-A) | 186 | 7 | 13.44 | 4.49 | 0.33 | 0-21 | 0.87 |
|  | Depression (HADS-D) | 185 | 7 | 6.91 | 4.25 | 0.31 | 0-21 | 0.84 |

As shown in Table 7, practitioners who had worked through the intervention, scored on average above the mid-point on all four measures of practitioner self-efficacy, intentions, and outcome expectancies. This indicates that they had high levels of self-efficacy for communicating empathy and optimism, strong intentions to achieve their personal goals, and belief that achieving personal goals would have valued consequences. Qualitative data from the practitioner interviews further suggested that the practitioners were able to change some of their consultation behaviours to implement techniques from EMPathicO into their consultations. For example, “It has definitely impacted my consult. I think I’m consciously structured them differently so I am letting the patient talk. Uh, get to know what I want to know. Try to reflect back on some of those goals, and then again I make sure I finish on something positive” (PCP03, Physiotherapist). Practitioners described the training content to be easy to implement in practice and perceived that just small changes could make a difference to the consultation. Whilst participants generally found it easy to immediately implement the training techniques, there was perceived value in slightly longer term engagement to support behaviour change: “Sometimes a short period of times not long enough to ingrain the change in your style or in your behaviour, but after say three months you might have actually made the shift that you hadn’t realised” (PCP01, GP). Follow-up email prompts, a quiz, a short revision module, and the opportunity to revisit specific previously bookmarked content were suggested as ways to help practitioners embed new behaviours in practice.

#### Effective Engagement with EMPathicO

Effective engagement refers to sufficient and appropriate engagement with an intervention to facilitate the intended outcomes.[59] Because the modified design separated the practitioner and patient elements of the study, we were unable to measure patients’ ratings of clinician empathy and optimism and to relate those to training utilisation. Instead, we explored clinicians’ patterns of utilisation and experiences of the training to gain qualitative insights into engagement and potential updates or tweaks that could be made before a full trial.

Practitioners accessed EMPathicO between one and ten times (median = 3.5, total 59) and typically did so for less than an hour in total, spending longer on the content modules than on the reflection and goal setting modules (Table 9). Participants accessed the intervention mostly during working hours 09:00-18:00 (49 sessions), and all sessions took place between 05:00 and 22:00. Interviewees described the time to complete the training as appropriate and not too burdensome.

**Table 9.** Mean and standard deviation time spent by practitioners on EMPathicO (n=12)

|  | Time spent (minutes) |  |
| --- | --- | --- |
| Module | M | SD |
| Empathy | 12.08 | 14.30 |
| Optimism | 14.95 | 11.77 |
| Osteoarthritis | 17.93 | 7.61 |
| Reflection | 7.93 | 5.94 |
| Goal Setting | 3.94 | 3.09 |
| Total | 56.84 | 24.82 |

Practitioners described EMPathicO as clear, user-friendly, and relevant. For example: “it was easy to access, it was user friendly, it seemed quite straightforward and following the instructions through the course, it seems very targeted, it was brief enough not to become too onerous. I think some of these online learning tools can become quite burdensome in the time that they take, and they don’t allow you to read through at the speed that you want to. So it was brief and to the point, but had enough information to allow you to grasp what the aims of it were and for you to sort of personalise it to your own experience.” (PCP09, GP).

The content was described as straight forward and not heavy going, but well-referenced enough with good links to further evidence if desired. Practitioners liked the modular structure of EMPathicO that could be completed all at one session, or separately over different days, and found it easy to navigate with a good mixture of learning resources including text, video, and reflection. Some found it helpful to leave time between modules to reflect on and try to implement the ideas suggested.

The empathy module was generally described as a helpful refresher of fundamental points that clinicians felt that they already knew, and this was the case for very experienced and more recently trained practitioners, for GPs and physiotherapists. For example, “some of that, well I suppose I felt was maybe a bit basic, but it’s- it’s a recap isn’t it? So, it’s not wasted learning” (PCP10, GP). Most thought it was helpful to bring together multiple aspects of empathy in one place in a way that facilitated explicit, focused, reflection on their current practice. One physiotherapist had not thought about the use of empathy as a treatment and found the ideas more novel. Practitioners found it helpful to be reminded of the importance of non-verbal communication for communicating empathy and some reflected on times when their non-verbal communication may have diverged from their verbal communication.

The optimism module was generally seen as relevant, novel, implementable and thought-provoking: “Novel and it was made to be very achievable, and probably is something we don’t do normally” (PCP02, GP). It felt relevant to practitioners who, even if they saw themselves as generally optimistic people, described how this may not be consistently communicated to patients in practice especially towards the end of a long day or week: “So by Friday we kind of had that drain on your mood all week and sometimes you can find when you’re with a patient that is particularly negative, you know it hits you at the right time and you can see that spiralling yourself, with the way you’re kind of going, well, maybe you’re right, there is no hope to your knee pain or whatever it might be.” (PCP04, Physiotherapist). Aspects that were highlighted by practitioners as being particularly novel and implementable were conceptualising optimism as a form of treatment, using optimism in “difficult consultations,” using optimism later in the consultation but not at the start, specific phrases suggested in EMPathicO to convey optimism, and reframing safety netting in a more positive way.

Practitioners reflected on the complexities of optimistic communication in the context of a weak evidence base, in situations that seemed to require supporting patients to accept the status quo, and when patients hold what the practitioner considers to be unrealistic expectations. Content about the need to tailor optimism to individuals was well-received: “So particularly the parts that spoke about being specific and being realistic, I thought were kind of helpful, rather than just a broad statement about trying to be optimistic. So yeah, sort of tailoring it through the consultation was useful” (PCP05, GP).

The OA module was described as straightforward; practitioners were positive about the benefits of empathy and optimism in OA and felt the module would help them to enhance consultations accordingly. Participants suggested adding content on how to deal with patients who struggle to be optimistic, and those who consider that they have already adhered to recommended exercises or medication without the desired outcome. GPs found the video of an OA GP consultation somewhat idealistic in that the patient consulted primarily for knee pain, accepted the given treatment options, and had no competing health needs. Physiotherapists found the video overly simplistic: “If I look at it with my biases of physio having 45 - 60 minutes for the new patient, it does take a reductionist approach in terms of like here’s a couple of exercises, you’ll be fine. Things can be more complicated than that I would suggest. But in a 15-minute consult I thought it was a really good way to show the elements of empathy, positive language and then the kind of the warming up” (PCP04, Physiotherapist).

When engaging with the reflection module, most participants described using the EMPathicO checklist to guide them. Participants often reflected on a consultation that they had conducted in the previous couple of days to ensure they could recall things clearly. Some also took a broader approach, considering what they do in more general terms, but found this to be less specific or useful. Most found the reflection and goal-setting modules to be useful and were able to set themselves some goals to change practice, for example: “I did find that reflection helpful, it’s jarred me into thinking about it, which I hadn’t ever done before“ (PCP01, GP).

Fig 8 shows the goals set by practitioner interviewees. Goals about optimism were more commonly set than goals about empathy, consistent with practitioners finding the optimism module novel and implementable within routine consultations. The two most commonly set goals associated with the optimism module were positive safety-netting and positive language.

**Fig 8.**
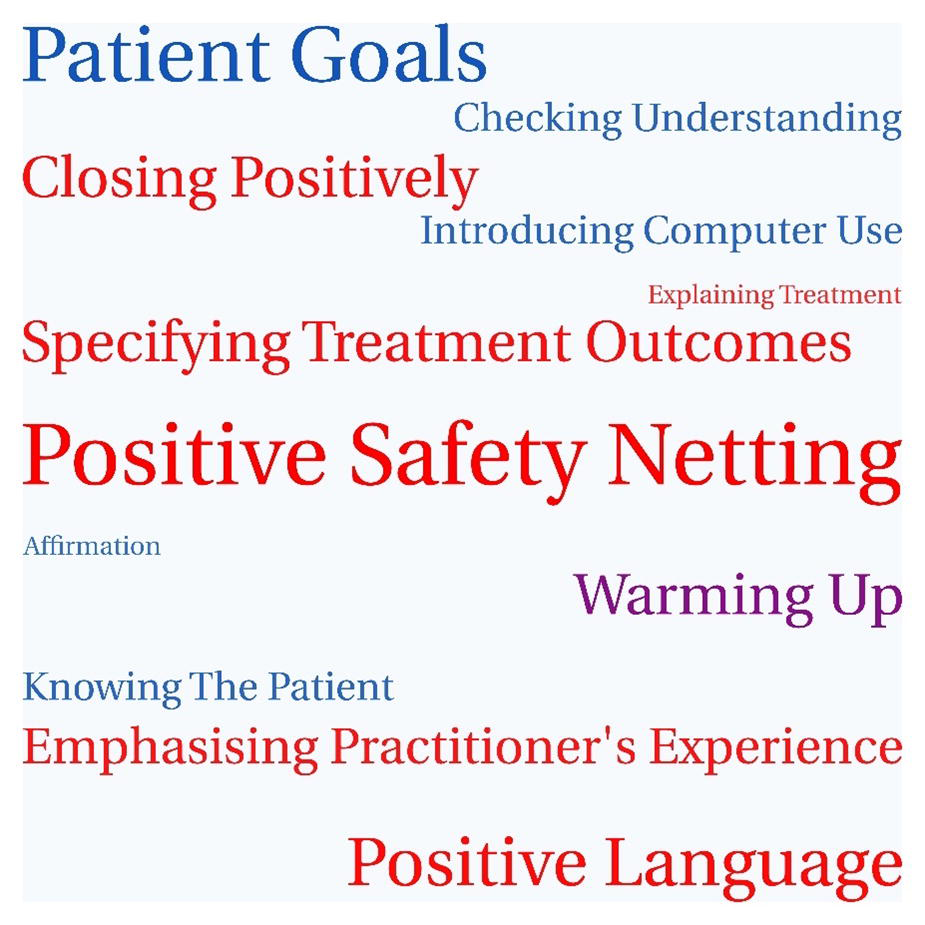
Goals set by practitioner interviewees.

*Note. Goals in red are associated with the optimism module; goals in blue are associated with the empathy module; goals in purple are associated with both modules. The size of each goal represents the number of practitioners who chose it*.

Positive safety netting was considered easy to tweak and simple to implement, and potentially beneficial for patients. Many participants reflected that their language in consultations was not positive enough and found it helpful to see the examples of how to avoid negative terms, to frame treatments more positively and to close consultations with a positive summary. For example:

“I think the thing about negative phrases, that’s probably the biggest thing I’ve taken from this, is trying to think about how I phrase things and rather than saying ‘well, it works for 50% of people but not the others.’ Thinking about actually ‘this works for over half of people, lots of patients find it very successful, lots of patients get some benefit from it’. And trying to think of it in a much more positive way. So, I’m not trying to mislead people but, thinking about it in and trying to just- just remove that negativity I suppose. Which I suppose sometimes just creeps in without us really maybe realizing it” (PCP10, GP).

The most commonly set goal associated with the empathy module was to ask about and refer back to patients’ own goals. Participants reported that they might establish how the condition affect the patient but not how it affects what patients want to achieve. Most reported that they didn’t commonly make treatment and management plans linked to the patient’s own goals and thought this could be done quite easily. Physiotherapists reported being familiar with goal-setting in general but less familiar with a collaborative approach to goal-setting or a more open approach to patients’ own goals. For example, “I’ve done quite a bit around goal-setting before. But again, I think it’s just useful just to make sure that when we set goals with our patients, they are collaborative, and we’re not necessarily leading the patient on a goal that we think is best for them, and making sure that they are part of that, that process as well” (PCP07, Physiotherapist).

## Discussion

During 2020, we completed a feasibility study of methods for evaluating EMPathicO in a cluster- randomized controlled trial in primary care. Despite the first COVID-19 lockdown in England occurring a few weeks after commencing the trial, agile modifications meant it was still possible to collect and analyse relevant data. Before modifications, we had secured sponsorship and ethics approval, established a recruitment pipeline for practices including first contact physiotherapists and successfully set-up 5 practices; we also identified barriers to practice recruitment. After the first lockdown, when practices had no capacity to recruit patients, we modified the study design to looked separately at practitioner activities (intervention, practitioner reported measures) and patient activities (patient reported measures). While moving patient recruitment outside of practices meant it was not possible to test planned practice-based recruitment methods, to explore recruitment in practices serving a diverse range of communities, to collect data on harms for patients, or to collect data to inform estimated effect sizes, these issues can be informed by prior literature and other study objectives were addressed to varying degrees.

Working with the clinical research networks was an effective and efficient way to recruit practices and practitioners, although variability in service support costs offered across different networks may result in geographical variation in practice recruitment. Barriers for practice participation included seeing insufficient patients with OA, not wanting to film consultations, using telephone consultations for pain management, and being too busy during the winter months with seasonal increases in consultation rates (primary care has higher winter increases in consultation rates than secondary care [60]). Strategies that might overcome these include recruiting and working closely with practices who have first contact practitioners to whom OA patients are triaged; removing the requirement to film consultations, thus removing a barrier to practitioner participation and the inclusion of telephone consultations; not starting the trial in January/February and planning for seasonable fluctuations in patient recruitment. Practitioners who enrolled in the trial were enthusiastic and committed to the study and suggested it would be relevant to newly qualified and experienced PCPs from diverse professional backgrounds. They also warned that some PCPs would likely prioritise training in more biomedical topics than in communication skills. Simplifying practitioner-facing study documents and strengthening the description of the training to appeal to a broad audience could help.

Targeted Facebook advertisements was the most effective way of recruiting patients via social media, but retention into the follow-up survey was suboptimal particularly among those who had consulted for reasons other than OA. Patients were enthusiastic about research on practitioner- patient communication and keen to help. Retention could be improved by emphasising study benefits; timely reminders; offering postal and/or telephone formats in addition to web-based; giving incentives not conditional on completion; and working flexibly e.g., with PPI input, to engage participants in the study. [61–64] [65] Because most patients in this feasibility study were recruited via social media they may differ from patients in the main trial who will be recruited via primary care practices. Being invited to take part by one’s own GP practice may enhance recruitment and retention rates compared to social media recruitment, by making the study more personally relevant and credible, although patient recruitment remains challenging across trials.[66] There was extensive missing data on patient ethnicity and the sample was likely lacking in diversity; this needs to be addressed in the full trial.

The modified design left limited scope to address our second aim and associated objectives, about randomisation procedures, consent procedures and eligibility criteria. Practice randomisation was approved by the research ethics committee, worked for those practices who reached that stage, and was not mentioned as deterring participation by practices or practitioners. Multiple approaches to informed consent in the original study design were approved by the research ethics committee; taking consent online (and reconfirming verbally before any qualitative interview) was acceptable and effective in the modified study design. Practitioner interviewees supported including a range of PCPs from diverse professional groups in our work.

Findings from qualitative and quantitative analysis of practitioner and patient data can inform the selection of outcome and process measures in a future full trial. While our plan was approved by the research ethics committee to have PCPs video-record consultations (both as part of the trial and as part of the EMPathicO training), practices and practitioners were not enthusiastic about doing this and some said they would not participate if videoing consultations was compulsory. This is concerning because studies that require videoing consultations may recruit biased samples of practitioners and patients.[67] Because of this, and the challenges experienced running this trial during 2020 (while members of the team also, among other things, worked clinically, cared for children when schools were closed, and delivered teaching online), we deprioritised exploring methods of analysing filmed consultations.

Bespoke questionnaire items designed for this study to measure practitioners’ self-efficacy, intentions, and outcome expectancies related to empathy and optimism, and patients’ perceptions of practitioner optimism, were internally consistent, were acceptable to participants, and the patient-reported measures correlated with measures of conceptually related constructs. Patient interviewees and survey respondents who provided qualitative feedback were generally positive about the content, length, and relevance of the survey (for the individual and/or the researchers) but did not like multiple very similar questions, although this did not put them off answering them. The main difficulty participants described was being unsure how to answer questions when their own consultation experience did not appear to fit. Consistent with this, individual survey items with higher rates of missing data tended to be those that assumed a consultation had been about a specific symptom, condition, or illness and/or were more readily applicable if a consultation had resulted in a prescription or referral for treatment. Whereas some participants had consulted about multiple issues or to discuss test results or similar, and some did not expect and/or did not receive a clear recommendation or prescription at the end of their consultation (e.g., they were sent for tests or investigations). More careful testing and work with PPI is needed to ensure the survey makes sense for patients who have consulted for multiple or non-symptomatic reasons (e.g., test results) and/or who have not received a prescription or other treatment recommendation as an outcome of the consultation. Survey flow and choice of questionnaire tools seem to be particularly challenging when trying to conduct pragmatic applied research in primary care with representative samples of all patients who consult PCPs, rather than restricting eligibility to sub-groups defined by diagnosis and/or consultation outcome.

It was not possible to establish likely effect sizes or to explore data for indicative changes in outcome and process measures. However, there were clear indications in the PCP data that engaging with EMPathicO led to increased communication of optimism and/or empathy during consultations, either through increased use of familiar techniques or uptake of novel techniques. EMPathicO was generally well-received and seen as relevant, brief, and engaging; the empathy module was typically seen as a valuable reminder while the optimism module was seen as comparatively novel; the OA module was helpful in providing examples of techniques in one specific context; and PCPs engaged well with the reflection and goal-setting modules and reported already implementing some changes to their consultations. These findings have contributed to the process of finalising EMPathicO ready for full trial.

## Conclusion

PCPs were keen to reflect on and further improve their communication skills and were prepared to undertake brief online training (even during the pressures of COVID). They found EMPathicO accessible, sufficiently brief, relevant and engaging and felt they were able to learn and implement the techniques for communicating clinical empathy and realistic optimism. They began implementing techniques immediately after completing training and found the techniques relevant to patients consulting for painful and non-painful conditions. Patients found the planned outcome and process measures acceptable and were willing to complete them online; minor problems with funnelling were identified and remedied; and patients were keen to take part in research that could improve primary care consultations. With some relatively minor changes (e.g., tweaks to EMPathicO, removing the requirement for videoing consultations, and incentivising patient questionnaires) it is feasible to proceed to a full trial of the effects of EMPathicO on patient outcomes in primary care. This is important because it would be the largest trial of an empathy and optimism intervention to be undertaken in primary care, capturing patient health outcomes. If proven effective in such a trial, EMPathicO could be rolled out at scale to enhance PCP communication skills and improve patient outcomes. Broader learning for other trials include the value of extensive testing to design questionnaire wording and structures to ensure primary care patients attending for diverse reasons and with diverse consultation outcomes are shown questions that are relevant to their situation, and the distinct lack of enthusiasm for video-taping consultations even among PCPs who are keen to enhance their communication skills.

## Acknowledgements

We thank the patients and primary care practitioners who took part in this project and all those who facilitated patient recruitment. We thank Jessima Hunter for sharing thoughts from a patient’s perspective. University of Southampton students assisted with transcribing qualitative data and/or testing questionnaires, including Alice Holden and Niamh Wood. The quantitative data for this paper was collected using Qualtrics software. Copyright © Qualtrics. Qualtrics and all other Qualtrics product or service names are registered trademarks or trademarks of Qualtrics, Provo, UT, USA. https://www.qualtrics.com.

## Data Availability

Data cannot be shared publicly because consent was obtained to share data for secondary research with suitably qualified individuals. Data are available from the University of Southampton Institutional Data Repository (https://doi.org/10.5258/SOTON/D3263) for researchers who meet the criteria for access to confidential data.

## Supplementary Material Captions

S1 TIDieR checklist

S2 Consort checklist – extension for pilot and feasibility trials S3 Consort checklist - conserve extension

S4 Appendix: Planned Methods for Original Study Design S5. Trial Protocol

S6. Appendix: Bespoke Questionnaire Items

S7. Appendix: Topic Guide for Practitioner Interviews S8. Appendix: Topic Guide for Patient Interviews

