## Supplementary material for "Feasibility trial of a new digital training package to enhance primary care practitioners’ communication of clinical empathy and realistic optimism": S1

TIDieR

### EMPathicO (Communicating clinical EMPathy and realistic Optimism in primary care consultations)

**Why:**

**Rationale for elements essential to the intervention to change practitioner behaviour**

1. Evidence-based information about the consequences of communicating clinical empathy and realistic optimism and verbal persuasion that communication skills can faciltate efficient consultations, may enhance primary care practitioners' expectancies that it is feasible to communicate clinical empathy and realistic optimism in primary care consultations and that doing so can enhance patient outcomes.
2. Acknowledging barriers to enacting communication skills, and supporting practitioners' problem-solving may enhance practitioner self-efficacy to communicate clinical empathy and realistic optimism in consultations.
3. Providing instruction and (video) demonstration of how to communicate clinical empathy and realistic optimism in consultations,and user stories, may enhance practitioner skills in communicating clinical empathy and realistic optimism in consultations.
4. Enabling practitioners to monitor their communication behaviour and supporting them to reflect on their communication behaviours, to set goals and action plan changing their communication behaviours may strengthen practitioner behavioural intentions to communicate clinical empathy and realistic optimism in consultations.
5. Enhancing practitioners' expectancies, self-efficacy, skills, and intentions about communicating clinical empathy and realistic optimism may enhance their actual communication of clinical empathy and realistic optimism in consultations.

**Rationale for practitioner behaviour changing patient outcomes**

1. Enhancing practitioners' expressions of clinical empathy may increase patients' perceptions of empathy and decrease patient anxiety.
2. Enhancing practitioners' expressions of realistic empathy may increase patients' perceptions of practitioner optimism and decrease patient anxiety.
3. Increasing patients' perceptions of practitioner optimism may increase patient treatmetn outcome expectancies.
4. Increased perceptions of empathy, increased treatment outcome expectancies, and decreased anxiety, may increase patient enablement, satisfaction with the consultation, and health-related quality of life.

5. Increased perceptions of empathy, increased treatment outcome expectancies, and decreased anxiety, may decrease patient pain intensity, pain interference, and symptom severity.

**What (material):** This digital training intervention includes text, images, and film. The materials are structured into the following sections:

- Introduction (gives an overview of the content and duration of the intervention, includes a short quiz about optimism and empathy, information on the team behind the training);
- Empathy (presents latest research on aspects empathy including personalisation, validation, and increasing expressions of empathy as the consultation progresses; includes multimedia activities);
- Optimism (presents latest reserach on outcome expecancies; includes multimedia activities);
- Osteoarthritis (provides specific examples of communicating clinical empathy and realistic optimism in consultations about OA; links to NICE guidelines for OA);
- Reflections (guidance on how to reflect on recent consultations and identify opportunities for improving the communication of clinical empathy and realistic optimism);
- Goal-Setting (activities to set goals and make plans based on previous sections);
- Menu (gives access to all prior content);
- Goal Review (presents user's personal goals back to them, prompts reflection on progress and invites revision/extension to goals).

**What (procedures):** The digital training intervention presents the Introduction section first, after which users can choose the order in which to review the empathy, optimism, and osteoarthritis sections. After reviewing these sections, the user works through the reflections section then the goal-setting section, and then is able to access all prior sections from the menu. The goal review section is made available 4 weeks after the goal-setting section has been completed, and users are prompted about this via email. When working through the digital training intervention, users are encouraged to engage in guided reflection, goal-setting, and action planning.

**Who provided:** The intervention was provided remotely in the form of self-directed e-learning. As such, intervention recipients had no interaction with any 'providers' as such.

**How (mode of delivery; individual or group):** Intervention was delivered online via a website. Intervention recipients accessed it individually. There was no interaction with other recipients as part of the intervention.

**Where:** Intervention recipients accessed it using an internet connected device (PC recommended) in a place of their choosing, e.g. at work, at home.

**When and how much:** Intervention recipients accessed it at a time of their choosing. The intervention was intended to take up to approximately 75 minutes to complete. It could be done all at once or in multiple sittings (user's choice).

**Tailoring:** No tailoring, although as described above recipients could choose the order in which they accessed some intervention sections.

**Modification:** No modifications were made to the intervention during this study.

**How well (planned):** There is no straight forward way to assess fidelity of this self-directed e-learning intervention. Usage data were captured to give an indication of number of times and length of time people logged on to the intervention, and time spent logged on to each section.

**How well (actual):** Practitioners accessed EMPathicO between one and ten times (median = 3.5, total 59) and typically did so for less than an hour in total, spending longer on the content modules than on the reflection and goal setting modules. Participants accessed the intervention mostly during working hours 09:00-18:00 (49 sessions), and all sessions took place between 05:00 and 22:00.
