## Supplementary material for "Feasibility trial of a new digital training package to enhance primary care practitioners’ communication of clinical empathy and realistic optimism": S3

| CONSERVE-CONSORT Extension | | | | | |
| --- | --- | --- | --- | --- | --- |
| Item | Item Title | Description | | | Page No. |
| I. | Extenuating Circumstances | Describe the circumstances and how they constitute extenuating circumstances. | | | 7 |
| II. | Important Modifications | a. Describe how the modifications are important modifications. | | | 7-8 |
|  |  | b. Describe the impacts and mitigating strategies, including their rationale and implications for the trial. | | | (see below) |
|  |  | c. Provide a modification timeline. | | | 8 |
| III. | Responsible Parties | State who planned, reviewed and approved the modifications. | | | 6 |
| IV. | Interim data | If modifications were informed by trial data, describe how the interim data were used, including whether they were examined by study group, and whether the individuals reviewing the data were blinded to the treatment allocation. | | | N/A |
| CONSORT Number and Item | | For each row, if important modifications occurred check “direct impact” and/or “mitigating strategy” and describe the changes in the trial manuscript or supplement. Check “no change” for items that are unaffected in the extenuating circumstance. | | | Page No. |
|  |  | No Change | Impact* | Mitigating Strategy** |  |
| 1 | Title and abstract | X |  |  |  |
| 2 | Introduction | X |  |  |  |
| 3 | Methods: Trial Design |  |  | X | 7-8, Fig4 |
| 4 | Methods: Participants |  |  | X | 7 |
| 5 | Methods: Interventions | x |  |  |  |
| 6 | Methods: Outcomes | x |  |  |  |
| 7 | Methods: Sample Size |  |  | X | 12 |
| 8-10 | Methods: Randomisation |  | x |  | 7 |
| 11 | Methods: Blinding |  | x |  | 7 |
| 12 | Methods: Statistical methods |  | x |  | 7-8 |
| 13 | Results: Participant flow |  |  | X |  |
| 14 | Results: Recruitment |  |  | X |  |
| 15 | Results: Baseline data | X |  |  |  |
| 16 | Results: Numbers analysed | X |  |  |  |
| 17 | Results: Outcomes and estimation |  | X |  | 8 |
| 18 | Results: Ancillary analyses | X |  |  |  |
| 19 | Results: Harms |  | x |  | 40 |
| 20 | Discussion: Limitations | X |  |  |  |
| 21 | Discussion: Generalisability | X |  |  |  |
| 23 | Other information: Registration | X |  |  |  |
| 24 | Other information: Protocol |  | x |  | 7-8 |
| 25 | Other information: Funding | x |  |  |  |
| *Aspects of the trial that are directly affected or changed by the extenuating circumstance and are not under the control of investigators, sponsor or funder.  **Aspects of the trial that are modified by the study investigators, sponsor or funder to respond to the extenuating circumstance or manage the direct impacts on the trial. | | | | | |

NB: Page numbers entered above refer to page numbers in the author’s submitted manuscript.
