## Supplementary material for "Feasibility trial of a new digital training package to enhance primary care practitioners’ communication of clinical empathy and realistic optimism": S4

### S4 Appendix: Planned Methods for Original Study Design

#### Participants, Recruitment, and Consent

We recruited general practices with assistance from local Clinical Research Networks (CRNs) in Wessex, Thames Valley and West Midlands, West of England, and Kent Surrey and Sussex. The CRNs advertised the study to local practices who had not been involved in our intervention development work (participants in these studies had seen prototypes of EMPathicO). Interested practices returned an expression of interest to the research team who then liaised with the practice to identify eligible practitioners (primary care providers e.g. GP, physiotherapist, or practice nurse). To be eligible, practitioners had to see people with OA in primary care on a regular basis. To assess feasibility in diverse settings, we aimed to recruit 20 practitioners from 10 practices to include: high/low deprivation index; urban/rural; large/small; training/non-training practices. PCPs were offered feedback on the trial, certificates, CPD guidance, and NHS support costs and research costs to cover their time for participation in line with recommendations from the NIHR-CRN. PCPs randomised to the control group were offered access to the EMPathicO digital training at the end of the study. All PCPs received a participant information sheet and the opportunity to ask any questions before giving consent in writing and/or via the trial website.

Adult patients who were consulting with a participating practitioner were eligible to take part. Patients were excluded who are unable to speak English, unable to consent or complete questionnaires (for example, because of severe mental illness, severe distress, very unwell generally, and difficulty reading or writing). To be eligible for inclusion in the pre-planned OA sub-group, patients had to be consulting a participating PCP in relation to clinically diagnosed hip and/or knee OA, where OA is the only reason for consulting or one of two main reasons for consulting; minimum 45 years old (as per NICE guidance for OA[64]). We aimed to recruit up to 60 patients (3 per PCP) with clinically-diagnosed hip and/or knee OA[65, 66] who are seeking care for OA; and up to 120 other patients. We planned to test the feasibility of multiple approaches to patient recruitment (summarized in Appendix A Table 1). In all approaches, consent was requested separately for (1) patient-completed questionnaires at baseline/post-consultation/follow-up, (2) (a) filming the consultation for the PCP to reflect on and (2) (b) filming the consultation for the research team to analyse, and (3) contact for interview. The consent form requested patient’s contact details for subsequent correspondence regarding post-consultation questionnaires, follow-up questionnaires and qualitative interviews.

To examine the feasibility of recruitment we planned to record: PCP recruitment rates (number of practices and individual PCPs recruited per week as a function of number invited); PCP attrition rates (number of practices and individual PCPs dropping out of the study and reasons given); patient recruitment rates (number of all-consulters and OA patients recruited per PCP per recruitment session); and patient attrition rates (number and proportion of consented all-consulters and OA patients formally withdrawing from the study post-baseline or lost to follow-up, and reasons given).

###### Appendix A Table 1. Approaches to Patient Recruitment

| Recruitment Approach | Raise Awareness of Study | Provide Full Information about Study | Eligibility Screening^a^ | Consent collected | Pre-consultation measures | Post-consultation measures^b^ |
| --- | --- | --- | --- | --- | --- | --- |
| Researcher-in-practice (1) | In general practice, via posters, display screens, reception staff. Pre-consultation. | Researcher in person, in private area. Pre-consultation. | Researcher in person, in private area. Pre-consultation. | Researcher in person, in private area. Pre-consultation. | On paper or on researcher’s device or patient’s device, with researcher support | On paper or on researcher’s or patient’s device, with researcher support |
| Researcher-in-practice (2) | In general practice, via computerised check-in. Pre-consultation. | Researcher in person, in private area. Post-consultation. | Computerised check-in. Pre-consultation. | Computerised check-in (provisional). Researcher in person, in private area (post-consultation) | Computerised check-in. | On paper or on researcher’s or patient’s device, with researcher support |
| Researcher-in-practice (3) | In general practice, via posters, display screens, reception staff. Pre-consultation. | Researcher in person, in private area. Post-consultation. | Researcher in person, in private area. Post-consultation. | Researcher in person, in private area. Post-consultation. | Not collected. | On paper or on researcher’s or patient’s device, with researcher support |
| PCP in consultation (1) | In general practice, via posters, display screens, reception staff. Pre-consultation. | PCP at start of consultation. | PCP at start of consultation. | PCP at start of consultation. | PCP at start of consultation. | On paper or on patient’s device. |
| PCP in consultation (2) | In general practice, via posters, display screens, reception staff. Pre-consultation. | PCP at start of consultation. | PCP at start of consultation. | PCP at start of consultation (provisional). Researcher / research nurse post-consultation. | PCP at start of consultation. | On paper or on researcher’s or patient’s device, with researcher support |
| Mail out (1) | Invitation packs mailed approximately one week in advance to patients with pre-booked appointments. To include cover letter, information sheet, eligibility screening form, baseline measures, and written consent. | | | | | On paper or on patient’s device. |
| Mail out (2) | Database search and mailed invitations to patients to book an appointment with a participating PCP. Those who book an appointment are then invited into the study via Mail out (1) process. | | | | | On paper or on patient’s device. |

^a^ At the end of each recruitment session, PCPs completed a Clinical Record Form (CRF) for all consenting patients to record: patient’s unique identifier for the study (allocated on consent), confirmed clinical diagnosis of OA hip and/or knee, age on day of consultation (<45 or 45 and older), and PCP’s view on whether the patient is unable to consent or complete questionnaires (e.g., because of severe mental illness, severe distress, very unwell generally, and difficulty reading or writing).

^b^ Post-consultation measures to be completed within 3 days. Additional procedures related to post-consultation measures were: including questionnaires in initial approach to patients; PCP handing questionnaires to patient at end of consultation; researchers posting/emailing questionnaires on day of consultation.

##### Randomization and Blinding

Cluster randomisation was conducted at the practice level using a 1:1 ratio. Randomizing individual PCPs would have risked cross-contamination within practices if practitioners had discussed the EMPathicO training with each other.

Stratification was planned for the full trial (by practice size and urban/rural) but was not deemed necessary for the feasibility study (as we were not assessing intervention effectiveness) and is not particularly useful when only randomising 10 practices. Blocked randomisation was planned, with random block sizes of 4 and 6.

The researcher randomly allocated practices to intervention or control after all participating PCPs at the practice had successfully recorded five baseline consultations (including at least one consultation regarding hip and/or knee OA). Allocations were generated using an Excel file pre-programmed by the trial statistician.

The statistician was intended to be blinded to allocation until the analysis was completed. It was not possible to blind PCP participants to allocation, as they would know whether or not they are undertaking the training. Similarly, it was not possible to blind to allocation those researchers involved in supporting the intervention. However, it would be possible to blind the patient participants to allocation, as long as PCPs do not disclose this to their patients. We had planned to explore in the feasibility trial the possibility of blinding some of the research team (e.g. those involved in recruiting and collecting patient data) to allocation.

##### Interventions

###### EMPathicO

Consenting PCPs in practices randomised to the intervention group were asked to work through EMPathicO within 3 weeks.

###### Control Group

Consenting PCPs in practices randomised to the control group were asked to practice as usual throughout the trial. They were asked not to look at their videoed consultations until the end of the trial and were told they could have access to EMPathicO when the trial was finished.

##### Outcome and Process Measures

Table 2 lists all outcome and process measures for each group and time-point. In the original study design, we planned to collect baseline measures pre-consultation, followed by immediate outcomes within 3 days post-consultation and follow-up at 2 weeks post-consultation; this would permit an exploration of the feasibility of these timepoints. In line with the OMERACT-OARSI core outcome domains we asked practices to notify us of the death of any patient participants during the study period.

We had also planned to collect recordings of patient consultations that could then be scored for the presence of EMPathicO behaviours. This would have enabled a direct assessment of the extent to which PCPs implemented the behaviours taught in EMPathicO. We aimed to collect films of 5 pre-randomization baseline consultations per PCP (to include at least 1 OA consultation) and up to 9 consultations per PCP recorded at least 5 weeks after joining the study (post-intervention for the EMPathicO group). PCPs were asked to angle the camera towards the PCP to capture their verbal and non-verbal communication behaviours. The intervention group were asked to film consultations as part of the intervention, so they could review and reflect on their communication behaviours in the reflections section of the intervention. Both groups were asked to film consultations as part of the trial, so that we could examine the feasibility of comparing a sample of baseline and post-randomisation films within and between groups, to directly assess any changes over time in PCP communication behaviour. This method permits a direct measure of communication behaviour to supplement patient-reported perceptions of practitioner empathy and optimism. To minimise selection bias, PCPs were asked to seek consent from sequential patients attending in two to three whole sessions of practice until they had obtained the required number of films (5 pre-randomisation films, 9 post-randomisation films). To minimise possible contamination, the control group were instructed not to review their filmed consultations until they had completed the trial.

###### Appendix A Table 2. Outcome and Process Measures

| Construct | Measure | N items | Pre-EMPathicO | Post- EMPathicO | Pre-consultation | Post-consultation (<3 days) | Follow-up (14 days) |
| --- | --- | --- | --- | --- | --- | --- | --- |
| **Patient Reported Outcome** |  |  |  |  |  |  |  |
| Pain intensity | Numerical Rating Scale | 1 | - | - | ALL | - | ALL |
| Symptoms | Symptom change | 1 | - | - | - | - | ALL |
|  | Symptom bothersomeness | 1 | - | - | ALL | - | ALL |
| OA symptoms | HOOS and KOOS[29-31] |  | - | - | OA | - | OA |
| Satisfaction with consultation | MISS for UK general practice[37] | 21 | - | - | - | ALL | - |
| Enablement | Modified PEI[36] |  | - | - |  | ALL | ALL |
| Health-related quality of life | SF-12 v2[41] [40] | 12 | - | - | - | ALL | ALL |
| Wellbeing | Short Warwick Edinburgh Wellbeing Scale[38] | 7 | - | - | - | ALL | ALL |
| Pain Medication Change | Bespoke Osteoarthritis Pain Medication Questionnaire | 5 | - | - | - | - | ALL |
| Adverse events | Bespoke adverse events form | 2 | - | - | - | - | ALL |
| **Patient Reported Process** |  |  |  |  |  |  |  |
| Perceptions of PCP empathy | CARE[47] | 10 | - | - | - | ALL | - |
| Anxiety | Anxiety subscale of the HADS[62, 63] | 14 | - | - | - | ALL | - |
| Perceptions of PCP response expectancies | Bespoke item | 1 | - | - | - | ALL | - |
| Response expectancies | Expectancy subscale of the CEQ[48] | 3 | - | - | - | ALL | - |
|  | Treatment Expectation Questionnaire (TEX-Q) | 11 | - | - | - | ALL | - |
| Treatment credibility | Credibility subscale of the CEQ[48] | 3 | - | - | - | ALL | - |
| **Practitioner Reported Process** |  |  |  |  |  |  |  |
| Self-efficacy for conveying empathy & optimism | Bespoke self-efficacy scale | 8 | - | PCP |  |  |  |
| Outcome expectancy for conveying empathy & optimism | Bespoke outcome expectancy scale | 8 | - | PCP |  |  |  |
| Intentions to convey empathy & optimism | Bespoke intentions scale | 4 | - | PCP |  |  |  |
| **Directly Assessed Process** |  |  |  |  |  |  |  |
| Practitioner empathy behaviours | Filmed consultations |  | RES | RES |  |  |  |
| Practitioner realistic optimism behaviours | Filmed consultations |  | RES | RES |  |  |  |
| Practitioner intervention usage | LifeGuide data |  |  | RES |  |  |  |

KEY: OA = completed by OA group only; ALL = completed by all patient participants; PCP = completed by primary care practitioner; RES = Researcher assessed.

##### Qualitative Data

We planned to invite participating PCPs and other practice staff who had a role in the trial to take part in a telephone interview or focus group to explore barriers/facilitators to implementing the trial and barriers/facilitators to accessing/ implementing EMPathicO.

We invited a varied sample of patients to take part in a semi-structured telephone interview to explore patients’ experiences of trial processes and measures including their consultation, and questionnaire relevance and burden. We had intended to sample purposively to ensure we interviewed some patients: from each arm of the trial; from different primary care practices; who were recruited using different methods; and who had different patterns of missing data.
