## Supplementary material for "Feasibility trial of a new digital training package to enhance primary care practitioners’ communication of clinical empathy and realistic optimism": S5

### Research Protocol Final v3.0

### The TIP Study: Expectation Management for Patients in Primary Care: Feasibility Trial of a New Digital Intervention for Practitioners

Short name: The TIP Study: Talking in Primary Care

Authors: Felicity Bishop, Hazel Everitt, Jeremy Howick, Paul Little, Christian Mallen, Leanne Morrison, Lucy Yardley, Beth Stuart, Stephanie Hughes, Kirsten Smith, Jane Vennik, Mohana Ratnapalan, Emily Lyness, Hajira Dambha-Miller.

Sponsors: University of Southampton

Funders: National School for Primary Care Research

Reference numbers:

| | |
|---|---|
| University of Southampton/ERGO | 52146 |
| IRAS | 270728 |
| Portfolio | |
| National School for Primary Care Research | 389 |

#### Investigators

**Dr Felicity Bishop**
Associate Professor, Centre for Clinical and Community Applications of Health Psychology, University of Southampton, Southampton, SO17 1BJ


**Dr Hazel Everitt**
Professor of Primary Care Research
Primary Care and Population Sciences
University of Southampton
Aldermoor Health Centre
Aldermoor Close, Southampton SO16 5ST


**Dr Jeremy Howick**
Senior Researcher
Nuffield Dept Primary Care Health Sciences
University of Oxford
Radcliffe Observatory Quarter
Woodstock Road, Oxford OX2 6GG


**Prof Christian Mallen**
NIHR Research Professor in General Practice
Primary Care Sciences
University of Keele
Staffordshire, ST5 5BG


**Dr Leanne Morrison**
Lecturer in Health Psychology, Centre for Clinical and Community Applications of Health Psychology, University of Southampton, Southampton, SO17 1BJ
Primary Care and Population Sciences, University of Southampton, Southampton, SO16 5ST


**Prof Paul Little**
Professor of Primary Care Research
Primary Care and Population Sciences
University of Southampton
Aldermoor Health Centre
Aldermoor Close, Southampton SO16 5ST


**Professor Lucy Yardley**
School of Psychological Science, University of Bristol
Office 1D20, The Priory Road Complex,
Priory Road, Clifton BS8 1TU


**Dr Beth Stuart**

Primary Care and Population Sciences
University of Southampton
Aldermoor Health Centre
Aldermoor Close, Southampton SO16 5ST


#### Researchers

**Mrs Stephanie Hughes**
Research Fellow
Primary Care and Population Sciences
University of Southampton
Aldermoor Health Centre
Aldermoor Close, Southampton SO16 5ST


**Dr Kirsten Smith**
Research Fellow
Primary Care and Population Sciences
University of Southampton
Aldermoor Health Centre
Aldermoor Close, Southampton SO16 5ST


**Dr Jane Vennik**
Research Fellow
Primary Care and Population Sciences
University of Southampton
Aldermoor Health Centre
Aldermoor Close, Southampton SO16 5ST


**Dr Hajira Dambha-Miller**
GP and Research Fellow
Primary Care and Population Sciences
University of Southampton
Aldermoor Health Centre
Aldermoor Close, Southampton SO16 5ST


**Dr Emily Lyness**
Academic Clinical Fellow
Primary Care and Population Sciences
University of Southampton
Aldermoor Health Centre
Aldermoor Close, Southampton SO16 5ST


**Dr Mohana Ratnapalan.**
Academic Clinical Fellow
Primary Care and Population Sciences
University of Southampton
Aldermoor Health Centre
Aldermoor Close, Southampton SO16 5ST


#### Abbreviations

Empathica: Expectation Management for Patients in Primary Care. [the name of the overarching project]

EMPathicO: Empathy and Conveying Optimism. [the name of the digital training intervention that we have developed and are trialling]

GP: General Practitioner

HOOS: Hip and Disability Osteoarthritis Score

KOOS: Knee Injury and Osteoarthritis Score

MISS: Medical Interview Satisfaction Scale

NICE: National Institute for Health and Care Excellence

NRS: Numerical Rating Scale

OA: Osteoarthritis

PCP: Primary Care Practitioner, e.g., GP, physiotherapist, nurse

PEI: Patient Enablement Index

TEX-Q: Treatment Expectation Questionnaire

TSC: Trial Steering Committee

#### Plain English Summary

Osteoarthritis pain is common, costly, and challenging to manage in busy primary care settings. While various drug-based and non-drug-based treatments are recommended, patients still experience pain, poor quality-of-life, and drug side effects. Regardless of which treatment patients receive, excellent practitioner-patient communication can significantly reduce patients' pain while improving quality of life and satisfaction with care. Patients experience less pain after consulting practitioners who show empathy and encourage optimism about treatment. Yet practitioners vary widely in how much they show empathy, use a positive approach, and/or use key non-verbal skills. A simple intervention concentrating on improving key elements of empathy and non-verbal communication is likely to be effective and efficient. We have developed an online training package for primary care practitioners (including General Practitioners - GPs, physiotherapists, and nurses) to enhance their consultation skills to show more empathy, improve their non-verbal communication skills, and encourage patients with osteoarthritis to have positive yet realistic expectations. This training package is called EMPathicO. Ultimately, we aim for our training package to enable practitioners to improve the long-term effectiveness of all drug and non-drug therapies for osteoarthritis pain, reduce patients' pain and improve quality of life.

We plan to conduct a small 'feasibility' trial to help us design a large, fundable, clinical trial to test the online empathy training package against usual GP care.

Our aims for the feasibility trial are to assess a range of ways to recruit practices and their patients to participate in a trial and what approaches are most effective and acceptable. We will also assess ways to consent patients, the practicalities and acceptability video record consultations, ways to collect our proposed outcome measures and assess GP use and experience of the online training tool. We will involve patient representatives in the design of the feasibility study to help ensure proposed procedures are relevant and realistic.

Methods. This feasibility trial will be undertaken in a diverse range of general practices in the South of England and will involve approximately 10 GP practices, 20 primary care practitioners, and up to 280 patients.  Practices will be randomised, and practitioners in 5 of the GP practices will undertake the EMPathicO online training and those in the other 5 GP practices will continue normal practice. Those who continue normal practice will have access to the training after the feasibility study is completed.  Data collected for the feasibility study will include: video recordings of consultations with patients, patient-reported questionnaires, and practitioner-completed questionnaires.  with osteoarthritis to form a baseline and a resource to use during the training.  Patient-reported questionnaires ask them about their symptoms, quality of life, satisfaction and ability to cope with their illness.  Practitioners and practice staff will be asked to participate in focus groups and/or telephone interviews.  Patients will be asked to participate in telephone interviews.  The focus groups and interviews will explore how participants found the feasibility study procedures and the online training.

Results. We will collect information on all the feasibility trial procedures to inform the design of a large trial to test the clinical and cost effectiveness of the online training

#### Expert Summary

Osteoarthritis (OA) pain is prevalent, personally and economically costly, and challenging to manage in busy primary care settings. Pharmacological and non-pharmacological treatments are recommended but patients report adverse effects and limited benefits. Regardless of which treatment patients receive, excellent practitioner-patient communication can significantly improve pain, quality of life, and satisfaction. Previous work suggests that people in pain have better outcomes when practitioners communicate empathically and encourage realistic and positive expectations about treatment. However, this evidence remains to be implemented in practice in a pragmatic format that engages practitioners effectively, and practitioners vary widely in how much they express empathy to their patients. We have addressed this by developing a digital intervention to enhance primary care practitioners' skills in the context of managing OA pain, focusing on empathic communication and expectation management. This training package is called EMPathicO. Furthermore, we anticipate that these skills are also relevant to patients consulting for other conditions. This feasibility study is designed to see how best to evaluate our intervention in an eventual cluster randomised controlled trial in primary care. Our aims and associated objectives are:

1) to establish methods to maximise recruitment and minimise attrition, in practices with a range of socio-demographic areas (barriers and enablers)
    a. to assess recruitment rates associated with different methods of recruitment
    b. to assess retention rates
    c. to identify barriers to recruitment of practices, PCPs, and patients, and ways to overcome them
    d. to identify enablers of recruitment of practices, PCPs, and patients, and ways to harness them
    e. to identify barriers to retention of practices, PCPs, and patients, and ways to overcome them
    f. to identify enablers of retention of practices, PCPs, and patients, and ways to harness them
2) to identify feasible randomisation and consent procedures and finalise inclusion/exclusion criteria
    a. to test the feasibility of cluster randomisation.
    b. to test the feasibility of different ways of taking practitioner and patient consent, as outlined below.
3) to finalise outcome and process measures
    a. to test the practical and ethical feasibility of video-recording consultations
    b. to explore the relevance, feasibility and acceptability of potential outcome measures
    c. to explore the relevance, feasibility and acceptability of potential process measures
    d. to explore options for outcome and process measures for all-consulters and OA consultations
    e. to explore feasible methods of analysing filmed consultations
    f. to establish likely effect sizes
    g. to explore data for indicative changes in outcome and process measures
    h. to explore effective engagement with EMPathicO

To achieve these aims, we will conduct a cluster-randomised feasibility trial in 10 primary care practices (20 practitioners, 80 patients with osteoarthritis, and up to 200 patients attending primary care for other conditions) in the Wessex area. Practices will be randomised (1:1) to our new digital training package or to usual care control. We will explore different ways of recruiting and consenting patients. Patients will complete patient-reported outcome and process measures. Practitioners will complete additional process measures. We will collect extensive data on intervention usage, recruitment and retention rates, and will explore the recording and analysis of consultations. We will conduct semi-structured qualitative telephone interviews with practitioners, practice staff, and patients. The data will be analysed and interpreted to further improve the intervention, refine our planned trial design including procedures, outcome measures, and sample size. We will consider a full trial feasible and apply for funding if: we achieve 70% of our intended recruitment (14 PCPs, 42 OA patients); if we achieve 50% recruitment we will modify methods; if we achieve under 50% we will investigate reasons to see whether it can be addressed in a full trial. AND 2. 70% of recruited GPs log onto the EMPathicO intervention on LifeGuide; if 50% log on we will modify our incentive and/or reminder plan during the feasibility study.

### Background

#### Introduction

In the UK, one third of adults over 45 have sought treatment for osteoarthritis (OA), primarily through general practice, and in 2010 OA was the 11th leading cause of disability. Disability due to OA increased substantially from 1990 to 2010 and will continue increasing with an aging population.[1] NICE Guidelines recommend a patient-centred approach to OA including information and self-management, non-pharmacological interventions (e.g. exercise, manual therapy, aids and devices), and pharmacological management (paracetamol, NSAIDS, corticosteroids, capsaicin, opioids), before considering referral for joint surgery.[2] Non-invasive interventions typically demonstrate only small to moderate effects. Opioids and intra-articular corticosteroids may have larger short-term benefits but patients are concerned about adverse effects and many patients discontinue their use after a few weeks.[3] While these treatments can help patients, their full potential is often not realised.[4] Although practitioners typically perceive musculoskeletal pain to be well-managed, patients report ongoing pain and disability.[5]

Regardless of which therapy a patient receives, excellent practitioner-patient communication has the potential to significantly improve patients' symptoms, quality of life, adherence to and satisfaction with care, producing modest benefits that are comparable to many pharmaceutical interventions.[6-8] Furthermore, sub-optimal consultations represent missed opportunities for benefit and can even be harmful, causing: worse quality of life and symptom management, unwanted prescriptions and non-adherence;[9,10] unnecessary economic costs;[10] deviation from guideline-recommended treatment;[11] and increased complaints and litigation.[12,13] Despite communication skills being essential in medical and allied health professional training, patients still report dissatisfaction with practitioner-patient communication.[14,15] Our recent systematic review showed that the extent to which PCPs express empathy is typically low and varies widely.[16] Fortunately, PCPs are willing to engage in training and even very brief interventions can successfully improve communication skills, including interventions concentrating on non-verbal skills which take no additional time in the consultation and so are likely to be very efficient.[17,18] However, few interventions have been tested clinically for effects on patients' health,[19] few have been sufficiently well described to allow implementation, and (where details are available) most interventions are prohibitively complex, expensive, and time-consuming, which makes engagement and uptake in the current climate in primary care extremely unlikely. Our research addresses these key limitations.

#### Empathico: Enhancing care through empathy and optimism

We are currently finalising a new brief digital training package for PCPs, called EMPathicO. This training package is designed for PCPs to enhance their communication of empathy and optimism through verbal and non-verbal behaviours. Our decision to focus on these aspects was influenced by a recent systematic review conducted by team members demonstrating the potential benefits of communicating empathy and positive expectations[20] and prior work demonstrating the importance of non-verbal communication in primary care.[17,21,22]

Training practitioners to enhance their communication of empathy and optimism through verbal and non-verbal behaviours could enhance care for diverse conditions. Indeed, much of the evidence that underpins the importance of these behaviours for patient outcomes, and that we have drawn on to develop EMPathicO, is derived from studies of various conditions including but not limited to osteoarthritis pain. However, interventions targeted to specific audiences and conditions are likely to be more relevant (to recipients) and possibly more effective, particularly if they are also tailored to individuals.[23,24] Therefore, in developing EMPathicO we aimed to produce a training package that not only has the potential to be broadly applicable to primary care consultations but also includes the necessary content to allow PCPs to tailor their new skills for use in OA consultations.

To develop EMPathicO we used the systematic multi-component person-based approach (PBA) to put intervention users and beneficiaries at the heart of the design and development process.[25] We integrated evidence-based and theory-based approaches[26] to ground our training package in relevant evidence and theory. We used the BCW (Behaviour Change Wheel) and COM-B (Capability, Opportunity, Motivation model of Behaviour) as our comprehensive theoretical framework to guide intervention design and complement the PBA's focus on intervention users' perspectives.[27]

The work that informed the development of EMPathicO is currently being concluded and written up for publication and includes:

- two literature reviews analysing the components of tested interventions included in a recent systematic review[20] related to communicating empathy (7 interventions) and positive expectations (22 interventions), plus components analysis of a further 35 interventions on empathy;
- a major meta-ethnography synthesizing evidence from 21 studies on patients' and 12 studies on PCPs' perspectives on primary care consultations for OA;
- a behavioural analysis of the communication of empathy and positive expectations;
- a qualitative interview study of 20 PCPs' views about digital training in empathic and optimistic communication in general and for patients with OA in particular;
- two qualitative interview studies of patients' views about doctors' communication for OA (using enacted consultations and written vignettes as stimuli for discussion; ongoing).

Findings from this extensive body of development work were used to draft guiding principles and a logic model that were in turn used to guide the development of an initial draft of EMPathicO. We are currently conducting two further 'think aloud' studies with PCPs (n=20) to iteratively refine the training package based on our analysis of PCPs' experiences and perspectives, with reference to the guiding principles and logic model.

#### Aims and Objectives

This feasibility study is needed to establish parameters and methods for a subsequent definitive trial. Following NIHR guidance on pilot/feasibility studies, the aims and associated objectives are:

1) to establish methods to maximise recruitment and minimise attrition, in practices with a range of socio-demographic areas (barriers and enablers)
    a. to assess recruitment rates associated with different methods of recruitment
    b. to assess retention rates
    c. to identify barriers to recruitment of practices, PCPs, and patients, and ways to overcome them
    d. to identify enablers of recruitment of practices, PCPs, and patients, and ways to harness them
    e. to identify barriers to retention of practices, PCPs, and patients, and ways to overcome them
    f. to identify enablers of retention of practices, PCPs, and patients, and ways to harness them
2) to identify feasible randomisation and consent procedures and finalise inclusion/exclusion criteria
    a. to test the feasibility of cluster randomisation.

b. to test the feasibility of different ways of taking practitioner and patient consent, as outlined below.

3) to finalise outcome and process measures
    a. to test the practical and ethical feasibility of video-recording consultations
    b. to explore the relevance, feasibility and acceptability of potential outcome measures
    c. to explore the relevance, feasibility and acceptability of potential process measures
    d. to explore options for outcome and process measures for OA consultations and others
    e. to explore feasible methods of analysing filmed consultations
    f. to establish likely effect sizes
    g. to explore data for indicative changes in outcome and process measures
    h. to explore effective engagement with EMPathicO.

### Methods

#### Design

Mixed methods feasibility trial in primary care, designed to evaluate methods for a cluster-randomised trial of EMPathicO in all-consulters with a pre-planned sub-sample of patients with hip and/or knee OA. Henceforth, the participants who are not consulting with hip and/or knee OA are referred to as 'others'. The activities to be undertaken by practices and practitioners, and patients, are summarised in Figure 1 and Figure 2 respectively. The timings are subject to change during the feasibility trial depending on recruitment rates: for example, if necessary, we will extend the time available for recruiting patients.

**Figure 1: Practice and Practitioner Activities**

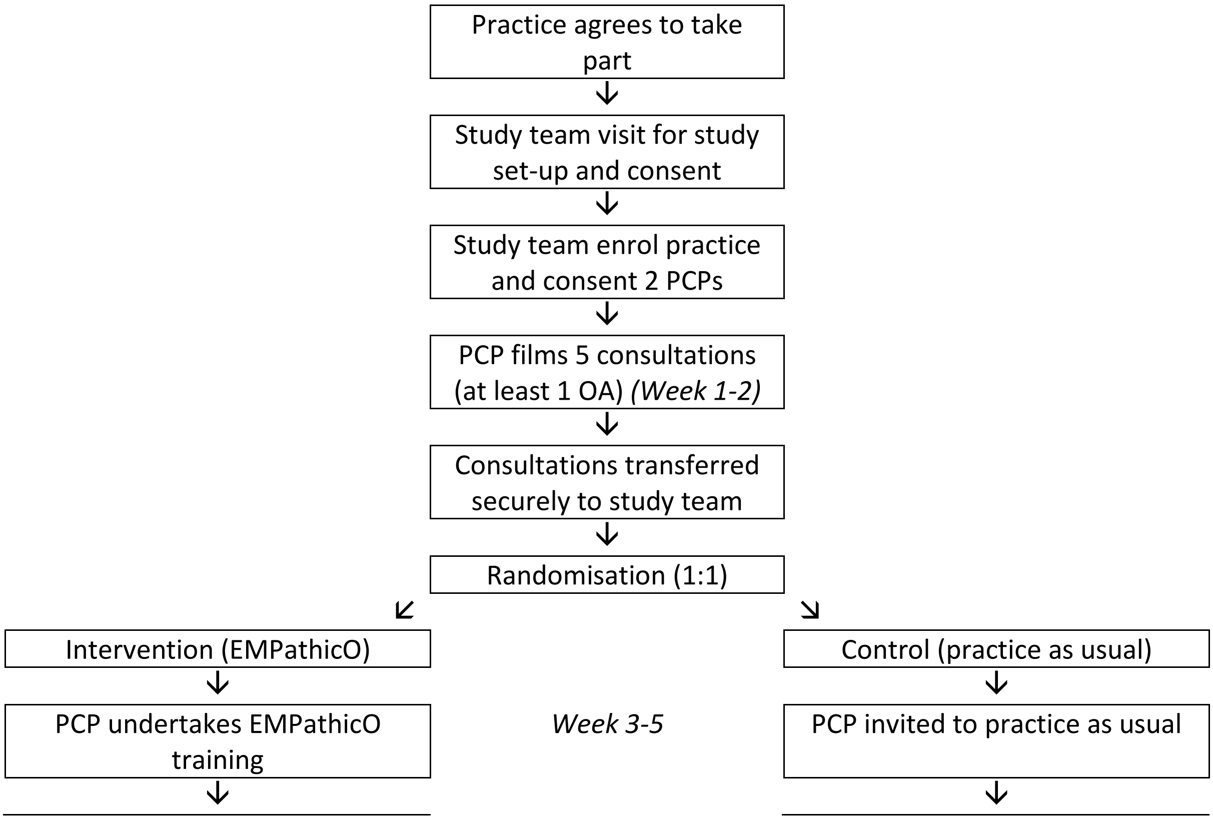

| | | |
|---|---|---|
| PCP recruits patients (3 OA, 6 others) and films consultations | *Week 6-9* | PCP recruits patients (3 OA, 6 others) and films consultations |
| ↓ | | ↓ |
| PCP and practice staff invited to focus group/telephone interview | *Week 10-16* | PCP and practice staff invited to focus group/telephone interview |

Note. * Patients whose consultations are filmed pre-randomisation are not otherwise included in the feasibility trial.

#### Figure 2: Patient Activities (Post-Randomisation)

| EMPathicO | *Timepoint for completion* | Control |
|---|---|---|
| ↓ | | ↓ |
| PCP recruits patients (3 OA, 6 others) and films consultations | *Week 6-9* | PCP recruits patients (3 OA, 6 others) and films consultations |
| ↓ | | ↓ |
| Consent<br>Pre consultation questionnaire<br>Post consultation questionnaire | *Week 6-9* | Consent<br>Pre consultation questionnaire<br>Post consultation questionnaire |
| ↓ | | ↓ |
| Follow-up questionnaires | *Weeks 8-11 (2 weeks post-consultation)* | Follow-up questionnaires |
| ↓ | | ↓ |
| Telephone interview (with a purposive sub-sample) | *Weeks 9-13 (2-4 weeks post-consultation)* | Telephone interview (with a purposive sub-sample) |

As Figure 1 shows, both the control group and the EMPathicO group are being asked to film some consultations at two time-points, before and while enrolling patients in the trial. One could argue that the control group should not be filming any consultations, as filming and reviewing one's consultations is a key, and we believe active, component of EMPathicO. However, we anticipate that any future definitive trial will be more likely to attract funding and more likely to influence practice if we can demonstrate that the content of the training is effective above and beyond any non-specific effects of filming and reviewing one's consultations. While this means that the control group does not constitute usual care, it may approximate usual care for newly qualified GPs who are encouraged to practice reflectively and for whom consultation training has been highlighted as particularly important.[28 29]

In the KEPe-Warm trial, GPs had 1 to 3 weeks to practice implementing their new skills before data collection commenced.[17] We have retained this by allowing 3 weeks for PCPs to undertake the training and practice implementing it in practice before commencing patient recruitment.

#### Participants

Participants will be PCPs (primary care providers e.g. GP, physiotherapist, or practice nurse) who see people with OA in primary care on a regular basis and their patients.

#### Inclusion/Exclusion Criteria

##### Practices:

Inclusion criteria: Primary care practices within Wessex CRN.

Exclusion criteria: Participated in Empathica Development studies 2 or 4 (think aloud studies), as these involve looking at prototypes of the intervention. If they were to be included in this feasibility trial they could contaminate the control arm.

##### Practitioners:

Inclusion criteria: primary care provider (e.g. GP, physiotherapist, or practice nurse) seeing people with OA on a regular basis. While we expect most if not all practitioner-participants to be GPs, we want to try to recruit first-contact primary care physiotherapists and practice nurses too because these practitioners (particularly physiotherapists) will be increasingly involved in managing patients with OA in primary care in the future.

Exclusion criteria: none.

##### Patients:

Inclusion criteria all-consulters: Adults.

Inclusion criteria OA sub-sample: Consulting a participating PCP in relation to clinically diagnosed hip and/or knee OA, where OA is the only reason for consulting or one of two main reasons for consulting; minimum 45 years old (as per NICE guidance for OA[2]).

Exclusion criteria: Patients who are unable to speak English, unable to consent or complete questionnaires (for example, because of severe mental illness, severe distress, very unwell generally, and difficulty reading or writing).

#### Sample Size

##### Practices

To assess feasibility in diverse settings, 10 practices within Wessex CRN will be recruited to include: high/low deprivation index; urban/rural; large/small; training/non-training practices.

##### Practitioners

Up to 20 consenting PCPs.

##### Patients

For randomisation: Up to 60 patients (3 per PCP) with clinically-diagnosed hip and/or knee OA[30 31] who are seeking care for OA; and up to 120 other patients. An additional 100 patients (20 with OA) will be recruited for baseline video-recordings of consultations; these patients will not complete any questionnaires or interviews. Anecdotally we expect that for every 1 adult patient consulting for hip/knee OA there will be approximately 18 adult patients consulting for other conditions. However, we have decided to put an upper limit on the number of others to be recruited to this feasibility trial because:

1. Our main focus during intervention development and trial design has been hip/knee OA and we anticipate showing a larger effect in this more narrowly defined subgroup where pain is a primary complaint.
2. While we anticipate needing a larger sample to detect an effect in the more heterogeneous group of others, we do not anticipate needing a sample that is 18 times larger than the OA subgroup, which may be what we achieve if we simply invite all patients attending a participating PCP until we have 3 patients with OA (based on anecdotal evidence of approximate numbers of OA consultations per surgery). Such over-recruitment would be an unnecessary burden on patients' time and on our limited resources.
3. These numbers will provide enough information in the feasibility trial to be confident in our methods of recruiting these different groups and to estimate the variance in the primary outcome measure to inform power calculations for the planned definitive trial.

##### Randomization.

Cluster randomisation at the practice level, 1:1 ratio, randomising 5 practices to intervention and 5 to control. Randomizing individual PCPs would risk cross-contamination within practices.

In the full trial, we intend to stratify by practice size and location (urban/rural). However, this is not necessary in a feasibility trial (as we are not assessing intervention effectiveness) and is not particularly useful when only randomising 10 practices. Blocked randomisation will be used, with random block sizes of 4 and 6. This will be implemented by means of an Excel file, programmed by our statistician (Dr Beth Stuart).

Randomization will be done after PCPs have successfully recorded five baseline consultations (including at least one consultation regarding hip and/or knee OA). When the researchers have confirmed that all participating PCPs in a practice have successfully recorded their baseline consultations, the research team will enter the practice into a cell in the Excel file which will then randomly allocate and display the trial group.

##### Blinding

The statistician will be blinded to allocation until the analysis has been completed. It is not possible to blind PCP participants to allocation, as they will know whether or not they are undertaking the training. Similarly, it is not possible to blind to allocation those researchers involved in supporting the intervention. However, it is possible to blind the patient participants to allocation, as long as PCPs do not disclose this to their patients. It may also be possible to blind some of the research team (e.g. those involved in recruiting and collecting patient data) to allocation; we will explore this in the feasibility trial.

#### Interventions

##### EMPathicO

EMPathicO comprises 7 modules that take approximately 30 minutes to complete in total, plus additional time to review one's consultations (the duration of which is left to the PCP's discretion). It is currently being finalised and so is still subject to minor changes. The current prototype is summarised in Table 1 and Figure 3.

#### Table 1. Summary of Draft EMPathicO Digital Training Package

| Module | Aims | Content | Key Evidence Source from the EMPathicO development work |
|---|---|---|---|
| 1. Introduction | To address barriers. To engage and enthuse. | Information on the content and duration of the programme and a short quiz about empathy and optimism. | Qualitative interviews with PCPs. |
| 2. Empathy | PCPs to express empathy effectively using verbal and non-verbal behaviours. | Information on latest research in personalisation, validation and “warming up” with multimedia activities | Systematic review and components analysis of empathy intervention trials. Qualitative interviews with patients and PCPs. |
| 3. Optimism | PCPs to express realistic optimism about recommended interventions. | Information on latest research in outcome expectancies with multimedia activities. | Systematic review and components analysis of expectations intervention trials. Qualitative interviews with patients and PCPs. |
| 4. Osteoarthritis | PCPs to tailor their expression of empathy and optimism to account for issues specific to OA. | Specific examples on using empathy and optimism in OA and further information on NICE recommendations for treating OA. | Meta-ethnography of patients’ and PCPs’ perspectives on OA consultations. |
| 5. Reflecting on practice | PCPs asked to reflect on video recordings of up to 5* of their own pre-training consultations (at least one of which concerns hip/knee OA), to identify opportunities to enhance their communication of empathy and realistic optimism. | Instructions on what to look out for when watching consultations. Interactive exercises to identify opportunities for improvement. | KEPe-Warm intervention and think aloud interviews with PCPs. |
| 6. Goal setting | PCPs select up to 3 behaviours to change and formulate an action plan for doing so. | Interactive activity to set goals and make plans based on previous sections. Email triggered in 1-2 weeks to invite participants to record | KEPe-Warm intervention and think aloud interviews with PCPs. |

| | | | |
|---|---|---|---|
| | | additional consultations and review/change goals. | |
| 7. Goal review | PCPs asked to reflect on video recordings of up to 5 of their own post-training consultations (at least one of which concerns hip/knee OA), to review achievement of personal goals. | Goals shown, participants encouraged to reflect on what went well and set/change goals. | KEPe-Warm intervention and think aloud interviews with PCPs. |

Note. * PCPs will be asked to film at least 3 consultations (one with OA) but can choose to film more if they would like to review more as part of the EMPathicO training. In KEPe-Warm GPs typically reviewed no more than 5 of their own pre-training consultations, hence our recommendation here.

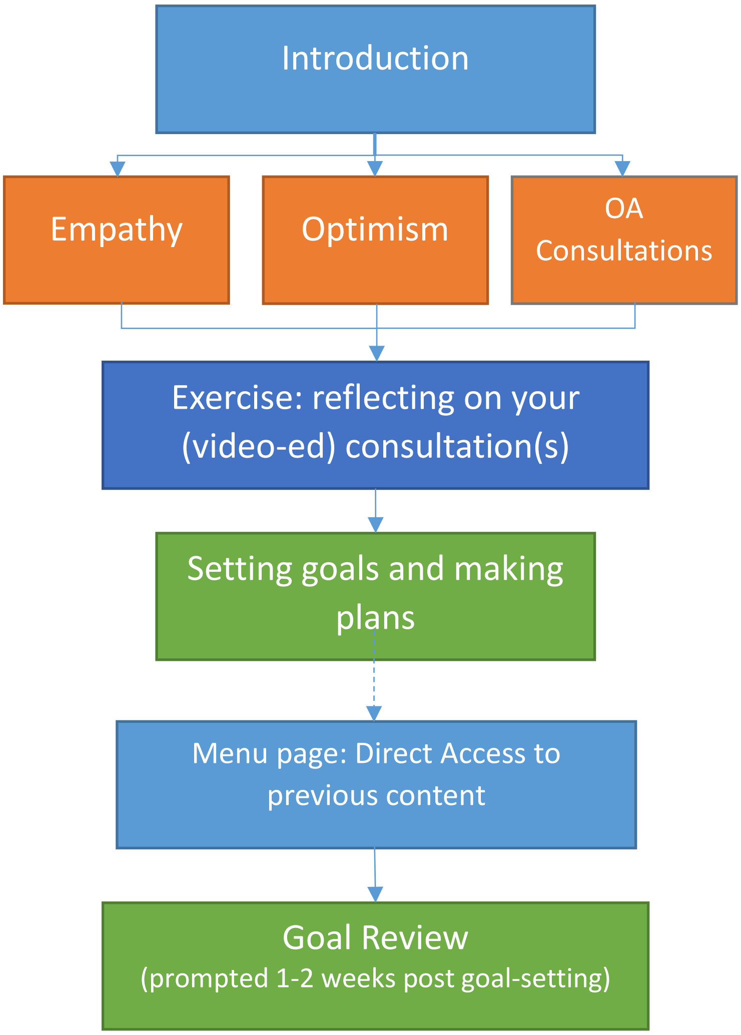

#### Figure 3. Depiction of Flow through EMPathicO

##### Control Group

Consenting PCPs in practices randomised to the control group will be asked to practice as usual throughout the trial. They will be asked not to look at their videoed consultations until the end of the trial. They will be advised that they will be given access to EMPathicO when the trial is finished.

##### Filming Consultations Procedures

As shown in Figure 1, PCPs in both groups are asked to film some consultations at two time points, before randomisation and again post-randomisation, during patient recruitment. In these films, the camera will be angled towards the PCP in order to capture their verbal and non-verbal communication behaviours. Depending on the set-up of the consulting room, this will mean that the patient is highly unlikely to be filmed face-on and much more likely to be filmed from behind or from the side. These films serve two purposes.

The first purpose relates to the intervention and is primarily relevant to the intervention group, who will undertake EMPathicO training in the 1-3 weeks after the pre-randomisation filming. For the intervention group, these pre-randomisation, baseline films will be used to reflect on their practice and to identify changes to implement in response to the EMPathicO training, while the post-randomisation films may be used for self-reflection on the effectiveness of attempts to implement the EMPathicO training. The control group will not have any training from the trial team in the weeks between filming sessions, and will be asked not to review their filmed consultations until they have completed the trial. At this point the control group will be given access to EMPathicO and can use their filmed consultations for self-reflection as part of the training.

The second purpose relates to the trial. We plan to explore the feasibility of analysing a sample of baseline and post-randomisation films (to include at least one hip/knee OA consultation per practitioner). The analysis will involve coding the consultations for evidence of empathic behaviours and the communication of optimism and we anticipate developing a checklist based on EMPathicO to facilitate this process. Comparisons will then be made to estimate between-group similarity at baseline and within group changes from baseline to follow-up. Filming in the control group as well as the intervention group will thus permit us to directly assess any changes over time in PCP communication behaviour, rather than just relying on patient perceptions of PCP communication.

For each set of 5 films needed for the trial, we will ask PCPs to seek consent from sequential patients attending in two to three whole sessions of practice until they have obtained the required number of films. We anticipate that this will be sufficient to capture (a) 5 films for PCPs in the intervention group to reflect on, at least one of which will involve a patient consulting for hip and/or knee OA (in KEPe-Warm, GPs typically reflected on no more than 5 filmed consultations[17]), and (b) 5 films (at least one of which will involve a patient consulting for hip and/or knee OA) for the research team to analyse for evidence of empathic behaviours and the communication of empathy. We consider these as potentially but not necessarily overlapping sets, as some patients may give consent for recordings to be used for the PCP's reflections, but not for being viewed by researchers.

#### Recruitment and Consent Procedures

##### Practices

Practices will be recruited with assistance from the CRN. The CRN will advertise the feasibility study to local practices meeting our inclusion criteria, and endeavouring to ensure we have a range of practices in terms of socio-demographics of the community served, at least one in each of an urban, suburban, and rural setting, at least one large and one small practice, at least one training practice and one non-training practice. This will proceed in discussion with the research team. Interested practices will return an expression of interest to the research team.

##### Practitioners

To be recruited by the research team working with the CRN. The research team will liaise with interested practices, provide information about the study including the PIS for individual practitioners, and be available to answer questions remotely (phone, email).

The research team will then arrange to meet with interested practices and complete the following tasks in person:

1) Introduce the study and go through study set up procedures.
2) Provide recording equipment and instruction on its use. (Instruction will also be provided on the study website.)
3) Collect practice details and formally enrol the practice in the study.

We will trial two approaches to taking consent from individual PCPs.

1) In person, e.g. at the study set up meeting.
2) Online, via the Lifeguide research platform.

PCPs will be offered feedback on the trial, certificates, CPD guidance, and NHS support costs and research costs to cover their time for participation in line with recommendations from the NIHR-CRN. PCPs randomised to the control group will also be offered access to the EMPathicO digital training at the end of the study.

##### Patients

###### *Preferred Approach*

Our preferred approach to patient recruitment, which we will trial in all but one practice, is as follows. A researcher will be situated in the practice during all patient recruitment sessions. We will raise awareness that the study is taking place through posters in public spaces (e.g. reception, waiting rooms), through messages on computerised check-in equipment, and by asking reception staff to mention it to all patients who are booked in to see the participating PCP during a patient recruitment session. The posters, messages, and reception staff will direct patients to the researcher for further information. The researcher will then screen, inform, invite, take verbal and written consent, and collect baseline measures on patient arrival, before the patient is called to see the PCP. They will also ask patients to return to them after seeing the PCP in order to complete the post-consultation measures. We will try different approaches to collecting the baseline and post-consultation measures - on paper, on the researcher's tablet or laptop, on the patient's smartphone or other device that they have with them. The research team will contact (email/phone) any consented patients who do not then return to the researcher to complete the post-consultation measures, reminding them to do so ideally within 3 days of the consultation.

We will choose one practice that already uses EMIS, to try a slightly different approach. Here, we will use an EMIS questionnaire module to raise awareness about the study, screen for eligibility, take a provisional consent, and collect baseline measures on patient arrival and check-in for their appointment. The researcher will then discuss the study in full with the patient after their consultation, offering the option to withdraw or to confirm consent and complete the post-consultation patient-reported measures (as described above). This option has the advantage of minimising extra work for reception staff and ensuring that all patients who check-in electronically for an appointment with a participating PCP will be invited into the study.

The drawback to this preferred approach is that some patients may arrive very close to their appointment time and, if the PCP is running to time, will not have very long to consider the information about the study. We will therefore be very clear that, if patients do consent, then they have the option to change their mind and withdraw consent without penalty by contacting the researchers at any time.

*Alternative Approaches*

If our preferred approach is not working as well as we would like (see progression criteria below), then we would like to try the following opportunistic approaches (1 to 3) and advanced mail out approaches (4-5) to recruiting patients to participate in the trial:

1) PCP to inform, invite, take verbal and written consent, and collect baseline questionnaire at the start of the consultation.
2) PCP to inform, invite, take verbal consent, and collect baseline questionnaire at the start of the consultation. Research nurse/researcher to take written consent post-consultation.
3) Research nurse/researcher in reception/ waiting room to inform, invite, take verbal and written consent, and collect post-consultation outcomes, immediately post-consultation. NB This approach means no pre-consultation baseline data will be collected and that the consultation will not be filmed.
4) Invitation packs to be sent approximately one week in advance to patients with pre-booked appointments with a participating PCP. To include cover letter, PIS and written consent.
5) Database search and mailed invitations to patients to book an appointment with a participating PCP. To include cover letter, PIS and written consent. While this would represent a significant departure from usual practice, we would like to trial this approach if other approaches are proving ineffective.
6) Patient participants will be recruited through social media/patient organisations/snowball sampling. Patients who have planned/have had a recent telephone consultation with their PCP will give consent online and complete pre (if possible) and post consultation questionnaires.

We may also want to try the following different ways of collecting post-consultation questionnaires, if we need to adjust our preferred approach:

7) Post-consultation questionnaires to be included in initial approach to patients.
8) PCP to hand out post-consultation questionnaires at the end of each consultation for patients to complete in the waiting room and hand back at the practice in a sealed envelope or to take home and post back to the research team.

9) Research team to email post-consultation questionnaires on the same day as the consultation, requesting completion within 3 days.

*Recruiting of Patients with OA and Others*

As described above (Sample Size / Patients), post-randomization we intend to recruit up to 60 patients consulting for OA and up to 120 patients consulting for other conditions. We shall closely monitor recruitment to ensure we stop recruiting all-consulters when we have reached 6 per PCP. This will likely mean we then need to focus only on recruiting patients for the OA sub-sample, and therefore we have planned the following options for identifying patients who will be potentially eligible for this sub-sample and who are attending to see a participating PCP. Our preferred approach is to tweak our preferred recruitment approach outlined above, such that posters in public areas will specify the study is for patients consulting with hip and/or knee pain and/or OA, and reception staff will be asked to screen patients and only ask those who are potentially eligible to speak with the researcher. In the practice trialling the EMIS questionnaire module for recruitment we will tweak this to screen for potentially eligible patients with OA too.

#### Box 1. Script for Reception Staff/EMIS questionnaire module to use to identify patients consulting about OA.

| |
|---|
| The [doctor/nurse/physiotherapist] that you're seeing [today/on date] is taking part in some research that you might be eligible for. Can I ask "have you come to see [PCP name] about knee and/or hip pain / OA".<br><br>If NO – that's fine, thank you.<br>If YES hip/knee –continue with *Preferred Approach* outlined above. |

*Patient Consent*

In all approaches, consent will be itemised to garner consent for (1) patient-completed questionnaires at baseline/post-consultation/follow-up, (2) (a) filming the consultation for the PCP to reflect on and (2) (b) filming the consultation for the research team to analyse, and (3) contact for interview. The consent form will request patient's contact details. This will enable researchers to contact patients directly with invitations to complete post-consultation questionnaires, follow-up questionnaires and take part in a qualitative interview via email, mail, and/or telephone. The consent form will ask patients to indicate consent separately for each mode of contact.

Patients recruited through social media/patient organisations/snowball sampling will click on a link to the study information and online information sheet. Interested participants will complete an online consent form prior to completing the study questionnaires.

We have chosen to request consent for contact for interview, rather than consent for interview, at the point of recruitment because when undertaking opportunistic in-practice recruitment there will be variable amounts of time for patients to consider the participant information sheet and make a decision about taking part. The time available will depend on the time between the patient arriving at the surgery and being called in to see the PCP. Therefore, we want to focus on only those aspects of the study for which consent is required at that time point, i.e., the questionnaires and filming the consultation.

We have produced a separate participant information sheet and consent form covering the semi-structured telephone interviews. These will be sent directly from the researchers (by post or email, with a cover letter) to patients who consent to contact for interview, approximately 2-4 weeks post-baseline. Patients will be asked to reply to the researchers (by post or email or phone) and will have the opportunity to ask questions over email and by telephone. Given that the interviews will be conducted over the telephone, we would like to trial the feasibility of obtaining written informed consent for interviews or whether we can obtain verbal consent only for interviews. A script for obtaining verbal consent has been produced.

##### *Confirming Eligibility: CRF*

A Clinical Record Form (CRF) will collect clinical data to confirm each patient meets the eligibility criteria. The researcher will work with the PCP at the end of each recruitment session to complete a CRF for all consenting patients. The CRF will record the following: patient's unique identifier for the study (allocated on consent), confirmed clinical diagnosis of OA hip and/or knee, age on day of consultation (<45 or 45 and older), and PCP's view on whether the patient is unable to consent or complete questionnaires (for example, because of severe mental illness, severe distress, very unwell generally, and difficulty reading or writing).

#### Recruitment and Consent Procedures for Filming Baseline Consultations (Pre-Randomization)

Our preferred approach to seeking patient consent for filming baseline consultations is for the PCP to inform patients and take written consent at the start of the consultation. Consent will be itemised separately for (a) filming the consultation for the PCP to reflect on and (b) filming the consultation for the research team to analyse. To ensure patients have the necessary information to make a decision and the consent process is efficient we will give patients a participant information sheet before they go in to see the PCP. Our preferred approach to this is for reception staff to give a participant information sheet to all patients arriving with an appointment to see the participating PCP during a filming session. Patients will then be able to read the information in the waiting room before going in to see the PCP, depending on the variability in the time available as noted above.

The drawback to this approach is that some patients may arrive very close to their appointment time and, if the PCP is running to time, will not have very long to consider the information sheet. We will therefore be very clear that, if patients do consent, then they have the option to change their mind and withdraw consent without penalty by contacting the researchers at any time.

Furthermore, in this approach the onus is on the PCP for monitoring the numbers of patients consenting to be filmed, and maintaining awareness of when they have achieved the target of filming five consultations including at least one consultation regarding hip and/or knee OA.

If necessary (e.g., if recruitment using our preferred method is slow or the burden on PCPs is too high), we will try using two other procedures for seeking patient consent for filming consultations.

One, we will have a researcher present in the practice and reception staff will direct patients to talk to the researcher about the study. The researcher will talk the patient through the information sheet before they are called in to their appointment. The PCP will then take consent as described in our preferred approach. This will make it easier for the research team to monitor consent and to notify the PCP when five consultations have been filmed (at least one for hip/knee OA).

Two, we will mail out participant information sheets with a cover letter approximately 1 week in advance to all patients booked in to see the PCP during a filming session. The PCP will then discuss the study and take consent as per our preferred approach outlined above. This has the advantage that patients will have longer to consider the request and will have time to contact the research team should they so desire. However, the major disadvantage of this procedure is that it would not be feasible for PCPs to do this in usual practice when undertaking EMPathicO training outside the context of a trial and so substantially reduces the likelihood and ease of ultimately rolling out our training for use as CPD by practicing PCPs.

The research team will review how long it takes to capture all the data required for PCP use during EMPathicO and for the researchers to have access to as part of the trial. We anticipate in particular it will be easier to capture the required "other" consultations than it will be to capture the OA consultation. We will ask the PCP to continue to seek consent to record consultations with patients consulting for hip and/or knee OA until one has been recorded with consent. We would like to try the following means of identifying such patients before proceeding with our preferred approach to consent outlined above.

a) PCP to ask patients "what's the main problem that you're here for".
b) Reception staff to ascertain from patients checking in to see the relevant PCP whether they are coming for hip/knee OA, using the script in Box 1. Reception staff to offer potentially eligible patients the participant information sheet, then alert the PCP to proceed to seek consent.
c) Questionnaire module on EMIS to ask patients when checking in to see the relevant PCP "what's the main problem that you're here for," with fixed response options of "knee and/or hip pain / osteoarthritis" or "something else". If knee and/or hip pain / OA, then EMIS to electronically notify the PCP.

#### Outcomes

Table 2 lists all outcome and process measures for each group and time-point. We will explore using paper forms of the questionnaires and their equivalent online versions at the different measurement points. We will use Qualtrics as our online platform for questionnaires for patients, as it is specifically designed for collecting questionnaire data and therefore more suited to this than Lifeguide. We will use Lifeguide as our online platform for questionnaires for PCPs, as they will already be using Lifeguide to access the intervention (EMPathicO group) and/or instructions for filming consultations (all PCPs), and so it will be familiar to them and avoids the need to log on to a different system.

While in a future definitive trial we would want to collect data on healthcare utilisation, for example from a notes review, we are confident this is a reasonably standard process that can be managed successfully and is therefore not a priority for looking at in this feasibility trial. It is more important in this trial to assess the feasibility of our planned patient reported outcomes, process measures, and our more specialist need to record consultations.

#### Table 2. Outcome and Process Measures

| Construct | Measure | N items | Pre-intervention | Post-intervention | Pre-consultation | Post-consultation | Follow-up |
|---|---|---|---|---|---|---|---|
| **Patient Reported Outcome** | | | | | | | |
| Pain intensity | Numerical Rating Scale | 1 | - | - | ALL | - | ALL |
| Symptoms | Symptom change | 1 | - | - | - | - | ALL |
| | Symptom bothersomeness | 1 | - | - | ALL | - | ALL |
| OA symptoms | HOOS and KOOS[32-34] | | - | - | OA | - | OA |
| Satisfaction with consultation | MISS for UK general practice[35] | 21 | - | - | - | ALL | - |
| Enablement | Modified PEI[36] | | - | - | | ALL | ALL |
| Health-related quality of life | SF-36[37] | 36 | - | - | - | ALL | ALL |
| | OR | | | | | | |
| | SF-12 v2[38 39] | 12 | - | - | - | ALL | ALL |
| Wellbeing | Short Warwick Edinburgh Wellbeing Scale[40] | 7 | - | - | - | ALL | ALL |
| Pain Medication Change | Bespoke Osteoarthritis Pain Medication Questionnaire | 5 | - | - | - | - | ALL |
| Adverse events | Adverse events form | 2 | - | - | - | - | ALL |
| **Patient Reported Process** | | | | | | | |
| Perceptions of PCP empathy | CARE[41] | 10 | - | - | - | ALL | - |
| Anxiety | HADS[42 43] | 14 | - | - | - | ALL | - |
| Perceptions of PCP response expectancies | Bespoke item | 1 | - | - | - | ALL | - |
| Response expectancies | Expectancy subscale of the CEQ[44] | 3 | - | - | - | ALL | - |
| | Treatment Expectation Questionnaire (TEX-Q) | 11 | - | - | - | ALL | - |
| Treatment credibility | Credibility subscale of the CEQ[44] | 3 | - | - | - | ALL | - |
| **Practitioner Reported Process** | | | | | | | |
| Self-efficacy for conveying empathy & optimism | Bespoke self-efficacy scale | 8 | - | PCP | | | |
| Outcome expectancy for conveying empathy & optimism | Bespoke outcome expectancy scale | 8 | - | PCP | | | |
| Intentions to convey empathy & optimism | Bespoke intentions scale | 4 | - | PCP | | | |
| **Directly Assessed Process** | | | | | | | |
| PCP empathy behaviours | Filmed consultations | | RES | RES | | | |
| PCP positive response expectancy statements | Filmed consultations | | RES | RES | | | |

| | | | | |
|---|---|---|---|---|
| PCP intervention usage | LifeGuide data | | | RES |

KEY: OA = completed by OA group only; ALL = completed by all patient participants; PCP = completed by primary care practitioner; RES = Researcher assessed.

#### Patient Reported Outcomes

##### Timing and Modality

We will explore whether it is possible to assess patient-reported outcomes at pre-consultation baseline, as this will enable us to control for baseline statistically, making the final analyses more powerful. If we cannot find a feasible way of collecting pre-consultation baseline data then we will not propose doing this for the subsequent full trial.

The post-consultation measures will be assessed as soon as possible after the index consultation, ideally within 3 days. This is because effects on outcomes such as satisfaction with the consultation and patient enablement, and possibly also those outcomes capturing symptom perception, are likely to be immediate. It is also important to assess process measures related to the consultation (e.g. patient perception of PCP empathy) as soon as possible after the index consultation in order to reduce potential recall bias.

The follow-up time-point is two weeks after the index consultation. In a full trial we would anticipate wanting a longer follow-up period (e.g. 1-3 months, and even longer for health economic outcomes). However, we do not have time for this within our current funding. Furthermore, a 2-week follow-up still enables us to test the feasibility of collecting follow-up data directly from patients. This timing also allows us to collect some indicative data on early attrition which we can combine with insights from similar studies with longer follow-ups to estimate likely attrition rates for the full trial.

##### Measures Completed by the OA Group

The patient-reported outcomes will differ slightly between the patients consulting for hip and/or knee OA and others, in order to ensure outcomes are as specific as possible, relevant to the patients, and map onto our logic model. For the OA sub-group, we have been guided by the recent publication of new OMERACT-OARSI core outcome domains for trials in hip and/or knee OA.[45] This paper specifies that the following five core outcome domains must be measured: pain, physical function, quality of life, patient global assessment of the target joint, and adverse events including mortality. The follow-up paper, not due for 2 years, will recommend specific measures of each domain.

The short form of the Hip and Disability Osteoarthritis Score (HOOS-12) and the Knee Injury and Osteoarthritis Score (KOOS-12) are our candidate disease-specific primary outcome measures for hip and knee osteoarthritis. The HOOS-12 and the KOOS-12 each assess pain, function, and quality of life, and produce an overall summary hip/knee impact score respectively. We will use the brief 12-item formats, which demonstrate promising psychometric properties and reduced participant burden compared to the full 40-item versions.[32-34] However, it is worth noting that the HOOS-12 and KOOS-12 were validated in patients undergoing joint replacement surgery and so it is important to explore their relevance to primary care patients in this feasibility study.

To assess the core outcome domain of patient global assessment of target joint,[45] the OA group will be asked to complete the symptom change item for all-consulters modified for osteoarthritis. This asks patients to rate specifically their knee or hip symptoms now compared to two weeks ago.

A pain numerical rating scale will also be used in the OA sub-sample, as it is likely to be practically feasible for patients to complete a single-item pain score before a consultation; whereas completing the 12-item HOOS or KOOS at a pre-consultation baseline would be more challenging, but we will explore whether it is feasible as part of this feasibility study.

Changes in osteoarthritis pain medication will be assessed using a bespoke Osteoarthritis Pain Medication Questionnaire. This instrument asks patients to list all the osteoarthritis pain medications they are using (to include all tablets, medicines, gels, and creams) and to rate whether and how, since starting the study, they have changed the amount that they use. In designing this instrument we drew on the design of the previously validated but complex Medication Change Questionnaire.[46]

#### Measures Completed by All Participants

All patients will be asked to complete measures of symptoms, patient enablement, satisfaction, well-being, and quality of life as shown in Table 2.

We will explore the feasibility of two candidate primary outcomes that can apply to both OA consulters and others: symptom change, and symptom bothersomeness. The symptom change item asks patients to rate their overall symptoms now compared to two weeks ago, and was adapted from the COOP-WONCA charts.[47 48] The symptom bothersomeness item asks patients to rate how bothersome their symptoms are, and was adapted from the item developed to assess severity of back pain in primary care.[49] These are both single item generic measures, feasible to collect from a large number of patients with diverse health conditions. They add to the wellbeing and quality of life scales by specifically capturing patients' perceptions of symptoms.

The Patient Enablement Index (PEI) captures the extent to which patients feel confident and empowered by a consultation to cope with their illness, to keep healthy and to help themselves. [36] The original publication described six items with 4 response options (much better/never/same or less/not applicable). Following other studies in the primary care and LifeGuide teams, we will modify the response scale to a 7-point agree-disagree Likert scale in order to increase sensitivity to change.

The Medical Interview Satisfaction Scale[50] (MISS) measures patient satisfaction with the consultation and was used detect post-intervention group differences in the Kepe Warm study.[17] We have chosen the version of the MISS that has been specifically adapted and revalidated to be culturally appropriate for UK primary care. Patient satisfaction with the consultation is an important outcome for patients.

Patient wellbeing will be assessed using the Short Warwick Edinburgh Wellbeing Scale.[40] The Warwick Edinburgh Wellbeing Scale underwent extensive development, focuses exclusively on positive aspects of wellbeing, and captures both hedonic and eudaimonic aspects of mental health.[51] The short version is quicker to complete and retains robust psychometric properties as a unidimensional interval level scale. [40]

For quality of life, we will use the SF-36

##### Adverse Events Monitoring

All patients will be asked to complete an adverse events form at the follow-up measurement point. This form is adapted from the ACTIB trial[52] and asks whether, since starting the study, participants have had any of the following events: a life threatening event, admission to hospital where you had to stay overnight, other medical events requiring medical attention. They are asked to provide details of any such events.  They are then asked “Has your health been adversely affected since the start of the study?” with Yes/No response options, and space for details if ‘Yes’.  In line with the new OMERACT-OARSI core outcome domains for trials in hip and/or knee OA[45] we will ask practices to notify us of the death of any patient participants during the patient-participation time period. Adverse events will be notified to relevant bodies, by the research team, as required.

#### Process Measures

A series of process measures will be used to assess key variables from our draft logic model (see Figure 4) that we hypothesise mediate the relationship between undergoing EMPathicO training and improved patient outcomes.

**Figure 4. Draft Logic Model for EMPathicO**

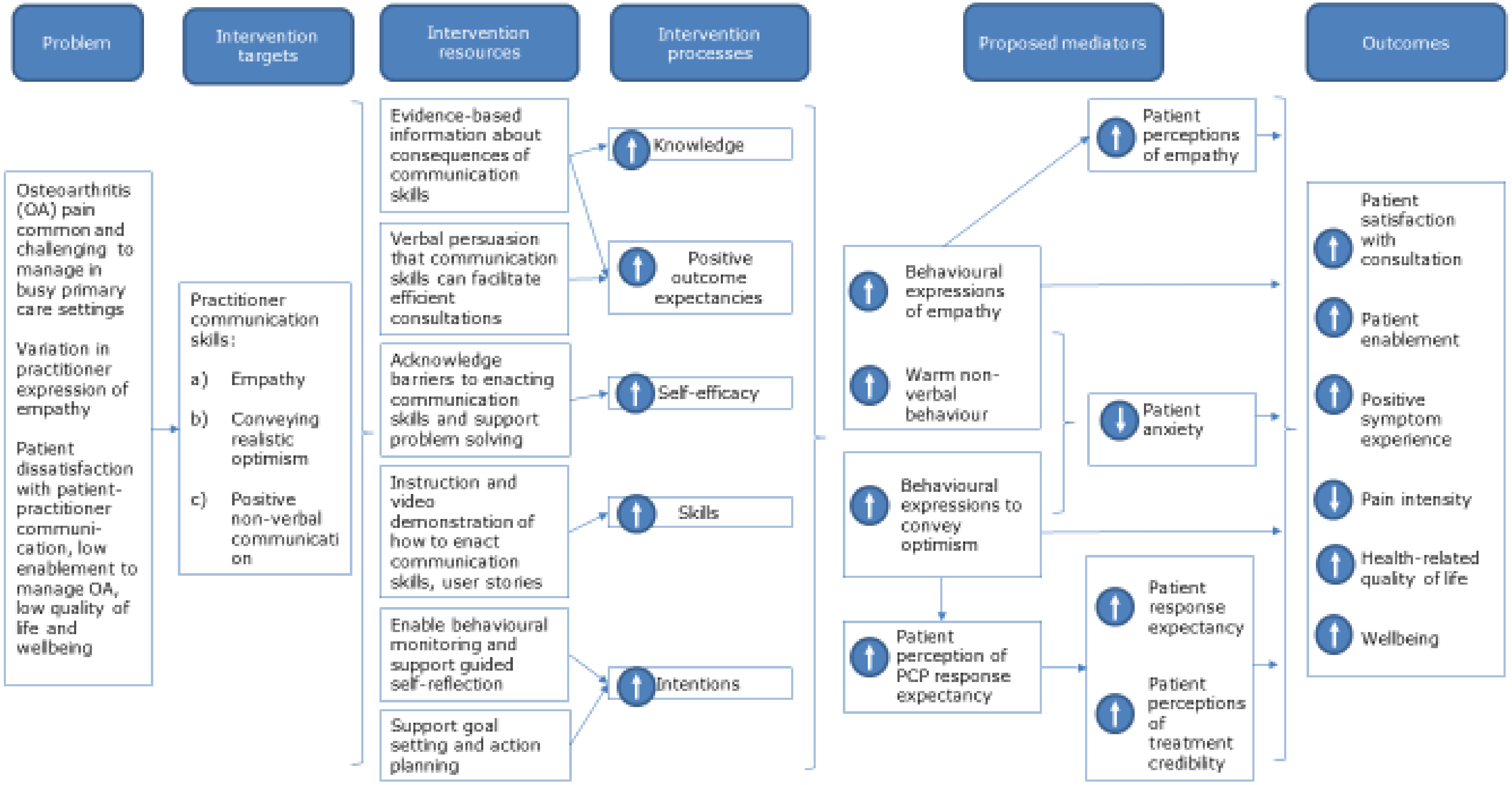

#### Directly Assessed

Intervention usage data from LifeGuide; this will tell us: when and for how long PCPs logged on to EMPathicO; which content was accessed (and for interactive components, engaged with) and for how long; the order in which content was accessed. Engaging with EMPathicO is an important component of our logic model. Collecting this data may suggest essential and non-essential parts of the intervention and will help us to understand engagement with EMPathicO, potentially informing further tweaks to the intervention before final full trial.

Films of 5 baseline consultations per PCP (at least 1 OA) and up to 9 consultations recorded at least 5 weeks after joining the study (post-intervention for the EMPathicO group). If all patient participants in the feasibility trial consent to having their consultation filmed (which we think is unlikely) then we could have a maximum of 280 filmed consultations (100 baseline consultations and 180 post-5 week consultations).

#### Practitioner Reported

Practitioners will be asked to complete newly developed self-report measures of self-efficacy, outcome expectancy, and intentions for conveying empathy and optimism in consultations. Bespoke items have been drafted for this study as it is important to assess these constructs in relation to the specific behaviours that we are targeting, i.e. conveying empathy and optimism, and existing scales do not do this. We have developed our items following recommendations from Bandura's work on outcome expectancies and self-efficacy,[53] the Theory of Planned Behaviour on intentions,[54 55] and the Health Action Process Approach on coping efficacy.[56] For example, Bandura recommends items assessing self-efficacy should concern the performance of a well-specified behaviour and should assess respondent's belief in their ability to perform that behaviour in difficult or challenging circumstances.[53] This idea overlaps with coping self-efficacy in the Health Action Process Approach. Bandura further advises that such circumstances or impediments should be identified from qualitative work with the target respondent group and that items should capture a range of circumstances to help avoid floor or ceiling effects. In developing our self-efficacy items we have therefore drawn on the findings from our interviews with PCPs conducted during the Empathica Development project.

#### Patient Reported

Patient perceptions of PCP clinical empathy will be assessed using the 10-item CARE.[41] This is a validated, reliable, questionnaire that has been used extensively in UK primary care settings to assess patient perceptions of GP clinical empathy. EMPathicO aims to improve PCPs' communication of clinical empathy, and we expect that any such changes, to be clinically meaningful, should be noticed by patients. Patients' perceptions of clinical empathy are therefore an important component of the logic model.

Patient perceptions of PCP response expectancies will be assessed using a bespoke single item drafted for this study (as we could find no existing measure of this construct). EMPathicO aims to encourage PCPs to communicate positive but realistic expectations about the effectiveness of recommended or prescribed medications, management plans, therapies, referrals, etc. Therefore, we expect that patients in the intervention group should be able to perceive their PCP holding generally positive expectations as to the effectiveness of their treatment.

The Credibility Expectancy Questionnaire (CEQ)[44] will be used to assess patient response expectancies and perceptions of treatment credibility. The expectancy subscale assesses the extent to which patients believe their symptoms will improve.  The credibility subscale assesses the extent to which patients believe their treatment to be credible in general for their condition.  The CEQ is reliable and valid and has been used across many diverse settings and patient populations, including OA and primary care.

We will also assess patient response expectancies with a recently developed questionnaire specifically designed to assess patient expectations with respect to the outcome of medical treatments, the Treatment Expectation Questionnaire (TEX-Q) (unpublished data, Meike Shedden-Mora, Jannis Alberts, Keith Petrie, Johannes Laferton, Yvonne Nestoriuc & Bernd Löwe, University Medical Center Hamburg-Eppendorf, Germany).  This questionnaire is a better fit than the CEQ which instead seems to capture more general outcome expectancies that are not closely linked to a particular treatment.  However, the TEX-Q is only now being validated in English and so we do not wish to rely on it alone as our only measure of response expectancies.  By including both the CEQ and TEX-Q in this study we plan to provide additional validation data that we can process with advice from the TEX-Q team with whom we are collaborating.

#### Recruitment Outcomes

PCP recruitment rates, i.e., number of practices and individual PCPs recruited per week as a function of number invited.

PCP intervention usage rates, i.e., number and proportion of consented PCPs logging on to the study website on LifeGuide.

Patient recruitment rates, i.e., number of all-consulters and OA patients recruited per PCP per recruitment session/day/week.  We would also like to collect from practice admin the number of patients seen per PCP per recruitment session.

PCP attrition rates, i.e., number of practices and individual PCPs dropping out of the study and reasons given (if any).

Patient attrition rates, i.e., number and proportion of consented all-consulters and OA patients formally withdrawing from the study post-baseline or lost to follow-up, and reasons given (if any).

Feasibility of timing of outcome and process measures.

#### Qualitative Data

We will invite participating PCPs and other practice staff who had a role in the trial to take part in a focus group (separate control/intervention) or a telephone interview.  Participants will discuss the barriers/facilitators to implementing the trial and barriers/facilitators to accessing/ implementing EMPathicO.  We may run more than one focus group for PCPs in each trial group, depending on participation rates, in order to minimise travel time for PCPs and ensure focus groups are of a suitable size to promote inclusive discussion (approximately 4-8 participants[57]).  We may need to interview some PCPs on the telephone if it proves prohibitively difficult to organise focus groups.  If we do this, the focus group topic guide will be adapted slightly to ensure it is appropriate for an individual interview, but the topics discussed will remain the same.  Initial analysis will begin shortly after commencing the focus groups/interviews, allowing data collection and analysis to proceed iteratively and to inform each other.  We will stop interviewing PCPs and staff when we have spoken to a range of individuals and have generated sufficient data to address our research objectives.

We will invite a varied sample of patients to take part in a semi-structured telephone interview with a researcher.  We will sample patients to ensure we speak with some patients: from each arm of the trial; from different primary care practices; who were recruited using different methods; and who had different patterns of missing data.  Interviews will explore patients' experiences of trial processes and measures including questionnaire relevance and burden.  Initial analysis will begin shortly after commencing the interviews, allowing data collection and analysis to proceed iteratively and to inform each other.  Compared to face to face interviews, telephone interviews reduce the need for time-consuming, polluting, and costly travel, and when interviewers are well-trained can still produce good quality, rich, data.  We will stop interviewing patients when we have spoken to a range of individuals and have generated sufficient data to address our research objectives.

#### Planned Analysis

Data will be downloaded from LifeGuide and Qualtrics, cleaned, and imported into SPSS/STATS for analysis.

Scale scores on all outcome and process measures as appropriate will be computed following published guidelines.

We will examine the psychometric properties of all bespoke items and the TEX-Q (previously validated in German but not English) before included them in analyses.

Trends and effect sizes on process measures and patient reported outcomes will be examined.

Recruitment rates, attrition rates, and intervention usage data from LifeGuide will be examined.

We will explore how to analyse the films of baseline and post-randomisation consultations.  This will include an exploration of how to sample films for analysis; how to code them reliably for the presence of techniques trained in EMPathicO using a simple checklist to be devised when the intervention is finalised; and how to explore for indicative evidence of PCP behaviour change following intervention.

We will analyse qualitative data descriptively using content analysis, possible supplemented by thematic analysis if time allows.  Qualitative data from PCPs and patients will be analysed to suggest improvements to EMPathicO and to explore potential mechanisms of action that may or may not be consistent with the logic model.  Qualitative data from PCPs and practice staff and patients and researchers' field notes will be analysed to suggest improvements to trial procedures.  Qualitative data from patients will be analysed to inform choice of process and outcome measures for the full trial.  Multiple researchers will be involved in the qualitative analysis to guard against idiosyncratic or overly-selective coding.  NVivo will be used to facilitate coding and ensure an audit trail of the analysis is maintained.

The full trial draft power calculation is conservatively based on systematic reviews suggesting a standardized effect size of 0.25 for optimising expectations; sample size calculations will be further refined by calculating completion rates, ICCs, and standard deviations here.

The intervention and full trial design will be finalised after considering both quantitative and qualitative findings.

Table 3 summarises the trial objectives and maps them to data collected and the planned analyses.

**Table 3. Summary of Objectives, Associated Data, and Planned Analyses**

| Objective | Data Collected | Planned Analysis |
|---|---|---|
| 1) to establish methods to maximise recruitment and minimise attrition, in practices with a range of socio-demographics (barriers and enablers) | | |
| a. to assess recruitment rates associated with different methods of recruitment | Recruitment rates by recruitment method, practice characteristics, and trial arm | Cross tabulation |
| b. to assess retention rates | Retention rates by recruitment method, practice characteristics, and trial arm | Descriptive |
| c. to identify barriers to recruitment of practices, PCPs (primary care practitioner e.g. GP, physiotherapist, or practice nurse), and patients, and ways to overcome them | Qualitative interviews and focus groups with patients, PCPs, and practice staff | Qualitative content analysis |
| d. to identify enablers of recruitment of practices, PCPs, and patients, and ways to harness them | Qualitative interviews and focus groups with patients, PCPs, and practice staff | Qualitative content analysis |
| e. to identify barriers to retention of practices, PCPs, and patients, and ways to overcome them | Qualitative interviews and focus groups with patients, PCPs, and practice staff | Qualitative content analysis |
| f. to identify enablers of retention of practices, PCPs, and patients, and ways to harness them | Qualitative interviews and focus groups with patients, PCPs, and practice staff | Qualitative content analysis |
| 2) to identify feasible randomisation and consent procedures and finalise inclusion/exclusion criteria | | |
| a. to test the feasibility of cluster randomisation. | Researchers' field notes<br>Qualitative interviews / focus groups with practice staff | Descriptive |
| b. to test the feasibility of different ways of taking patient consent, as outlined below. | Researchers' field notes | Descriptive |

| | | |
|---|---|---|
| | Qualitative interviews and focus groups with patients, PCPs, and practice staff | |
| 3) to finalise outcome and process measures | | |
| a. to test the practical and ethical feasibility of video-recording consultations | Researchers' field notes<br>Qualitative interviews and focus groups with patients, PCPs, and practice staff | Descriptive |
| b. to explore the relevance, feasibility and acceptability of potential outcome measures | Qualitative interviews and focus groups with patients and PCPs | Directed content analysis |
| c. to explore the relevance, feasibility and acceptability of potential process measures | Qualitative interviews and focus groups with patients and PCPs | Directed content analysis |
| d. to explore options for outcome and process measures for OA consultations and others | Qualitative interviews and focus groups with patients and PCPs.<br>Missing data/completion rates | Directed content analysis.<br><br>Descriptive |
| e. to explore feasible methods of analysing filmed consultations | Filmed consultations | Coding for evidence of Empathico techniques; reliability analysis. |
| f. to establish likely effect sizes | Questionnaire data | Effect size analysis |
| g. to explore data for indicative changes in outcome and process measures | Questionnaire data | Descriptive/Process |
| h. to explore effective engagement with EMPathicO | LifeGuide intervention usage data | Usage/Process |

#### Project Management

Table 4 shows the study Gantt chart. We plan to stagger recruitment and data collection across GP practices, commencing patient recruitment from approximately 2 practices at any one time. Each practice will initially have up to 5 weeks to recruit their patients (for those recruiting over Easter; 4 weeks otherwise). Allowing time to monitor and adjust our methods accordingly, the overall patient recruitment period will last a minimum of 14 weeks.

We will continue to meet regularly (typically weekly) to ensure effective team-working on this complex project, with monthly trial management group meetings and regular reporting as requested to our independent Trial Steering Committee (TSC). An additional Data Monitoring Committee was deemed not necessary by the research team and the TSC, as this is a low risk feasibility trial and we have a statistician on the TSC.

#### Ongoing Monitoring and Success Criteria

We will consider a full trial feasible and apply for funding if: 1. We achieve 70% of intended recruitment (14 PCPs, 42 patients); if we achieve 50% recruitment we will modify methods; if we achieve under 50% we will investigate reasons to see whether it can be addressed in a full trial. AND 2. 70% of GPs log onto LifeGuide; if 50% log on we will modify our incentive and/or reminder plan. We will aim for 80% completion of patient reported outcomes and change procedures if below 70%. We will monitor progress against these criteria at weekly team progress meetings and monthly trial management groups.

**Table 4. Gantt Chart for Feasibility Trial**

| Name | Begin date | End date |
|---|---|---|
| Feasibility trial gantt | 06/01/20 | 24/09/20 |
| Recruit and set up 10 practices | 06/01/20 | 27/03/20 |
| Recruit and set up practice 1 | 06/01/20 | 17/01/20 |
| Recruit and set up practice 2 | 06/01/20 | 17/01/20 |
| Recruit and set up practice 3 | 20/01/20 | 31/01/20 |
| Recruit and set up practice 4 | 20/01/20 | 31/01/20 |
| Recruit and set up practice 5 | 17/02/20 | 28/02/20 |
| Recruit and set up practice 6 | 17/02/20 | 28/02/20 |
| Recruit and set up practice 7 | 02/03/20 | 13/03/20 |
| Recruit and set up practice 8 | 02/03/20 | 13/03/20 |
| Recruit and set up practice 9 | 16/03/20 | 27/03/20 |
| Recruit and set up practice 10 | 16/03/20 | 27/03/20 |
| PCPs film 3 baseline consultations | 20/01/20 | 10/04/20 |
| Practice 1 filming | 20/01/20 | 31/01/20 |
| Practice 2 filming | 20/01/20 | 31/01/20 |
| Practice 3 filming | 03/02/20 | 14/02/20 |
| Practice 4 filming | 03/02/20 | 14/02/20 |
| Practice 5 filming | 02/03/20 | 13/03/20 |
| Practice 6 filming | 02/03/20 | 13/03/20 |
| Practice 7 filming | 16/03/20 | 27/03/20 |
| Practice 8 filming | 16/03/20 | 27/03/20 |
| Practice 9 filming | 30/03/20 | 10/04/20 |
| Practice 10 filming | 30/03/20 | 10/04/20 |
| PCPs undertake training/practice as usual | 03/02/20 | 01/05/20 |
| Practice 1 training/usual practice | 03/02/20 | 21/02/20 |
| Practice 2 training/usual practice | 03/02/20 | 21/02/20 |
| Practice 3 training/usual practice | 17/02/20 | 06/03/20 |
| Practice 4 training/usual practice | 17/02/20 | 06/03/20 |
| Practice 5 training/usual practice | 16/03/20 | 03/04/20 |
| Practice 6 training/usual practice | 16/03/20 | 03/04/20 |
| Practice 7 training/usual practice | 30/03/20 | 17/04/20 |
| Practice 8 training/usual practice | 30/03/20 | 17/04/20 |
| Practice 9 training/usual practice | 13/04/20 | 01/05/20 |
| Practice 10 training/usual practice | 13/04/20 | 01/05/20 |
| Recruit patients into trial and film consults | 24/02/20 | 29/05/20 |
| Practice 1 recruit patients | 24/02/20 | 20/03/20 |
| Practice 2 recruit patients | 24/02/20 | 20/03/20 |
| Practice 3 recruit patients | 09/03/20 | 03/04/20 |
| Practice 4 recruit patients | 09/03/20 | 03/04/20 |
| Practice 5 recruit patients | 06/04/20 | 08/05/20 |
| Practice 6 recruit patients | 06/04/20 | 08/05/20 |
| Practice 7 recruit patients | 20/04/20 | 22/05/20 |
| Practice 8 recruit patients | 20/04/20 | 22/05/20 |
| Practice 9 recruit patients | 04/05/20 | 29/05/20 |
| Practice 10 recruit patients | 04/05/20 | 29/05/20 |
| Data Collection | 09/03/20 | 26/06/20 |
| Collect patient follow-up data | 09/03/20 | 12/06/20 |
| Telephone interview patients | 23/03/20 | 22/05/20 |
| Phase 1 PCP focus groups / interviews | 06/04/20 | 01/05/20 |
| Phase 2 PCP focus groups / interviews | 01/06/20 | 26/06/20 |
| Download and clean Lifeguide data | 01/06/20 | 05/06/20 |
| Finish data collection | 26/06/20 | 26/06/20 |
| Data Analysis | 03/02/20 | 31/07/20 |
| Finish data analysis | 31/07/20 | 31/07/20 |
| Write up | 31/07/20 | 24/09/20 |
| Study close and archiving | 31/07/20 | 24/09/20 |

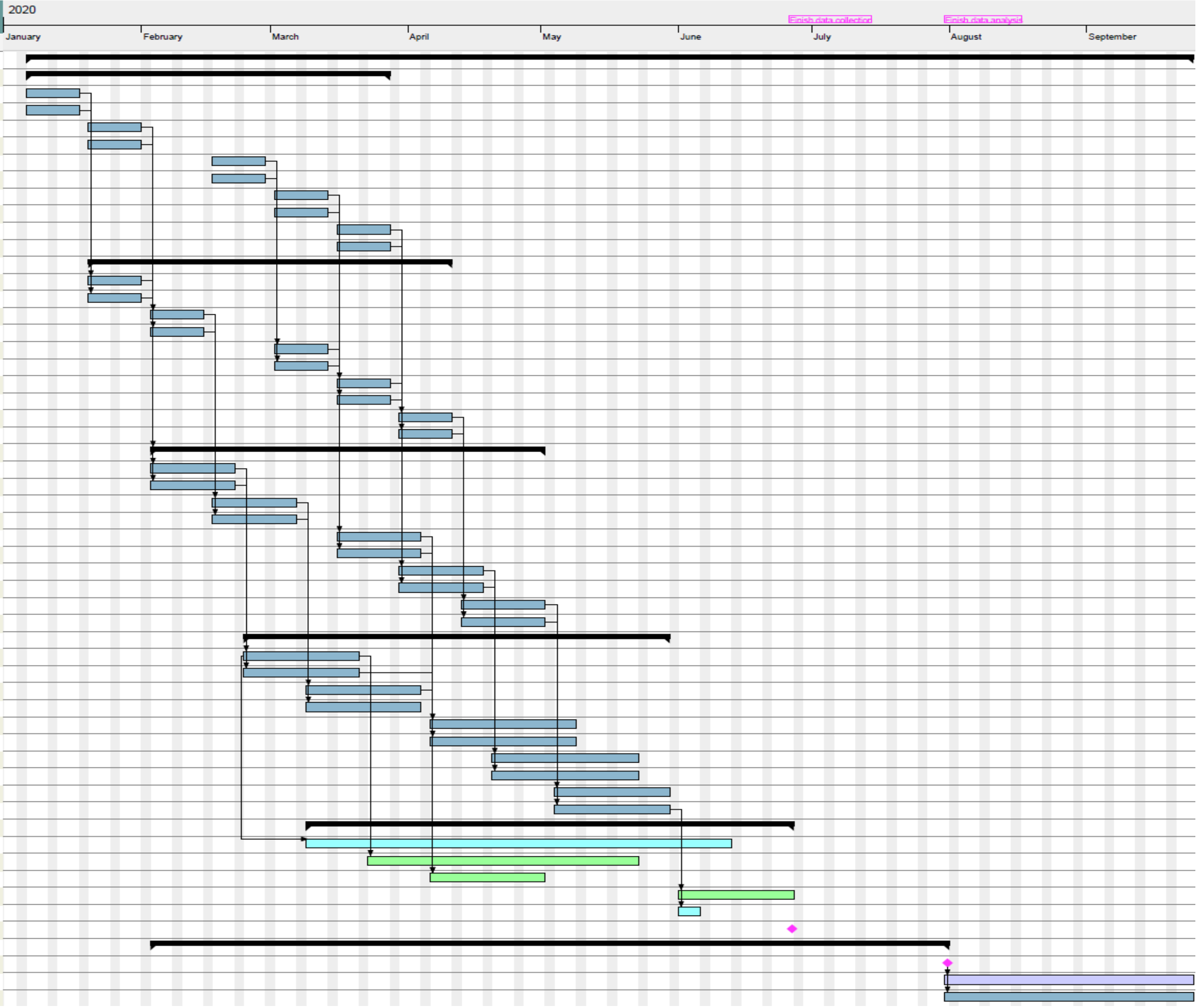

#### Dissemination Plans

We plan to write up this feasibility trial for publication in a peer reviewed journal and to disseminate results at primary care conferences, to all participants, and to the general public via a blog in *The Conversation* or similar. Depending on the results, we will apply for NIHR funding for a full trial of our intervention. After which, our training package could have widespread impact on practitioners' communication skills and patients' pain, quality of life, and satisfaction not only for patients with OA but more broadly for other groups of patients.

#### Patient Public Involvement (PPI)

Our research team includes one PPI representative on the trial management group who works with us as a collaborator, and a number of PPI representatives who contribute to specific aspects of the project and work with us on a consultation basis. In designing this feasibility trial, we have had input from our patient representatives Jennifer Bostock and Jessima Hunter. Appropriate recognition is being given for the contribution made by our PPI representatives, including e.g. payments, honorary contracts, authorships, co-applicant on planned future grants.

#### Ethical Issues

##### Informed Consent to Participate

All participants will receive information and will have the opportunity to ask questions prior to deciding whether to take part. They will be asked for verbal informed consent and/or written informed consent (as outlined above, we are trialling different approaches in this feasibility study), using pre-specified documents. We will ask for consent for using the data they provide for the purposes of this study.

##### Informed Consent for Secondary Research

We will ask for consent for making anonymised questionnaire data and video-recordings of consultations available for secondary research. This is because as part of current moves to make science more open, journals increasingly expect and ask for raw data to be made available. Furthermore, we are collecting a considerable body of filmed consultations in particular that might be a very useful resource for other researchers.

##### Debriefing

All participants will be debriefed verbally (interview participants) and/or in writing (all participants) and offered a copy of study findings.

##### Participant Payments

PCPs will be reimbursed for their time at rates calculated on advice from the CRN and monies will be paid to practices. This is essential to ensure effective recruitment and participation from PCPs. Patient participants will be offered £10 gift vouchers for participating in a telephone interview. This token amount was chosen to convey a sincere thanks without exerting pressure to participate.

##### Assessment and management of Risk

Researchers will follow the University of Southampton and Primary Care Department's Lone working policy when visiting primary care practices in connection with this study. All participants will be made aware that they can withdraw from the study at any time and it is not expected that the topic being discussed will cause them any undue stress.

### Data Management, Data Protection, Data Security

#### Storage and usage of participant data

##### Expression of interest

Expression of interest forms will be stored in a locked filing cabinet at University of Southampton, and destroyed after completion of this study.

##### Audio-recordings of participant interviews

Audio-recordings of participant interviews will be collected using a portable digital recording device. Following each interview, the audio-recordings will be transferred directly to the University of Southampton M drive project folder (accessible only by members of the study team and University of Southampton IT Services) and then deleted from the digital device. The audio data will be anonymised and identified by a unique participant ID only. Transcribing will be facilitated through a member of the research team or a University-approved third party, using only the participant ID. Transcribers will sign a confidentiality agreement to keep the data confidential; store the data securely; and delete the data when the transcription has been completed and receipt confirmed. The audio-recordings will be permanently deleted on study publication.

##### Video-recordings of consultations

Recordings of consultations will be collected using small digital video cameras. These recordings are needed by (a) the PCP who made the recording (for reflection and CPD purposes, and as part of the intervention) and (b) the research team. Therefore, they will be transferred from the recording device to (a) the PCP's work computer and (b) the research team.

To transfer the recordings from the device it will need to be connected to a computer. We will do this in one of two ways.

1) The PCP will connect the device to their work computer. They will then upload the recording(s) to their work computer and share them with the researchers using the University of Southampton safe-send service (formerly known as drop-off).
2) If the PCP's work computer does not permit direct transfer of files from an external device to the computer, then the researcher will attend the practice and upload the recording(s) to their University of Southampton encrypted laptop or an encrypted hard drive. The researcher will then share the recordings with the PCP using the University of Southampton safe-send service.

After successful transfer has been confirmed by the recipient, the recordings will be deleted from the device. The recordings will be transferred to the researchers and deleted from the device as soon as is practicable (within one week maximum).

If electronic file transfer is not possible, the device will be sent to the research team by secure courier.

On receiving the files, the research term will transfer them to the M drive project folder (accessible only by members of the study team and University of Southampton IT Services). Participant ID numbers will be used to label the files and link them to other data collected from the same

PCP/patient. Transcribing, if deemed necessary for analysis, will be facilitated through a member of the research team or a University-approved third party, using only the participant ID. Transcribers will sign a confidentiality agreement to keep the data confidential; store the data securely; and delete the data when the transcription has been completed and receipt confirmed. As outlined above, we will seek consent for archiving the audio-recordings and the transcripts for secondary analysis, as these constitute a potentially valuable resource that could be analysed further. For patients who consent to this, the recordings and transcripts will be stored by the data controller on an encrypted hard-drive and made available to researchers at University of Southampton for secondary research subject to additional ethical approval. For patients who do not consent to archiving for secondary research, the recordings will be permanently deleted on study publication and the anonymised transcripts archived for audit purposes only.

#### Questionnaires

Self-reported questionnaire data will be collected either in paper form or electronically via Qualtrics (as per participant choice). Data received in hard copy will be entered into a password protected database and combined with data downloaded from Qualtrics for analysis. Paper copies will be deleted on study publication. Given moves to enhance transparency in science and data sharing, we will seek patient consent to make questionnaire data available for secondary analysis. For patients who consent to this, their completed questionnaire data will be included in a database to be deposited in an existing secure data archive. A complete copy of the electronic database containing all patients' questionnaire data will be archived for audit purposes only.

#### Personal Data

Participant personal data will be collected and stored securely on a secure server at University of Southampton in compliance with the requirements of the General Data Protection Regulations and the Data Protection Act 2018. Data collected on participants' use of the online intervention will be collected by LifeGuide intervention authoring software and stored on secure, firewall protected servers, hosted by the University of Southampton. Only trained research personnel with specific roles within the project will have access to this server. Upon download, usage data will be stored in an encrypted, password protected file, stored on password protected computers. Personal data will be pseudo-anonymised by assigning a participant identifier code (PIC) which will be used to identify the participant during the study. An electronic file linking the PIC to the identifiable patient data will be kept separately in a separate secure place on the University of Southampton server. Only trained research personnel with specific roles assigned will be granted access to the electronic participant data. Identifiable information will be retained for 10 years after the study has finished in accordance with the procedures agreed by the sponsor. After this time, all identifiable data will be destroyed.

The results of the study will be written up in reports and publications. Anonymised quotations provided by participants during the interviews may be used to illustrate the findings, but participants will not be identifiable.

The anonymised research data (trial master file, transcripts) will be stored for 10 years after the end of the study in accordance with the procedures agreed by the sponsor. During analysis and write-up (approx. 2 years) it will be stored on a secure server or in a locked filing cabinet at University of Southampton, after which it will be stored off site at an approved storage facility that has been agreed by the sponsor.

56. Renner B, Schwarzer R. Risk and Health Behaviors. Documentation of the Scales of the Research Project: "Risk Appraisal Consequences in Korea" (RACK). http://www.gesundheitsrisiko.de/docs/RACKEnglish.pdf: International

University Bremen & Freie Universität Berlin, 2007.
57. Carlsen B, Glenton C. What about N? A methodological study of sample-size reporting in focus group studies. *BMC Med Res Methodol* 2011;11:26-26. doi: 10.1186/1471-2288-11-26
