## Supplementary material for "Feasibility trial of a new digital training package to enhance primary care practitioners’ communication of clinical empathy and realistic optimism": S6

### S6. Appendix: Bespoke Questionnaire Items

#### Practitioner-Reported Self-Efficacy for Communicating Empathy

Please rate how confident you are that you can perform the following behaviours as of now. Rate your degree of confidence by recording a number from 0 to 10 using the scale given below:

| 0 | 1 | 2 | 3 | 4 | 5 | 6 | 7 | 8 | 9 | 10 |
| --- | --- | --- | --- | --- | --- | --- | --- | --- | --- | --- |
| cannot do at all |  |  |  |  | moderately certain can do |  |  |  |  | highly certain can do |

1. Can convey empathy to patients who don’t help themselves
2. Can convey empathy to patients who are antagonistic
3. Can convey empathy when you are running late
4. Can convey empathy to patients you find difficult to like
5. Can convey empathy to patients who are very different to you
6. Can convey empathy to patients who disagree with you
7. Can convey empathy when you are feeling tired, angry, or frustrated

Scale score = Mean across all 7 items

#### Practitioner-Reported Self-Efficacy for Communicating Optimism

Please rate how confident you are that you can perform the following behaviours as of now. Rate your degree of confidence by recording a number from 0 to 10 using the scale given below:

| 0 | 1 | 2 | 3 | 4 | 5 | 6 | 7 | 8 | 9 | 10 |
| --- | --- | --- | --- | --- | --- | --- | --- | --- | --- | --- |
| cannot do at all |  |  |  |  | moderately certain can do |  |  |  |  | highly certain can do |

1. Can convey realistic optimism when you are running late
2. Can convey realistic optimism to patients who do not appear to be optimistic
3. Can convey realistic optimism to patients who have tried previous treatments with little success
4. Can convey realistic optimism to patients who came in wanting a different treatment to the one you are recommending
5. Can convey realistic optimism to patients with quite rigid expectations about treatment

Scale score = Mean across all 5 items

#### Practitioner-Reported Intention to Change Communication of Empathy and Optimism

Please select an option for each statement.

1. I expect to make the changes that I have set myself as part of EMPathicO
2. I want to make the changes that I have set myself as part of EMPathicO
3. I intend to make the changes that I have set myself as part of EMPathicO

| 1 | 2 | 3 | 4 | 5 | 6 | 7 |
| --- | --- | --- | --- | --- | --- | --- |
| Strongly disagree |  |  |  |  |  | Strongly agree |

Scale score = Mean across all 3 items

#### Practitioner-Reported Outcome Expectancies for Changing Communication of Empathy and Optimism

If I make the changes that I have set myself as part of EMPathicO then:

| 1a | My patients will feel better | Likely | ⭘ | ⭘ | ⭘ | ⭘ | ⭘ | ⭘ | ⭘ | Unlikely |
| --- | --- | --- | --- | --- | --- | --- | --- | --- | --- | --- |
| 1b | My patients feeling better is… | Good | ⭘ | ⭘ | ⭘ | ⭘ | ⭘ | ⭘ | ⭘ | Bad |
| 2a | My patients will be more satisfied with the care I provide | Likely | ⭘ | ⭘ | ⭘ | ⭘ | ⭘ | ⭘ | ⭘ | Unlikely |
| 2b | My patients being more satisfied with the care I provide is… | Good | ⭘ | ⭘ | ⭘ | ⭘ | ⭘ | ⭘ | ⭘ | Bad |
| 3a | My patients will feel more cared for | Likely | ⭘ | ⭘ | ⭘ | ⭘ | ⭘ | ⭘ | ⭘ | Unlikely |
| 3b | My patients feeling more cared for is… | Good | ⭘ | ⭘ | ⭘ | ⭘ | ⭘ | ⭘ | ⭘ | Bad |
| 4a | My patients will feel more optimistic about their treatment | Likely | ⭘ | ⭘ | ⭘ | ⭘ | ⭘ | ⭘ | ⭘ | Unlikely |
| 4b | My patients feeling more optimistic about their treatment is… | Good | ⭘ | ⭘ | ⭘ | ⭘ | ⭘ | ⭘ | ⭘ | Bad |
| 5a | My patients’ symptoms/response to treatment/self-management will improve | Likely | ⭘ | ⭘ | ⭘ | ⭘ | ⭘ | ⭘ | ⭘ | Unlikely |
| 5b | My patients’ symptoms/response to treatment/self-management improving is… | Good | ⭘ | ⭘ | ⭘ | ⭘ | ⭘ | ⭘ | ⭘ | Bad |
| 6a | I will feel more satisfied with the care I provide | Likely | ⭘ | ⭘ | ⭘ | ⭘ | ⭘ | ⭘ | ⭘ | Unlikely |
| 6b | My being more satisfied with the care I provide is… | Good | ⭘ | ⭘ | ⭘ | ⭘ | ⭘ | ⭘ | ⭘ | Bad |
| 7a | I will feel more emotionally drained by my patients | Likely | ⭘ | ⭘ | ⭘ | ⭘ | ⭘ | ⭘ | ⭘ | Unlikely |
| 7b | My feeling more emotionally drained by my patients is… | Good | ⭘ | ⭘ | ⭘ | ⭘ | ⭘ | ⭘ | ⭘ | Bad |
| 8a | I will feel more resilient | Likely | ⭘ | ⭘ | ⭘ | ⭘ | ⭘ | ⭘ | ⭘ | Unlikely |
| 8b | My feeling more resilient is… | Good | ⭘ | ⭘ | ⭘ | ⭘ | ⭘ | ⭘ | ⭘ | Bad |

Scoring. ‘a’ items scored 1 (unlikely) to 7 (likely). ‘b’ items scored +3 (good) to -3 (‘bad’). Reverse score item 7a and 7b. Compute 1a x 1b, 2a x 2b, 3a x 3b, etc. Scale score = Mean (a.b)

#### Patient-Reported Perceptions of Practitioners’ Response Expectancies

Version for patients reporting a treatment recommendation as a consultation outcome

Thinking about today’s consultation, how optimistic was the clinician that your treatment will help you?

| ⭘ | Extremely pessimistic (the clinician is certain that it will not help me) |
| --- | --- |
| ⭘ | Very pessimistic (the clinician seemed confident it would not help me) |
| ⭘ | Somewhat pessimistic (the clinician seemed to think it might not help me) |
| ⭘ | Neutral (the clinician seemed to think it might help me but it might not) |
| ⭘ | Somewhat optimistic (the clinician seemed to think it might help me) |
| ⭘ | Very optimistic (the clinician seemed confident it would help me) |
| ⭘ | Extremely optimistic (the clinician is certain that it will help me) |

Version for patients reporting no treatment recommendation as a consultation outcome

Thinking about today’s consultation, how optimistic was the clinician?

| ⭘ | Extremely pessimistic |
| --- | --- |
| ⭘ | Very pessimistic |
| ⭘ | Somewhat pessimistic |
| ⭘ | Neutral |
| ⭘ | Somewhat optimistic |
| ⭘ | Very optimistic |
| ⭘ | Extremely optimistic |
