## Supplementary material for "Feasibility trial of a new digital training package to enhance primary care practitioners’ communication of clinical empathy and realistic optimism": S7

### S7. Appendix: Topic Guide for Practitioner Interviews

**TOPIC GUIDE (Practitioners)**

**Briefing for interview**

1. Thank you for agreeing to take part in a research interview today. I am (*researcher name*), a researcher at University of Southampton.
2. The aim of this interview today is to explore your views of participating in the TIP study (Talking in Practice).
3. With your agreement I will audio-record our conversation. The recording will be transcribed but everything you say will be anonymous. I am only recording it so I don’t miss anything and make sure I have what you say written up accurately. Your name along with any names you mention, any places you mention and all other identifiable information will be taken out, so that if someone heard or read your interview they would not know who you are or where you work.
4. Your interview will remain confidential.
5. If at any time you do not wish to answer a question that’s okay, and if at any stage you wish me to stop the recorder, please let me know.
6. We can take a break at any time – let me know and I will stop the recording. We can either continue after the break, arrange another time to meet or stop there.
7. I would like to encourage you to be as honest as you can. There are no right or wrong answers. I am not here to judge you I am simply interested in your views and experiences.
8. Do you confirm that you consent to taking part in this research interview? *Take verbal consent (researcher to read out consent script over the telephone and record consent on verbal consent form)*
9. **TURN ON THE RECORDING DEVICE**

*Below are example questions which will be adapted for focus group sessions. The topic guide will evolve as the interviews and focus groups progress and may change in response to specific feedback from participants and as the TIP study progresses.*

**Topics of interest**

A. **Being approached for the study**
   a. What encouraged you/the practice to take part in the TIP study?
      i. Explore positive and negative views about the study in general.
   b. What were your expectations of taking part?
   c. Can you describe any reservations about taking part in the TIP study?
   d. How would you encourage other practices/practitioners to take part in a main trial?
B. **Baseline video recordings (if done):**
   a. Can you describe the process of obtaining the baseline video-recordings at your practice?
      i. How did you approach patients? What worked best for in your practice? Opportunistic recruitment? advanced booking?
      ii. Was there anything the study team could have done to improve this process?
   b. How did patients respond to being asked to participate? What were the main reasons for patients not participating in the baseline video-recordings?
   c. How did the consent process work in your practice? Who took consent? What problems did you encounter? How did you overcome any problems?
   d. Can you describe any problems you encountered when video-recording your consultations? What could be done to improve this process?
   e. What effect did video-recording have on your consultations i.e. time, intrusion etc
   f. How did you feel about video-recording your consultations.
   g. How were video-recording managed and transferred to University of Southampton? Any problems and how were they overcome?
C. **Views of video-recording consultations**
   a. How do you feel about video-recording your consultations with patients?

What do you see as the main challenges? How could we overcome them?

D. **Randomisation (if done):**
   a. How was randomisation communicated to you in your practice?
   b. What do you think about cluster randomisation? (randomisation at the practice level rather than the practitioner level?). What do you see as the benefits/downsides?
   c. How did you feel about your group allocation?
   d. Control group – how likely are you to use the EMPATHICO intervention at the end of the study?
E. Patient recruitment methods (if done in practice)
   a. What do you think about the patient recruitment methods? Researcher in waiting room? Advance mailout?
F. **Intervention Practices only**
   a. Please could you tell me your experiences of using EMPATHICO?
      i. Did you complete the EMPATHICO training intervention? All in one go? Over different sessions?
      ii. How easy was it to use? What made it easier, what made it more difficult? Where did you access the training (e.g. at home/at the practice?)
      iii. What were the main things that you liked/disliked about it?
           1. **Empathy module**
              What did you like about this section? What did you dislike about this section? What was new? What parts did you find most relevant/least relevant?
              In the current climate of remote consultations, which parts of the empathy module would be relevant, usable in telephone consultations? Video consultations? Face to face consultations when wearing PPE?
           2. **Optimism module**
              What did you like about this section? What did you dislike about this section? What was new? What parts did you find most relevant/least relevant?
              In the current climate of remote consultations, which parts of the optimism module would be relevant, usable in telephone consultations? Video consultations? Face to face consultations when wearing PPE?
           3. **Osteoarthritis module**

What did you like about this section? What did you dislike about this section? What was new? What parts did you find most relevant/least relevant?
In the current climate of remote consultations, which parts of the OA module would be relevant, usable in telephone consultations? Video consultations? Face to face consultations when wearing PPE?

4. **Reflection**
   Did you complete the reflections module?
   What did you like about this section? What did you dislike about this section? Please describe how you reflected on your baseline consultations. Video? Reflecting back on a recent consultation? How easy was this? Useful?
5. **Goal setting**
   Did you set yourself any goals of things you might change after completing EMPATHICO?
   Could you describe how you have implemented any of the goals? How did you find that?
   How did you implement them in a telephone/video/face to face consultations when wearing PPE?? What challenges did you encounter
iv. Please could you describe whether using EMPATHICO had any impact on your consultations?
    1. What were the main changes that you identified after completing the training?
    2. How successful do you feel you were in implementing the changes? If unsuccessful, what were the main reasons?
v. Overall, which sections were most relevant/least relevant to your clinical practice? Are there any sections that you missed? Why?
vi. What would encourage you to use implement the things you learnt during the training?
vii. What might be the barriers to using EMPATHICO in wider clinical practice?
viii. Would you recommend the training to other colleagues? Other PCPs?

G. **Completing questionnaires (if done)**
   a. Can you tell me about your experience of completing the TIP study questionnaires? How long did they take? Were there any questionnaires that you found difficult to complete? Did not seem appropriate?
   b. What could be improved?
H. **Further thoughts and anything to add**
   a. What aspects of the study do you feel could be improved?
   b. What aspects of the study did you particularly like/dislike?
   c. Is there anything else you would like to share about your experiences of taking part in the TIP study?

**Debrief**

- Ask if the participant has any questions about the study.
- Revisit consent – still happy for their interview to be used (anonymously)?
- **TURN OFF THE RECORDER**
- **Thank** participant for taking part in the interview
- **Offer copy of transcript when available**
- **Offer copy of study findings when available**
