## Supplementary material for "Feasibility trial of a new digital training package to enhance primary care practitioners’ communication of clinical empathy and realistic optimism": S8

### S8. Appendix: Topic Guide for Patient Interviews

**Briefing**

1. Thank you for agreeing to take part in a research interview today. I am (*researcher name*), a researcher at University of Southampton.
2. The aim of this interview today is to explore your views of participating in the TIP study (Talking in Practice).
3. With your agreement I will audio-record our conversation. The recording will be transcribed but everything you say will be anonymous. I am only recording it so I don’t miss anything and make sure I have what you say written up accurately. Your name along with any names you mention, any places you mention and all other identifiable information will be taken out, so that if someone heard or read your interview they would not know who you are or where you work.
4. Your interview will remain confidential.
5. If at any time you do not wish to answer a question that’s okay, and if at any stage you wish me to stop the recorder, please let me know.
6. We can take a break at any time – let me know and I will stop the recording. We can either continue after the break, arrange another time to meet or stop there.
7. I would like to encourage you to be as honest as you can. There are no right or wrong answers. I am not here to judge you I am simply interested in your views and experiences.
8. Do you confirm that you consent to taking part in this research interview? *Take verbal consent (researcher to read out consent script over the telephone and record consent on verbal consent form)*
9. **TURN ON THE RECORDING DEVICE**

*The topic guide will evolve as the interviews progress and may change in response to specific feedback from participants and as the TIP study progresses.*

**Topics of interest**

A. **Thoughts about your appointment**

I would like to ask you a bit about your appointment. You have told us a lot already by completing the questionnaires, but we would like to make sure we understand your experiences correctly and in more detail:

a. **Practicalities:
   Thinking about the appointment you had with your GP/nurse/physiotherapist:**
   i. How did you initially make your appointment?
   ii. (*Explore process of appointment/follow-up)*
   iii. How did the technology work? Did you have any problems? What device did you use for the appointment? (Phone/computer/tablet?)
   iv. Have you experienced remote consultations before (telephone/video/face-to-face?)?
b. **Overall views**:
   What was your overall thoughts about your telephone/video/face to face appointment. What did you like about it? Dislike? How did it compare with other consultations you’ve had in the past. What worked well, what didn’t work quite so well?
c. ***Before the interview – check through the patients answers to the CARE, MISS and treatment outcomes and not general views of the consultation and optimism with treatment. Note anything scored particularly low to explore in the interview. Refer to this when asking the following questions:***

**Thinking back to the appointment itself:**

i. How comfortable or at ease did you feel with the appointment?
   1. Can you tell me a bit more about that? What did they do or say to make you feel at ease?
ii. How did you feel about telling the doctor about your problems and concerns during the appointment?
    1. How was the doctor at letting you tell your story?
    2. How was this compared to your normal consultation in the surgery?
iii. How was the doctor at listening to you?
     1. What made you feel that the doctor was really listening?
     2. What else could they have done?
     3. How did that compare to your usual face to face consultations?
     4. Did you have a sense that they were using the computer?
iv. To what extent did you feel that the doctor knew about you and your situation? *(check if consulted with this person before)*
    1. What else do you feel would have been useful for them to know or find out before your appointment?
v. To what extent do you think the doctor understood your concerns?
   1. Can you tell me a bit more about how they showed you they understood?
vi. How was the doctor at showing you care and compassion?
    1. How did that compare to your usual consultations?
    2. What more could they do?
vii. How positive or optimistic was the doctor in the appointment?
     1. What sort of things did they do or say that appeared positive?
     2. How did that make you feel?
viii. How was the doctor at explaining things?
      1. How was the doctor with explaining your problem?
      2. How did they explain how your treatment would work?
      3. How optimistic were they the treatment would work for you?
ix. How was the doctor at making a plan of action with you?
    1. Can you tell me a bit about what plans you made with the doctor if your treatment did not work?
    2. How did the appointment finish? What was the last thing that they said and how did that make you feel?

B. **Being approached for the TIP study**
   a. Can you tell me where you saw the advert to take part in the TIP study?
   b. What was your understanding of what the research was about?
   c. What other information might have been helpful at that stage?
C. **Deciding to take part**
   a. Why did you decide to take part? What was your motivation?
   b. Can you tell me about any reservations you had about taking part?
D. **Completing questionnaires

   *Before the interview, check completion rates and whether any questions were missing.***
   a. Can you tell what you thought about completing the TIP study questionnaires?
      i. What device (Phone/tablet, computer) did you use to complete them?
      ii. Roughly how long did they take?
      iii. How did you feel overall about completing them?
   b. The questionnaires covered a range of topics including:

Your appointment

Your treatments

Your overall health

Thoughts and feelings

What did you think about completing these questions?

Did you have any difficulties in completing any of the questions?

How relevant did you find the questions?

What could we do to make it easier for patients to complete the questions?

*For patients who consulted for OA*: what did you think about the questions about your hip or knee pain? How relevant do you think they were?

c. Non-completers:
   i. We notice that you didn’t complete all the questions. Could you tell me a bit about why you didn’t finish the questionnaires? How could we encourage people to complete the questionnaires?
d. How was your experience of receiving follow up questionnaires
e. *If the second set of questionnaires were not completed:* If no, then what could we do to improve things for future patients?
f. What could be done to improved questionnaire completion?

E. **Further thoughts and anything to add**
   a. What aspects of the study do you feel could be improved?
   b. What aspects of the study did you particularly like/dislike?
   c. Is there anything else you would like to share about your experiences of taking part in the TIP study?

**Debrief**

- Ask if the participant has any questions about the study.
- Revisit consent – still happy for their interview to be used (anonymously)?
- **TURN OFF THE RECORDER**
- **Thank** participant for taking part in the interview
- **Offer copy of transcript when available**
- **Offer copy of study findings when available**

*The topic guide will evolve as the interviews progress and may change in response to specific feedback from participants.*
